# Impact of climate and acquired immunity on malaria burden in Senegal and The Gambia: an agent-based modeling study

**DOI:** 10.64898/2026.09.18.26363423

**Authors:** Miguel Garrido Zornoza, Cyril Caminade, Amady Dieng, Nakul Chitnis, Adrian M. Tompkins

## Abstract

Immunity against *Plasmodium falciparum* malaria is acquired slowly and incompletely through repeated infections, leading to differing levels of protection among individuals based on their history of exposure to the disease. Since malaria is a climate sensitive vector-borne disease, immunological history is shaped by the local climate. Nonetheless, compartmental malaria models, which may be used to inform policy, implicitly represent immunity as a population-average trait — an assumption that can lead to incorrect estimates of clinical prevalence by age, and of the overall burden of malaria. As the rate at which immunity is acquired depends on transmission intensity, and therefore on background climatic conditions, the error introduced by this assumption is expected to be climate-sensitive. We quantify how individual exposure histories shape community-level prevalence, and identify the age groups for which resolving immunity individually matters most across climatic regimes. For this, we introduce VECTRI-ABM, a grid-based framework coupling the climate-driven vector component of the VECTRI model to an agent-based representation of the human population, resolving immunity at the individual level. The framework was calibrated against entomological and epidemiological observations from Dielmo and Ndiop (Senegal, 1990–2003) and evaluated against reported entomological inoculation rates, Malaria Atlas Project estimates of parasite rate in children aged 2–10, and confirmed cases from Senegal’s National Malaria Control Program (*PNLP*). We then applied it across Senegal and The Gambia, which encompass a pronounced Sahelian-to-tropical rainfall gradient, and compared simulations in which immunity develops individually against a counterfactual where agents carry the local population-average immunity, thus mimicking the assumption made by standard models. Results show that the averaging approach misallocates burden across age, underestimating clinical prevalence in children, especially so in the wettest high-transmission regions, and overestimating adult asymptomatic reservoirs, with implications for transmission-targeting interventions. We find that the simpler approach to immunity is adequate for immunologically mature populations, typically above 15 years of age, and that the error it introduces elsewhere is climate-dependent.

**Author summary:** Malaria does not affect everyone equally. Protection against the parasite causing the disease builds up slowly, from repeated exposure over a person’s lifetime, so a child and an adult respond differently to the same infectious mosquito bite. This accumulation depends on how much malaria is circulating, which in turn depends on rainfall, since rain determines where mosquitoes can breed. Most models of the disease simplify this defining trait by giving everyone the same, average level of protection — we wanted to study the errors induced by this simplification. For this, we developed a novel framework that follows millions of people through time, each with their own history of exposure to malaria, and that drives mosquito populations with historical climate records in Senegal and The Gambia. We used it twice: once letting protection develop individually, and once forcing everyone to share the community average. We found the averaged approach fails to represent burden by age, underestimating illness in children, especially where transmission is intense. Above 15 years of age, both versions largely agree on clinical illness, with the average slightly overestimating how many adults carry the parasite without symptoms. Overall, a simple description of immunity misjudges disease burden as a function of age and local climate.

## Introduction

Malaria is a parasitic mosquito-borne infectious disease caused by 5 *Plasmodium* species, from which *Plasmodium falciparum* is the deadliest [1]. The latter is transmitted through the bite of infected *Anopheles* mosquitoes and, in particular, of the species *Anopheles gambiae s*.*s*. (*sensu stricto*), hereafter *An. gambiae*, which is widespread across most sub-Saharan Africa [2] and constitutes one of the most important malaria vectors in the continent [3]. As with many arthropod-borne diseases, climate plays a critical role in the distribution, intensity and duration of malaria transmission [4], especially in epidemic fringe regions with pronounced rain- or temperature-driven seasonalities [5, 6]. To this date, malaria continues to exert a large impact on global health, with annual estimates of 282 million cases and 610000 deaths in 2024, from which the WHO African region has a disproportionate contribution, carrying 95% of the global burden [1]. In this region, children under 5 years of age account for 75% of deaths [1], a reflection of an age-stratified vulnerability, partly driven by differential immunity against the disease.

Unlike many viral infections, where a single exposure may trigger life-long protection against the disease, immunity against malaria is acquired slowly and incompletely throughout an individual’s life via repeated infections, vanishing when exposure ceases [7]. This creates a hysteresis effect in the response against the disease, with varying levels of acquired immunity potentially modulating the sensitivity of the disease to climate [8, 9]. In this context, mathematical malaria models serve as essential tools bridging basic, conceptual mechanisms with population-level and region-specific observations. Such models have been used to delineate the current and future geographical range of the disease [10–12], to identify key factors driving disease dynamics [13], and to optimize and evaluate the effect of diverse intervention strategies [14]. Models explicitly considering the combined effect of climate and immunity on disease transmission dynamics are, however, scarce. The temperature-driven SEIR (Susceptible-Exposed-Infectious-Recovered) compartmental model from [15] included additional recovered states to represent boosting in immunity, with little resulting effect on simulated infection incidence. Similarly, [16] incorporated partial immunity and age structure, explicitly considering rainfall and temperature, to find only marginal added effects on malaria incidence. In contrast, work by [8], clearly illustrated the importance of capturing the interaction between immunity and climate, particularly in endemic settings, where immunity acted to buffer the effects of climate variability on malaria incidence.

Capturing this interplay between climate and individually-acquired immunity, however, poses structural challenges for compartmental approaches. Model design and structure generally support the specific objectives for which the model is developed [17], strategically omitting or simplifying supposedly secondary physical, biological or demographic factors. For instance, compartmental models effectively work with population-level averages, making them computationally efficient and suitable for estimating broad epidemiological trends, yet they are limited when such averages fail to represent individual-level traits relevant for malaria incidence at finer scales [17]. In this sense, Agent-Based Models (ABMs) represent alternative frameworks to capture fine-scale heterogeneities that modulate transmission. These frameworks have been used to evaluate the efficacy and cost-effectiveness of a range of interventions [18–23] as well as to incorporate host-vector interactions, such as individual attractiveness to mosquitoes [24]. Furthermore, ABMs allow for a finer representation of the human host biology, including both maternal [19, 20] and acquired immunity [9, 18, 24], and the varying duration of infections [25]. They can also include demographic factors like host age - used for example as proxy for biting rates, as it is correlated to the body surface area [18, 25].

Beyond individual-level traits, ABMs differ in their representation of the environment. While some ABMs incorporate transmission seasonality using theoretical periodic or sinusoidal forcings [26], others use realistic environmental and demographic drivers. These include the availability of water sources for oviposition and larval development [27], house location to inform blood-meal probabilities [22, 25] and meteorological data - such as rainfall and air temperature - to drive transmission seasonality [28–30]. Recently, Martin-Makowka *et al*. [31] coupled the OpenMalaria model to VECTRI [32] showing that climate inter-annual variability affects the modeled effectiveness of seasonal indoor residual spraying and its optimal deployment time. Along these lines, the work by Yamana *et al*. [9] integrated environmental data - rainfall, air temperature and topography - with immunity acquisition dynamics to conclude that immunity acted to damp differences in malaria prevalence among two entomologically different villages. Most ABM frameworks remain focused on specific [25, 27, 33] or hypothetical [21, 22] village settings. While some grid- and patch-based systems have been used to model relatively larger areas [25, 33–37], they mostly cover the immediate region surrounding a particular settlement, leaving a gap in the application of mechanistic, immunity-aware ABMs at a truly regional scale.

Using a climate-driven, individual-based modeling approach, we here address two related questions: (i) how does an individual’s cumulative history of exposure to malaria shape community-level prevalence, and does this relationship vary across different environmental conditions? (ii) which age groups carry the highest burden in a given environmental context, and for which is it crucial a fine-scale representation of acquired immunity? To answer these we contrast individual- and population-level representations of immunity, quantifying the biases that arise when individual-level immunity is replaced by a population-level average. For this, we extended and used the VECtor-borne disease community model of ICTP, TRIeste (VECTRI) model [32] by substituting its original compartmental SEIR component with an agent-based module. The new, grid-based mechanistic model, VECTRI-ABM, explicitly resolves the climate-sensitive vector life cycle while capturing individual-level immunity dynamics. The model was calibrated to entomological and clinical data from Senegal, and subsequently validated using spatiotemporal prevalence and incidence data from individual studies, from the official routine surveillance system of Senegal’s National Malaria Control Program (*Programme National de Lutte contre le Paludisme, PNLP*) and from alternative model estimates produced by the Malaria Atlas Project [38]. After calibration and evaluation, the model was used at a regional scale in Senegal and The Gambia. This region exhibits the highest variance in entomological inoculation rates (EIRs) [39], driven primarily by the rain-sensitive vector *An. gambiae* (Fig 1). Located at the interface between the Sahel in the north, a semi-arid region with very sparse rainfall, and a tropical climate in the south, the study area consequently presents a pronounced, rain-driven north-south gradient in *An. gambiae* abundance and malaria endemicity, thus constituting a perfect scenario to evaluate the effect of acquired immunity across a broad range of rainfall conditions.

**Fig 1.**
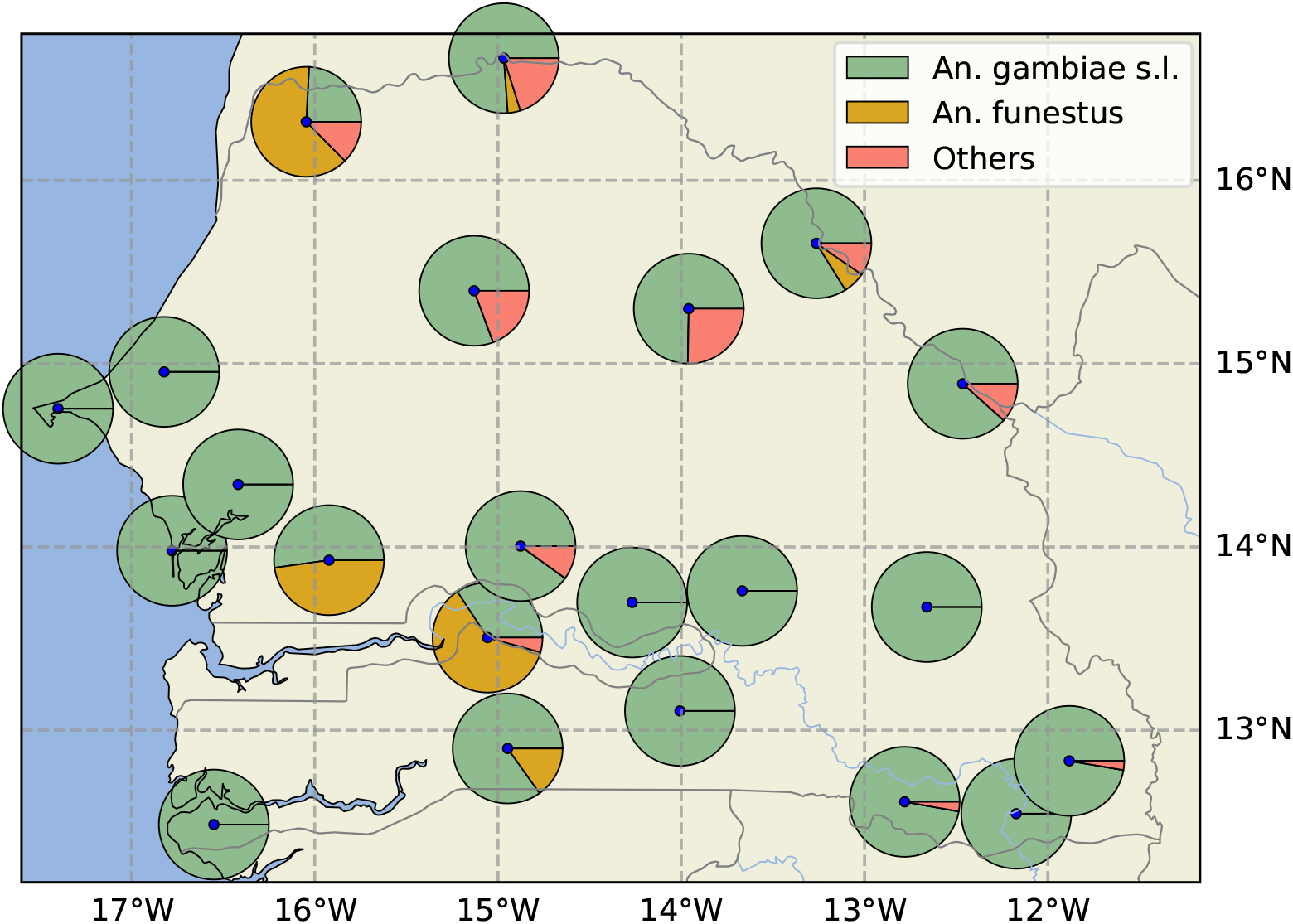
Malaria vector species distribution in Senegal as of 2020, digitized from [40]. *s*.*l*. ≡ *sensu lato*.

## Materials and Methods

### Agent-based model of human health

The complete model description following the ODD (Overview, Design concepts, Details) protocol [41, 42] can be found in S0. Agent-Based Model. We here provide an outline of the primary structure of the new VECTRI-ABM framework.

#### Malaria module

*P. falciparum* dynamics in the human host is simulated with a susceptible (*S*), exposed (*E*), asymptomatic (*A*), symptomatic (*I*), and recovered (*R*) model as follows (see Fig 2): when an initially susceptible agent receives an infectious bite it is infected with some probability *P*_*V* →*h*_ (*S* → *E*). Upon exposure, the agent undergoes an intrinsic incubation period (IIP) of 10 days [43] and then transitions to a symptomatic state (*E* → *I*) with a probability, *α*(*i*_*m*_), function of its immunity level, *i*_*m*_ ∈ [0, 1]. Following [44], we model *α*(*i*_*m*_) as a sigmoidal function of *i*_*m*_ (S0.7 - Submodels). This is

**Fig 2.**
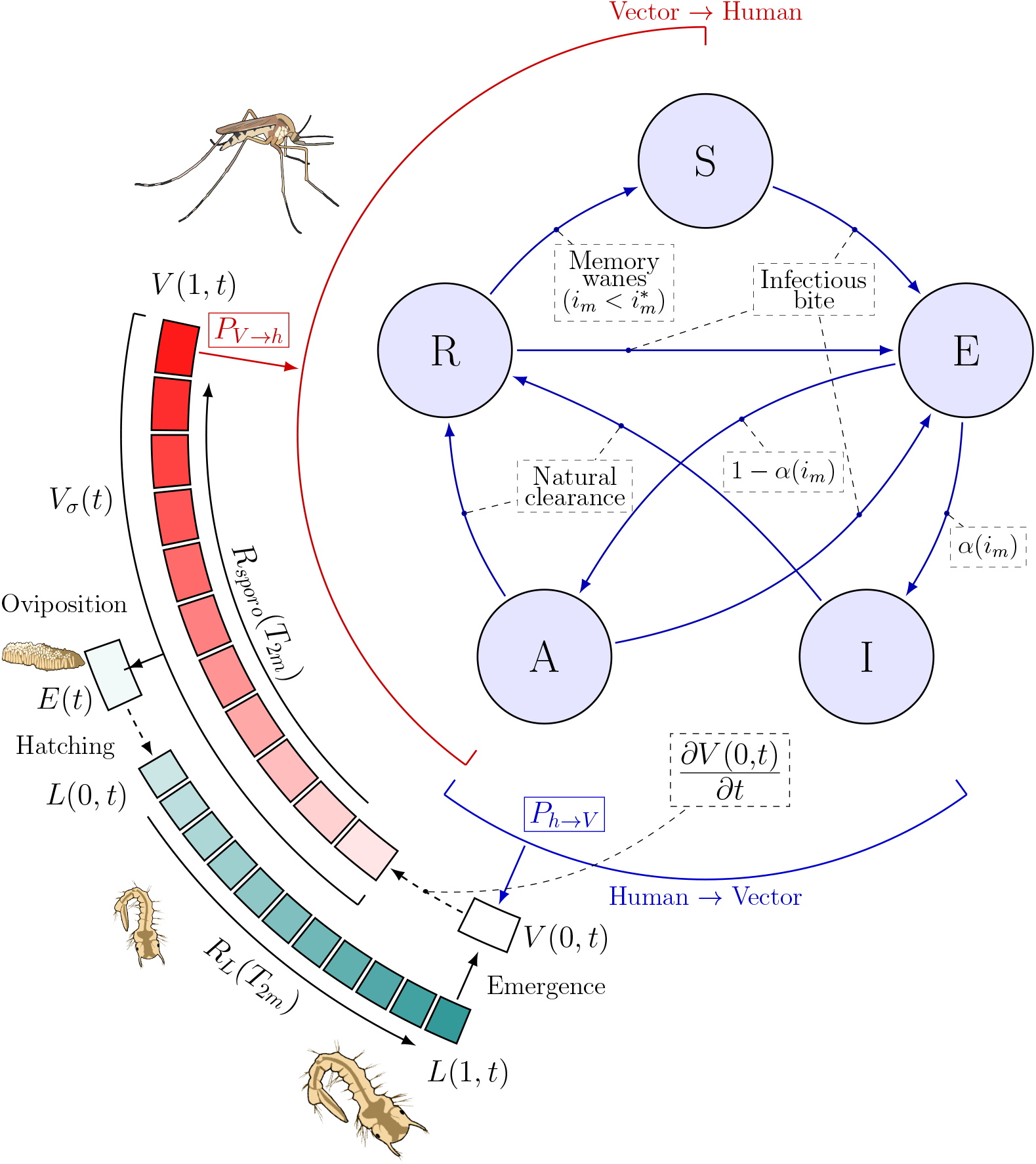
Framework schematics. Illustrations from NIAID NIH BioArt Source [53–55].

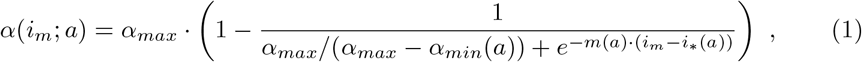

with (*a*) denoting age-dependent parameters, such as the pyrogenic threshold, *i*_∗_(*a*). When the immunity level is low, most agents shall transition to this state, progressively shifting towards asymptomatic infections (*E* → *A*) as they acquire immunity. The agent is asymptomatic with probability 1 − *α*(*i*_*m*_). Newborn agents additionally benefit from maternal immunity, a short-lived protection of ∼ 3 − 9 months [45, 46] transferred from immune mothers to their offspring [7] that renders them immune to clinical malaria while potentially carrying low-density asymptomatic infections and thus remaining infective to vectors [47]. Maternally protected agents thus always follow the *E* → *A* transition, regardless of *i*_*m*_, and remain infective to vectors. This protection is lost exponentially at a half-life rate 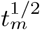 [45], after which the agent enters the standard SEIAR dynamics. Age-related effects and immunity dynamics, *d*(*i*_*m*_(*t*))*/dt*, are discussed in further details in S0. Agent-Based Model. A relevant advantage of using agents to represent groups or individuals is the possibility of straightforwardly resolving non-exponentially distributed events for arbitrary distributions. Bulk mechanistic models implicitly assume exponentially-distributed waiting times or resolve gamma-distributed transitions with the costly technique of decomposing single states into multiple intermediates [48]. From the agent perspective this is handled by assigning a counter, describing the time left for the event. The initial value of the counter is drawn from the particular distribution. In VECTRI-ABM, upon infection the duration of the disease (days) is drawn from a log-normal distribution,

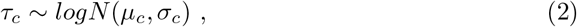

whose parameters are interpolated from log-normal distributions of typical clearance times in naive (*Pf* PR ∼ 0%) [49] and highly endemic (*Pf* PR ∼ 75%) [50] populations (S1.a Clearance times), as

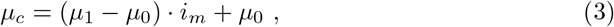

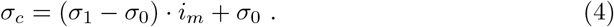

With (*µ*_0_, *σ*_0_) being the parameters of a log-normal distribution of clearance times of naive individuals and (*µ*_1_, *σ*_1_) those of individuals in the highly endemic region.

Gametocyte maturation in the bone marrow is generally considered to require 10–12 days [47], after which mature gametocytes travel to the peripheral blood, making the host infective to vectors. The sexual pathway, or gametocytogenesis, can however be present immediately after asexual *P. falciparum* low-density infections [51, 52], potentially at high levels before symptom onset and detection. These presymptomatic subjects already carrying mature gametocytes are here considered to belong to the exposed state, *E*. In the model, we simply assume agents to be fully infective to vectors immediately after the IIP. When the counter *τ*_*c*_ = 0 the agent is considered to have *cleared* the disease and transitions to the recovered state (*I, A* → *R*). Agents in asymptomatic or recovered states may transition to exposed (*A, R* → *E*) following an infectious bite. When the immunity level of a given agent is below a threshold value, *i*^∗^, the agent transitions back to a susceptible state (*R* → *S*). Further details on immunity dynamics can be found in S0.7 - Submodels.

#### Climate-driven vector model: VECTRI

To represent the vector component of the parasite’s life cycle we used the vector module of the VECTRI model [32]. VECTRI is a grid-based climate-sensitive compartmental model of vector ecology. At any particular location, (*lon, lat*), the dynamics of the female mosquito in its adult (*V* ), egg (*E*) and larval (*L*) stages is modeled as the set of coupled non-autonomous differential equations as follows:

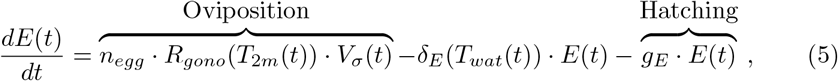

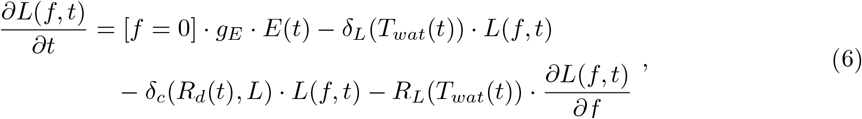

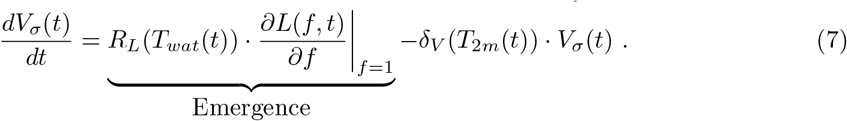

Here, *n*_*egg*_ is the average number of laid eggs per batch that result in female vectors, *R*_*gono*_ (day^−1^) the rate of the gonotrophic cycle, *δ*_*i*_ (day^−1^) decay rates associated to temperature (*i* = *E, V, L*) and crowding effects (*i* = *c*), *g*_*E*_ (day^−1^) the egg hatching rate, [∗] the Iverson bracket with *f* describing the fractional developmental stage of larvae, whose evolution is described by an advection equation along *f*, bounded to [0, 1], and *R*_*L*_ (day^−1^) the advection velocity of larval development. The dependencies *T*_2*m*_, *T*_*wat*_ and *R*_*d*_ are the air temperature at two meter height (^◦^*C*), the pond water temperature (^◦^*C*) and the daily rainfall (*mm/day*), respectively. The subscript *σ* in the vector density stands for the sum of vectors across all sporogonic stages. Analogously to the larval scheme, the growth of the parasite inside the vector is captured by an advection equation of the fractional developmental stage, *f*_*s*_, this is

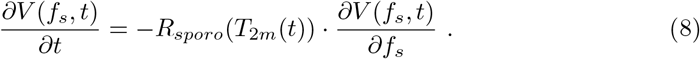

With *R*_*sporo*_(*T*_2*m*_(*t*)) (day^−1^) representing the temperature-dependent growth rate of the parasite. Intermediate values, *f*_*s*_ ∈ (0, 1), represent stages of parasite development where the vector is not yet infective to human hosts. With this nomenclature *V* (0, *t*) and *V* (1, *t*) stand for the density of susceptible and infective vectors, respectively. Eq(8) applies to *f*_*s*_ ≠ 0. The evolution of the case *f*_*s*_ = 0 requires special attention, as it interacts with the agent module. For a probability of human-to-vector transmission *P*_*h*→*V*_, the flux of newly-infected vectors reads as

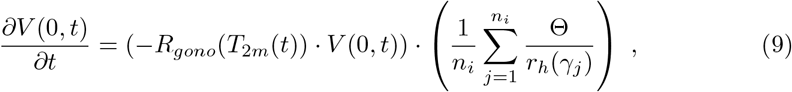

where Θ is a stochastic variable equal to 1 with probability *P*_*h*→*V*_, and zero otherwise. *r*_*h*_ is the human-to-agent ratio of each agent, *γ*_*j*_, in that particular location, *i*. This correction ensures quantities are independent on the chosen number of agents of a given simulation (see S0.7 - Submodels). The first parenthesis on the right hand side is the density of susceptible vectors searching for a blood meal, which is then multiplied by the success fraction, or the probability of host to vector transmission integrated across all infective agents (*γ*) in a given grid cell, *n*_*i*_.

The total vector density from Eq(7) thus refers to

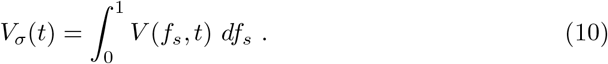

A major aspect of VECTRI is its ability to capture vector breeding habitats of diverse nature, such as temporary rain-fed or year-round ponds, the latter formed by permanent hydrological features such as rivers or lakes. The availability of breeding sites is modeled via a logistic function affecting the crowding decay rate of the larval density in Eq (6), *δ*_*c*_. For further details we refer the reader to Model calibration.

For clarity, the VECTRI model is summarized in the schematics of Fig 2. Decay rates and advection schemes are vector dependent, *e*.*g*., [32, 56]. For an explicit development of these schemes we refer the reader to [32, 56–58]. In this work we used the standard parameter setting for *An. gambiae*.

#### Agent-vector coupling

Both vector-to-human, *P*_*V* →*h*_, and human-to-vector, *P*_*h*→*V*_, transmission probabilities are the cornerstones of agent-vector interactions, and are fully developed in the S0. Agent-Based Model supplementary section. Briefly, agent-vector interactions are modeled through the human biting rate (*hbr* ), which relies on the vector-to-host ratio. In the VECTRI-ABM framework the *hbr* contains an agent-specific component and one common to all agents in a given spatial location. The latter reads as

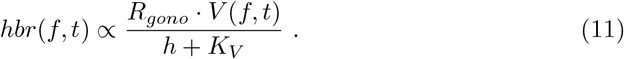

Here *R*_*gono*_ weights the vector density to obtain the blood-seeking proportion, *h* is the human population density, and *K*_*V*_ is a vector-specific parameter describing its diffusive properties. High human population densities will then act to dilute agent-vector interactions, and low densities (*h* ≪ *K*_*V*_ ) will yield a vector-dominated *hbr*. In a context where vector densities are driven by rainfall this will lead to fluctuations in the *hbr* typical of rainfall variability.

### Model calibration

To ensure the new framework accurately captures key entomological and epidemiological features of malaria transmission we performed a constrained multi-objective calibration of the model parameters using the open source Optuna [59] optimization suite. The calibration was executed in two successive stages, first focusing on parameters controlling the ecology of the vector of interest, *An. gambiae*, to then deal with parameters controlling the dynamics of the disease, *i*.*e*., immunity dynamics and vector-human interactions (S1.b Model parameters). Further details can be found in S5. Calibration.

The first fitting round (*Fit 1* ) used the multi-year monthly mean of daily *hbr* reported in Dielmo, western Senegal [60] in order to calibrate model parameters controlling vector density and seasonality. The second (*Fit 2* ) used the multi-year monthly mean of daily EIRs and the age-structured malaria incidence for the 1990–2003 period in the same study site. In addition, we used annual *Plasmodium falciparum* Entomological Inoculation Rates (A*Pf* EIRs) of the Ndiop neighboring village, where no permanent water bodies are found and incidence is highly rainfall-driven [8] (Fig S8). Dielmo is located in a marshy area in the proximity of the Nema river, thus facing year-round activity of *Anopheles funestus* accompanied by *An. gambiae s. l*. (*sensu lato*), which is most abundant during the rainy season. The flexibility of the VECTRI model to capture breeding sites with diverse characteristics allows us to use this study site for calibration purposes. As malaria incidence data results from the combined activity of both *An. gambiae s*.*l*. and *An. funestus*, the EIR and *hbr* data used for calibration was the sum of both vector species. This allowed us to calibrate rain-fed breeding sites in a scenario where *An. funestus* acted as a confounding factor. Disentangling both contributions was possible due to the year-round activity of *An. funestus*, which yields a clear signal outside the rainy season.

In both fitting rounds parameter values were sampled and selected according to their impact in minimizing the *RMSE* (Root Mean Square Error) between modeled and reported data. Further details on the calibration process and results can be found in S5. Calibration and S5.1 - Calibration tables.

### Model evaluation

#### Vector-human interactions

The model is evaluated spatially comparing simulated A*Pf* EIR against reported values for locations with different human population densities. In this way, we evaluate whether Eq (16) is a good approximation to interactions between vectors and human hosts. Studies reporting A*Pf* EIR values have been selected only when the main local vector was *An. gambiae s*.*l*. and there were no major interventions in place. These are: Barkedji [61], Ndiop, digitised from [8], Diohine [62], Kotior [62], Ngayokheme [62] and Takeme & Ousseuk [61].

#### Spatial prevalence

Additionally, since observed malaria endemicity data are spatially scarce for the study period (1990–2003) over Senegal and The Gambia, the spatial evaluation of the modeling framework was complemented by a comparison against the Malaria Atlas Project (MAP) parasite rate product (*Pf* PR_2–10_ map) [38]. It provides annual mean gridded estimates of the proportion of children aged 2–10 carrying microscopy-detectable *Plasmodium falciparum* parasites, derived from a Bayesian model driven by a set of environmental and socioeconomic factors and calibrated with scalar (point) prevalence data available over the continent. Rasters for the years 2000–2003 — the earliest available estimates overlapping with the simulated window — were accessed via the *malariaAtlas* R package [63] (release 202206) and averaged over those four years. Since the MAP grid is finer than the model grid, rasters were re-gridded by area averaging — each model cell takes the mean of all MAP pixels falling inside it. Results were aggregated to the first administrative level for spatial comparison purposes — 14 Senegalese and 6 Gambian regions. Given that MAP products report microscopy-detectable parasitaemia whereas VECTRI-ABM reports the total prevalence, including sub-patent infections, model values are rescaled for comparison by a spatially uniform detection factor, the least-squares optimum

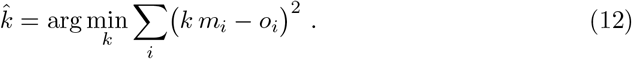

Here *m*_*i*_ and *o*_*i*_ stand for modeled and observed values in grid cell *i*, the sum running over cells where both are defined. The factor is fitted on the model grid before aggregation.

#### Seasonal cycle

The simulated seasonal cycle of malaria fever episodes was evaluated against monthly confirmed malaria cases from the official surveillance system of Senegal’s National Malaria Control Programme (*Programme national de lutte contre le paludisme, PNLP*). The control simulation covers the pre-intervention period 1990–2003, whereas surveillance covers 2009–2025. Since these periods do not overlap and correspond to substantially different levels of malaria control, the evaluation focuses on the timing and shape of the seasonal cycle rather than on absolute case numbers. For each year, monthly values were divided by the maximum monthly value within that year. The median seasonal cycle and the 10th–90th percentile range were then calculated across years for the simulation and surveillance data separately. From these median cycles we derived three summary metrics for the national domain and the northern (≥ 14.5^◦^N) and southern (< 14.5^◦^N) zones: the peak month, the full width at half maximum (FWHM) of the normalized cycle, and the Pearson correlation between the twelve monthly median values of the simulated and observed cycles, computed both month-by-month. The correlation coefficient was calculated twice: first with the raw normalized cycles and then after shifting the simulated cycle by one month. The latter was added *a posteriori* to account for the observed systematic phase lag between both seasonal cycles.

### Numerical experiments

In order to characterize immunity-driven biases in simulated malaria prevalence we performed two numerical experiments (E1,2), constituted by a total of four simulations. Model parameters controlling permanent water bodies, essential to capture the presence of *An. funestus* in Dielmo, were set to zero, as experiments focused on the rain-driven ecology of *An. gambiae* at regional scales. The population size was set to 4 million agents, as lower sizes showed a qualitative difference in the dynamics. This choice implied human-to-agent ratios that ranged from 1–5, either limit representing scarcely populated rural areas and dense urban regions, respectively (see S0.5 - Initialization). **E1: Main**. The first, *control* simulation, was run from 1990 to 2003. We chose 1990–2003 as this represents the period before interventions scale up in many African countries, including Senegal [64]. Model diagnostics were then aggregated into age classes, including symptomatic cases and agent-dependent immunity dynamics, *e*.*g*., 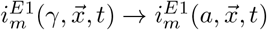. We shall address this simulation as *fine-scale*. The second, *counterfactual* simulation, was run analogously to the first, with the difference lying in the population immunity. Instead of allowing immunity to freely develop at the agent level, we set it to the *control* ‘s population average. This is, for all grid cells at each time, 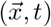, we calculated the average immunity across all ages, 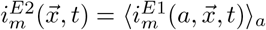, and assigned it to all agents in that particular cell. With 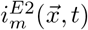 acting as an external forcing, we simulated a climate-sensitive evolution of the immunity level, which is however shared by the entire population, and thus neglects cumulative history of exposure. We shall address this simulation as *pop-average*. The analysis then focused on characterizing age-stratified differences in symptomatic (*I*) and asymptomatic (*A*) prevalence from both simulations, driven by their different immunity distributions. **E2: Maternal immunity**. We performed a second pair of factual-counterfactual simulations where maternal immunity was removed in both settings. This experiment allowed to separate this protection mechanism from other confounding factors simultaneously contributing to observed biases in E1, and particularly affecting agents during their early life stages (∼ 0–2 years of age).

#### Input Driving Data

The VECTRI-ABM framework uses the human population density and age structure of the simulated region to drive the agent module. The population density is used to allocate agents spatially and the age structure to generate the age distribution and use it to first assign initial ages to agents, and then to derive growth and mortality rates from the exponential fit to this age distribution. Further details are provided in S0.6 - Input data. Daily air temperature at two meter height, *T*_2*m*_ (^*o*^*C*), and rainfall, *R*_*d*_ (*mm/day*) are used to drive the vector dynamics. For the temperature and rainfall data we used the Climate Hazards Center InfraRed Temperature with Stations (CHIRTS) [65] and Climate Hazards InfraRed Precipitation with Stations (CHIRPS) [66] datasets, available at 0.05^*o*^ spatial resolution for the period 1990–2003. Demographic information was obtained from the WorldPop database [67] using the year 2000 (see Results - Fig 3). For simulation purposes, the age structure is interpolated, as the WorldPop’s estimates aggregate reports in 5 year intervals (see S3. Age methods - FigS6), while the VECTRI-ABM framework considers age at a daily resolution. Age data was interpolated to 1 year intervals, which were then used to initialize the agent’s age profile, assuming a uniform distribution of ages within each year (see S0.5 - Initialization).

**Fig 3.**
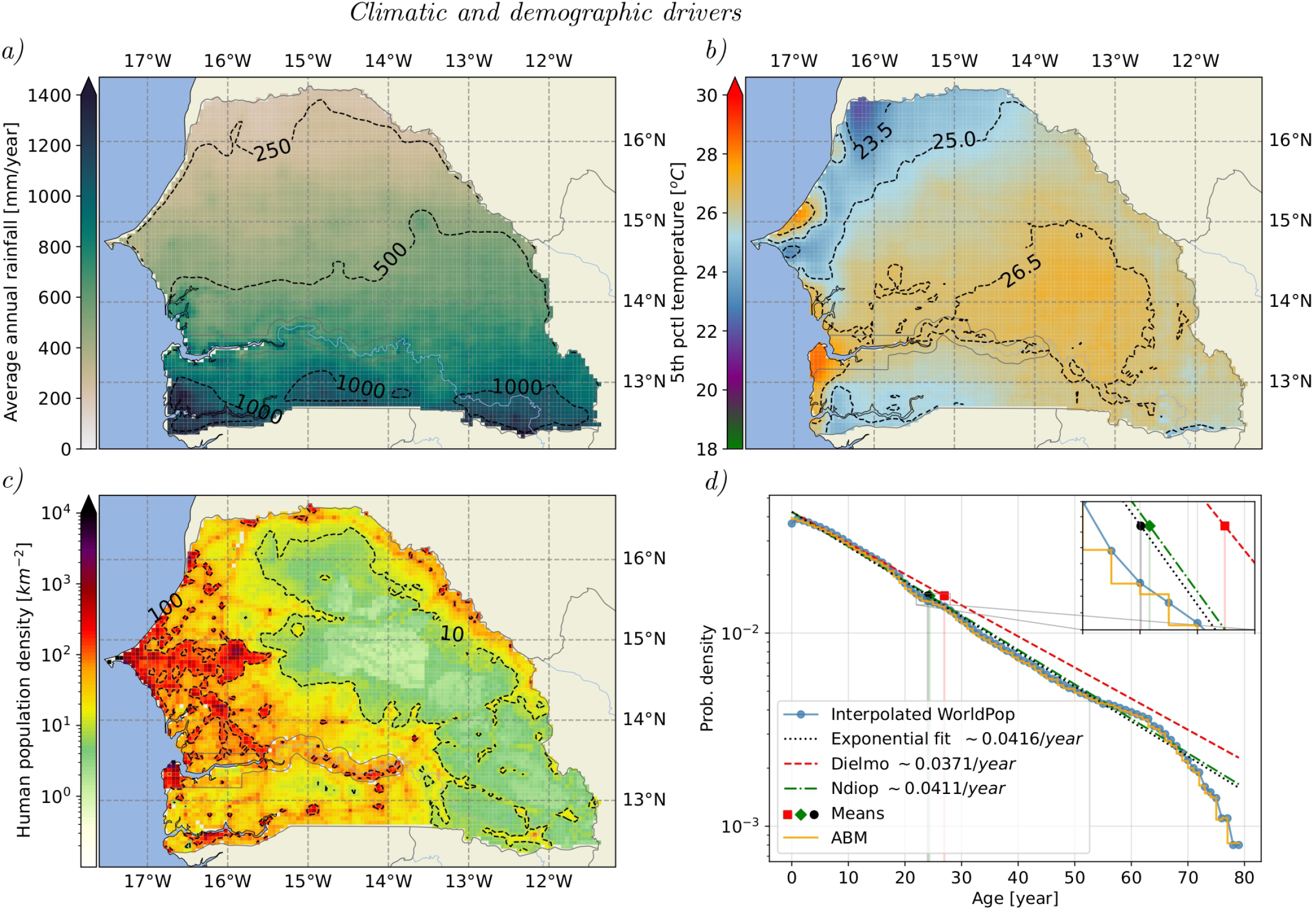
**a)** Annual rainfall 1990–2003 average. **b)** 5*th* percentile CHIRTS temperature in the 1990–2003 period **c)** Human population density as of 2000. **d)** Age distribution density for Senegal and The Gambia, as derived from the WorldPop’s, and compared to the ABM’s and those locally estimated for Dielmo and Ndiop [8].

**Fig 4.**
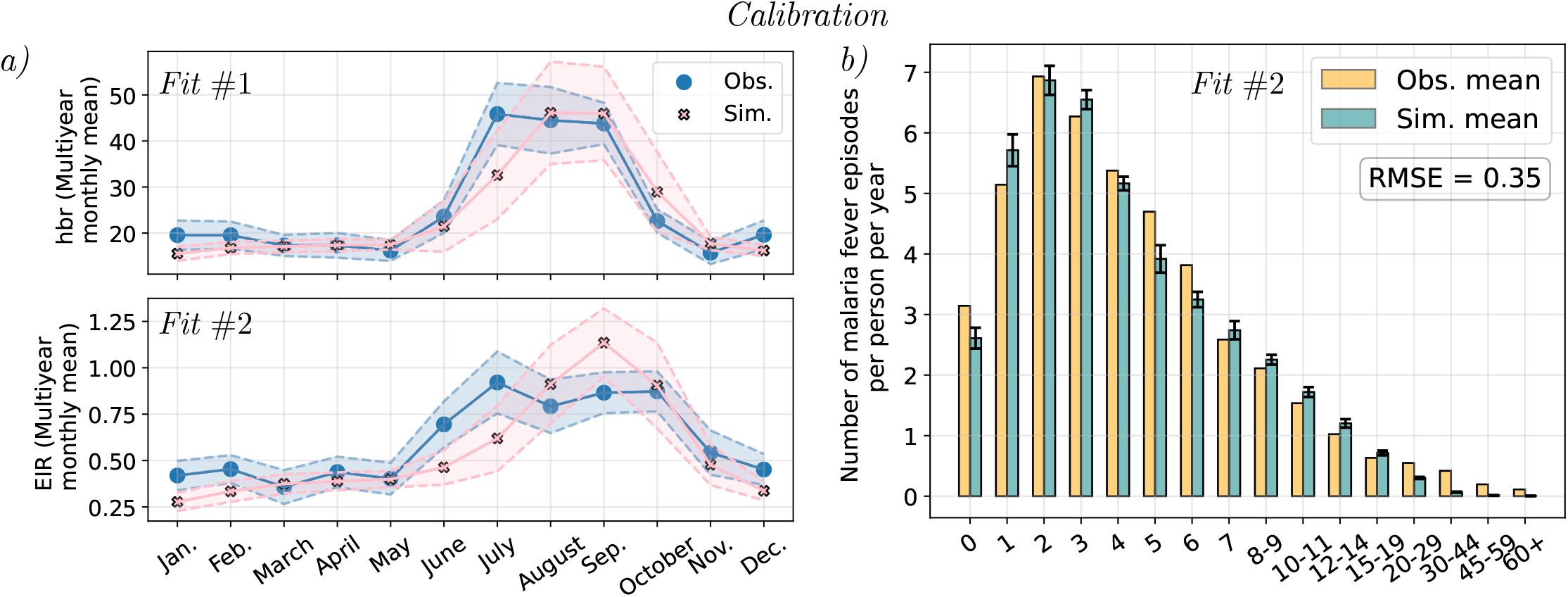
**a)** Simulated and observed multi-year monthly mean daily *hbr* and *Plasmodium falciparum* Entomological Innoculation Rate (*Pf* EIR). Averages are depicted by solid lines, while the mean *±*1 standard deviation is represented by envelopes, for the period 1990–2003 in Dielmo. **b)** Simulated and observed age-structured malaria incidence during the period 1990–2003 in Dielmo.

## Results

### Climatic and demographic drivers in Senegal and The Gambia

The region under scope shows a pronounced latitudinal rainfall gradient, anticipating differences in the distribution of *An. gambiae* populations and, consequently, in *P. falciparum* transmission (Fig 3a). The rainy season in Senegal and The Gambia is unimodal, approximately occurring from June to October. Annual minimum temperatures always remain above the threshold for sporogonic development (∼ 18^*o*^*C*), with no single day being below during the study period (Fig 3b). Temperature is thus a secondary driver that only limits vector populations in the eastern-most areas of the region, where temperatures exceed 36 ^*o*^*C* during short periods of time, via changes in mortality, constituting a hard constraint for the presence of the vector or the parasite (Fig S7), contrary to rainfall. Human population density values are larger in the west (Dakar region in Senegal and Kanifing and Brikama in The Gambia) and south-west (Casamance) part of Senegal, with some high-density regions found as well in the north-west (Saint-Louis) and the north-eastern Maurinatia-Senegal border (Matam region) (Fig 3c). The eastern part of the region is, however, scarcely populated, including northern areas with low levels of annual rainfall. The country’s age structure is exponentially distributed (Fig 3d) with a mean below 30 years of age. From the fit to the distribution we obtained an agent’s mortality rate of *µ*_*d*_ = 0.0416*/year* and assigned ages across agents appropriately in the numerical experiments (Fig 3d).

### Model calibration

#### Calibration: *Fit 1*

Reported *hbr* values in Dielmo show a marked transition between year-round low values during the dry season, followed by a sharp increase in the rainy season, from June to October and peaking in July. The first fitting round yielded modeled mean monthly *hbr* values that accurately capture reported observations (Fig 5a, top). A systematic discrepancy is, however, observed in the month of July, where simulated values are persistently lower than observations. Monthly standard deviations are of similar magnitude as well, both increasing during the rainy season, as expected from the rainfall’s inter-annual variability. Throughout the year *hbr* levels remain finite, given the village’s proximity to the Nema river, which sources permanent, year-round breeding sites exploited by *An. funestus*.

**Fig 5.**
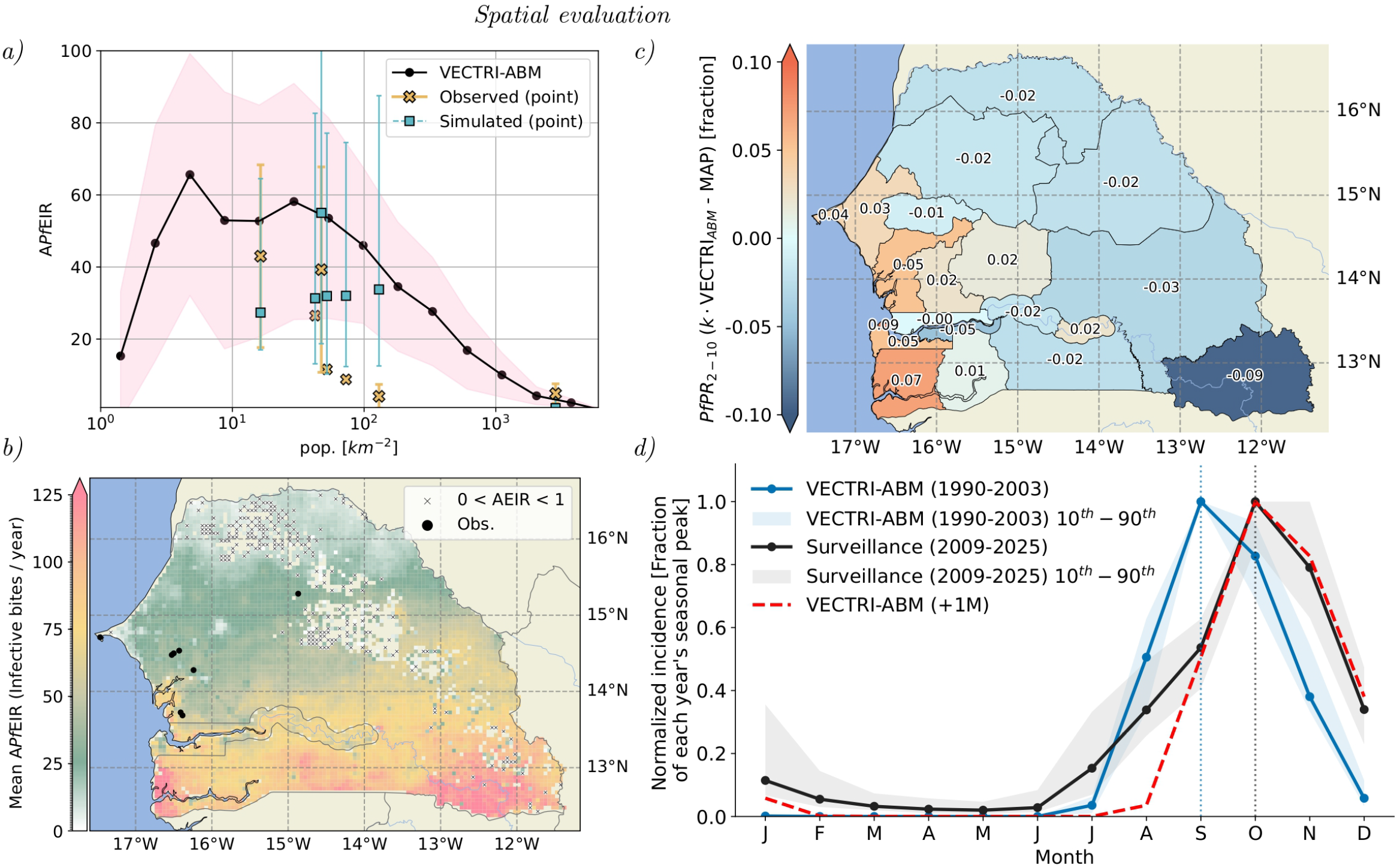
**a)** Spatial mean (black dots) and standard deviation (shading) of the time-averaged A*Pf* EIR as a function of the human population density. Crosses point single location observations and squares the corresponding same-location simulated values, accompanied by their 10*th* and 90th time percentiles. When reported, observations include the standard deviation. **b)** Simulated spatial distribution of A*Pf* EIR for the period 1990–2003 and spatial location of observations reported in a). **c)**District-mean *PfPR*_2−10_ difference,VECTRI-ABM minus MAP estimates, over 2000–2003, for the 14 Senegalese and 6 Gambian districts. Labels given differences as a prevalence fraction.**d)** Normalized seasonal cycle of malaria incidence: simulated monthly malaria fever episodes per person (VECTRI-ABM, 1990–2003) compared with confirmed malaria cases from the *PNLP* surveillance system (2009–2025). For each year, monthly values are divided by that year’s maximum; solid lines show the across-year median and shaded envelopes the 10th–90th percentile range. Model and surveillance periods do not overlap, so the comparison targets the timing and shape of the cycle rather than absolute case numbers.

#### Calibration: *Fit 2*

Similarly, reported EIR values follow a dry-wet transition, peaking in July, yet remaining high until October. The modeled monthly mean EIR values represent the transmission seasonality, with small biases towards lower values and the same discrepancy observed in the *hbr* calibration, this time extended to the month of June as well (Fig 5a, bottom). The model systematically overestimates EIR in September. The age-structured malaria incidence qualitatively and quantitatively captures the reported shape, by which incidence peaks early in a subject’s life to then decrease as a result of age and acquired immunity (Fig 5b). At the time of report, Dielmo was a holoendemic village, with constant transmission pressure that drove the early development of immunity against malaria. This is reflected in the early incidence peak, around 2 years of age, and the subsequent fast decrease, reaching an average of less than 1 episode per person per year a the age of 12. Across all age bands, the modeled *vs* reported RMSE in the number of malaria fever episodes is smaller than 1*/*2 case per person per year.

The second, multi-objective fitting round produced multiple parameter combinations that were considered of equivalent quality, this is, all belonging to the *Pareto* front. The final parameter combination, used subsequently in the numerical experiments, was manually selected according to visual assessment of the reproduced seasonality in the EIR and the simulated age-structured malaria incidence.

### Model evaluation

#### Vector-human interactions

At high human population densities (∼ 10^3^ *km*^−2^) the modeled A*Pf* EIRs are dominated by a strong dilution effect that progressively fades, allowing inoculation rates to increase at lower population density values (∼ 10^3^–10^2^ *km*^−2^) until a plateau is reached (10^1^ *km*^−2^, Fig 5a). This mean increase is accompanied by that of its standard deviation, marking a transition to rainfall-dominated dynamics, where the local human population density is no longer the main driver modulating transmission. As human population densities continue to decrease transmission becomes harder to sustain, with decreasing A*Pf* EIRs from ∼ 10^1^ [*km*^−2^] onward, until transmission is lost. Empirical observations of A*Pf* EIR during the simulation period (1990–2003) show a similar level of fluctuations and a similar transition from population density- to climate-dominated A*Pf* EIRs. Some observations lie below the modeled mean, with Ngayokheme and Takeme & Ousseuk being outside the range of typical simulated inter-annual fluctuations, thus pointing towards a modeling overestimation.

#### Spatial A*Pf* EIR & prevalence

Spatially, malaria transmission is a composite of the rainfall and human density maps (Fig 5b). A*Pf* EIRs are higher in wetter regions with intermediate human density values. These are predominantly located in the south, with large A*Pf* EIR values simulated over the south-eastern and south-western regions of Senegal, given that population densities are relatively lower. Simulated A*Pf* EIR maxima correspond to the Kédougou and Ziguinchor regions, historical hotspots of malaria in Senegal [68]. Moving northward along the rainfall gradient, transmission dynamics shift alongside human population density. Moderate A*Pf* EIRs in the west drop to no or barely-sustained levels in the central (∼ 15^*o*^*W* –14^*o*^*W* ) and northern regions, followed by a recovery in the east.

Differences between VECTRI-ABM and MAP *Pf* PR_2–10_ estimates, before correcting with the detection factor, *k*, are positive in all 20 administrative units, by 6–25 percentage points (Fig S9). This is an expected result when a total prevalence estimate is compared against a microscopy-detectable one. By applying the detection factor (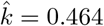, fitted over *n* = 6814 cells) the offset is removed, with the mean difference falling to 0.6 percentage points and the RMSE from 17 to 4.2 (Fig 5c). Differences are not spatially uniform, with a correlation of *r* = 0.25 (*p* = 0.29, *n* = 20 — administrative units), unchanged by the uniform re-scaling of the detection factor. We find a west-to-east bias between models, with larger discrepancies in coastal areas — most markedly in the Ziguinchor region (+7 percentage points), where the MAP product has the third-lowest *Pf* PR_2–10_ in the domain (0.085, not shown) — and in the south-eastern Kédougou region (−9.4), the highest in the MAP *Pf* PR_2–10_ product (0.227, not shown). **Seasonal cycle**. The model predicts a ∼ 2-month delay between both the onset and offset of clinical malaria incidence with respect to rainfall (Fig S10). It, however, shows a shorter ∼ 1 − 2-month delay between their respective peaks. Hovmöller diagrams illustrate these delays (Fig 6), broken down to key stages in the climatic and transmission cycles. The rainy season starts in the south in May, peaking in August and lasting until late October, with a gradual northward spread and a sharp retreat (Fig 6, top left). This is closely followed by vector proliferation from June to November (Fig 6, bottom left), with a subsequent a rise in EIR and incidence (Fig 6, top and bottom right, respectively), which peak in September. The malaria season appears symmetric in time, with a minor tail in northern regions (> 14.5^*o*^) and longer seasons in the south.

**Fig 6.**
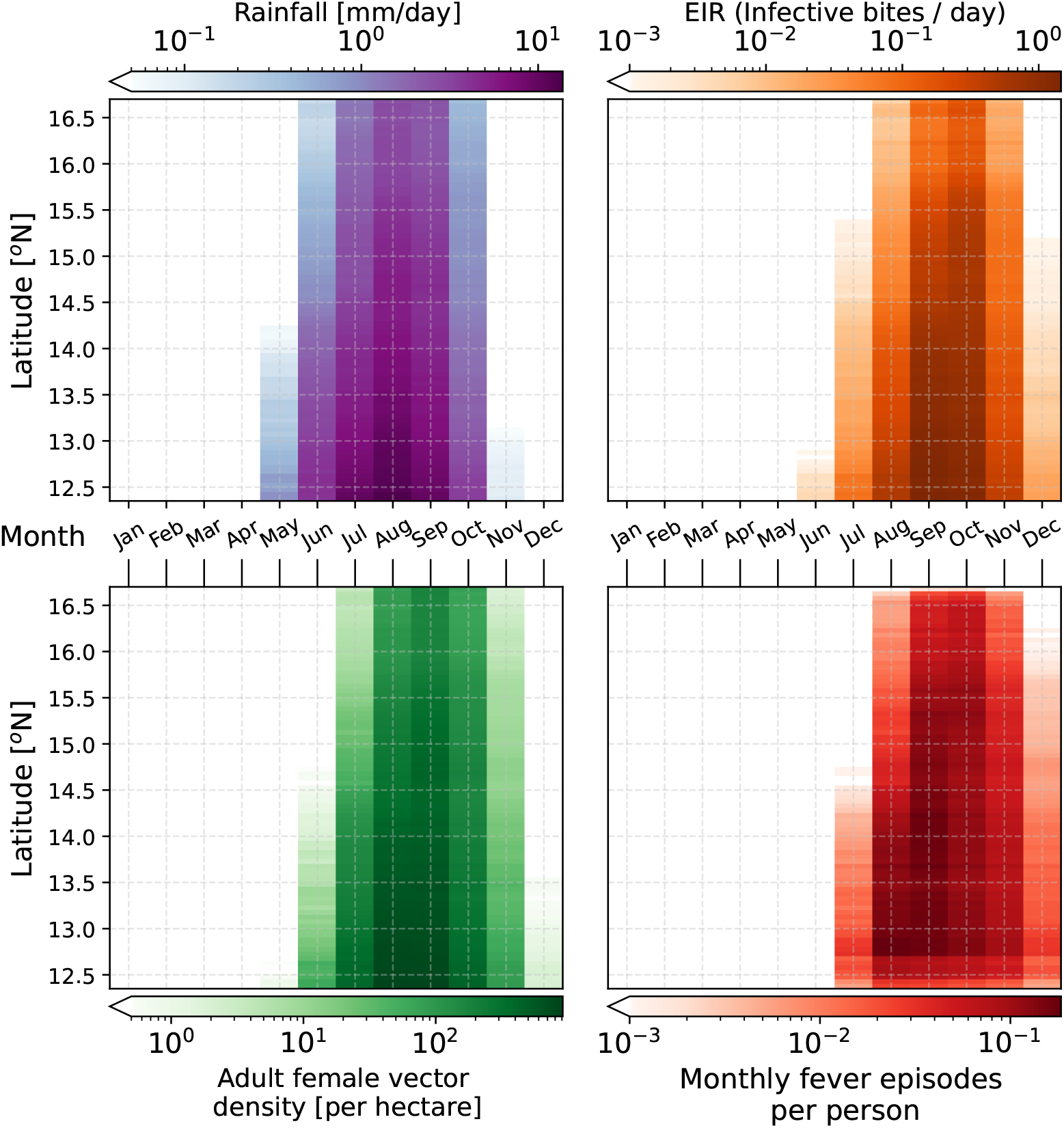
Hovmöller diagrams of daily rainfall, female adult vector density, daily EIR and monthly malaria fever episodes per person. These are the climatological means for each calendar month, at each latitude (zonal mean). The climatological averages are computed over the duration of the experiment (1990–2003).

Both the model and the *PNLP* surveillance data describe a single, strongly seasonal cycle, with simulated malaria fever episodes respectively peaking in September and October, *i*.*e*. we observe a one-month phase lag of the observations relative to the model (5d). Despite this shift, the overall width of the median seasonal cycle is similar: the full width at half maximum (FWHM) is 2.74 months in the model versus 2.83 months in the surveillance data at the national level (Table 1). The Pearson correlation between the twelve monthly median values is 0.806 for the raw seasonal cycles and increases to 0.967 once the simulated cycle is shifted by one month, indicating that temporal alignment accounts for a substantial part of the discrepancy, while differences in the seasonal tails remain (Table 1). Similar timing and width patterns are observed when the country is split at 14.5^◦^N into a drier northern and a wetter southern zone (Fig S11 and Table 1): in both zones the model peaks in September and the surveillance in October, with FWHM values of 2.34 (model) versus 2.53 months (observed) in the north and 3.18 versus 3.04 months in the south, and one-month-shifted correlations of 0.961 and 0.911, respectively. Qualitatively, the main residual discrepancy concerns the tails of the seasonal cycle: the surveillance data show a more gradual early-season build-up and a more persistent late-season tail than the model, particularly in the northern zone. This descriptive comparison does not identify the mechanism responsible for these differences. The interpretation of the timing and tail differences is developed in the Discussion.

**Table 1.** Summary statistics of the normalized malaria seasonality comparison.

| Zone | Peak Month |  | FWHM [months] |  | Correlation (Pearson) |  |
| --- | --- | --- | --- | --- | --- | --- |
|  | Model | Observed | Model | Observed | Same month | Shifted (+1M) |
| National | September | October | 2.745 | 2.828 | 0.806 | 0.967 |
| North ($\geq 14.5^\circ$) | September | October | 2.344 | 2.528 | 0.753 | 0.961 |
| South ($< 14.5^\circ$) | September | October | 3.179 | 3.038 | 0.770 | 0.911 |

Summary statistics of seasonality comparison between VECTRI-ABM (14 years) and surveillance data (17 years). FWHM: Full Width at Half Maximum. The Pearson correlation is computed between the twelve monthly median values, both for the raw seasonal cycles (“Same month”) and after shifting the simulated cycle by one month (“Shifted +1M”).

### Comparison of numerical experiments

#### Immunity-driven biases in simulated malaria incidence: age structure and rainfall dependence

The first numerical experiment, E1, where we compare the *fine-scale* and *pop-average* simulations, reveals systematic differences in the time-averaged symptomatic prevalence, *Ī*, across the full age spectrum (Fig 7). In denser human population regions (*h* > 50 *km*^−2^), where demographic stochasticity plays a secondary role, simulated daily prevalence biases range from 0% to 20%. The magnitude of these differences, Δ*Ī* ≡ *Ī*_fs_ − *Ī*_pa_ (fs: *fine-scale*, pa: *pop-average*), is strongly modulated by mean rainfall. Wetter locations, characterized by higher entomological inoculation rates (EIRs), present larger biases across the 0–11 year age classes, with the 1–5 and 8–9 year age classes being the most misrepresented by the *pop-average* model in dry and wet rainfall regimes, respectively (see golden dots in 7). Differences increase with age, until an inflection point is reached and these start to decrease. The 15–19 year age class constitutes a pivotal point where both the bias and its sensitivity to rainfall are approximately zero, thus marking the limit where age classes would be “well represented” by the *pop-average* model for any mean rainfall value.

**Fig 7.**
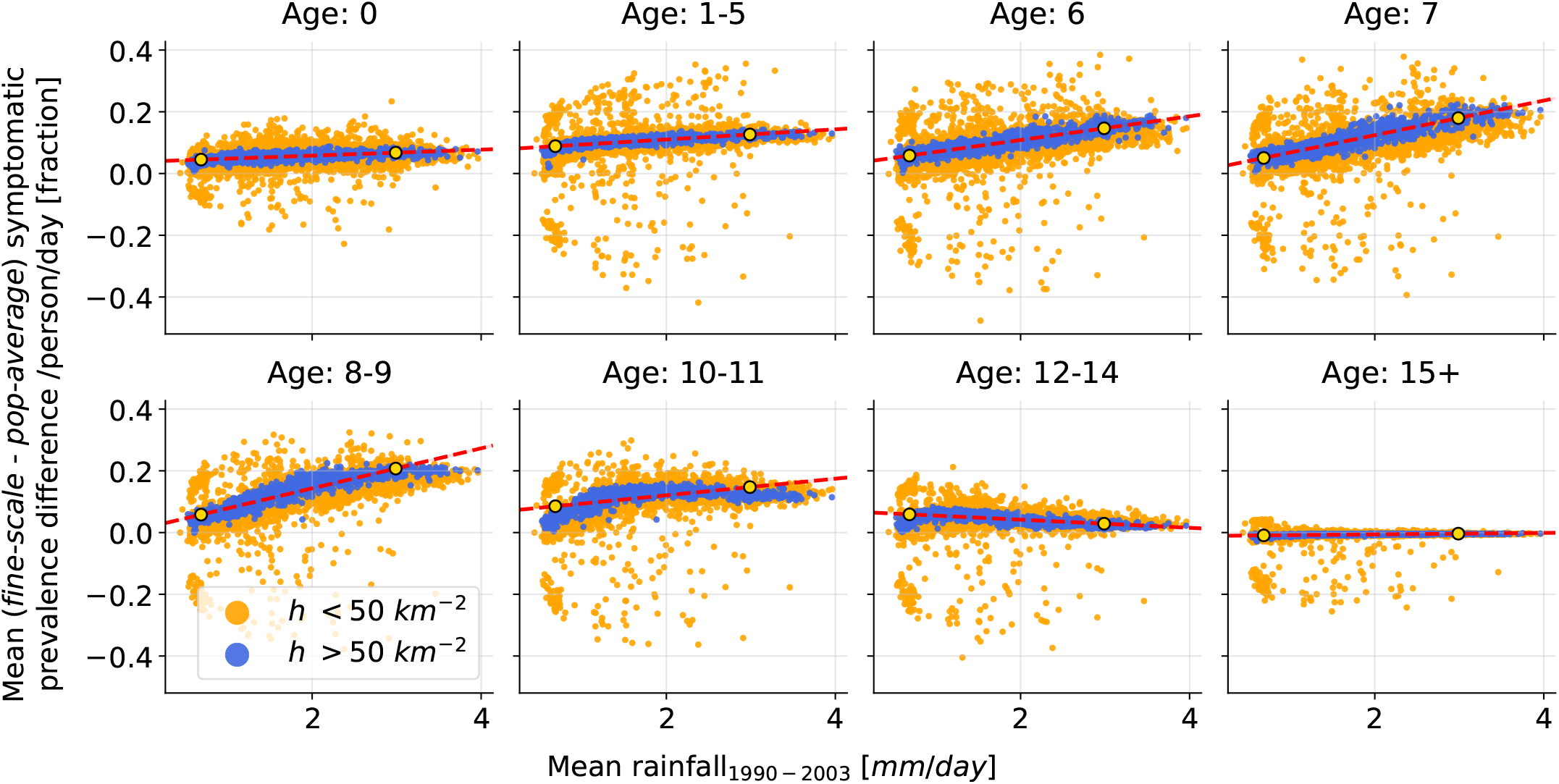
Age-disaggregated malaria symptomatic prevalence differences between the *fine-scale* and *pop-average* simulations against the mean daily rainfall during the simulated period (1990–2003). In orange we find human population densities below 50 *km*^−2^, with blue marking those above. Analysis excludes locations with average *APf EIR <* 10^−2^. Dashed red lines and golden dots providing three summary statistics of the calculated biases: the bias at low mean daily rainfall values, for which we choose the 5^*th*^ percentile, as there are no data with zero rainfall ; the high rainfall bias, for which we choose the 95^*th*^ percentile ; and the slope of the data points for each age class. These statistics are calculated from a linear regression of these data using human population densities *h >* 50 *km*^−2^.

#### Mechanisms of demographic heterogeneity

In order to understand the mechanisms behind these biases we decompose the mean simulated prevalence (E1) difference into its symptomatic, Δ*Ī*, and asymptomatic, Δ*Ā*, components (Fig 8a, bottom panel), and analyze their spatial means, ⟨Δ*Ī*⟩_*x*_ and ⟨Δ*Ā*⟩_*x*_, in terms of the factors that modulate incidence via acquired immunity, *i*_*m*_. These are symptomatic probability curves (Eq 1 and Fig 8b) and clearance times (Eqs 2-4). E2 was used here to isolate the contribution of maternal immunity, which acts as a confounding factor. We identify four distinct regimes (R) across the age spectrum, each dominated by a different combination of mechanisms.

**Fig 8.**
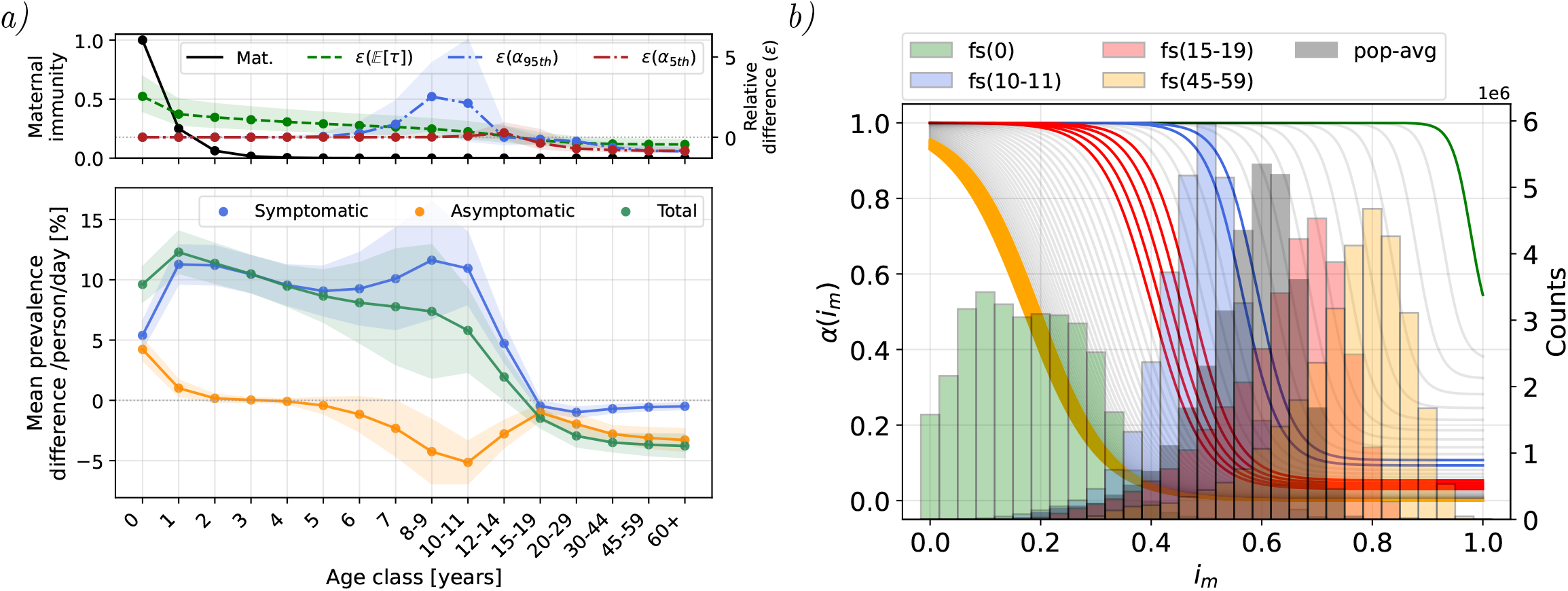
**a)** Bottom. Symptomatic, asymptomatic and total mean prevalence bias (fs-pa) per age class. Top. Maternal immunity decay function and relative differences, *ϵ*(*x*) = (*x*_fs_ − *x*_pa_)*/x*_pa_ between expected clearance time, E[*τ*], and symptomatic probability, *α*, the latter chosen at the 5*th* and 95*th* mean rainfall percentile locations. Prevalence and clearance time differences (top and bottom panels) are shown using the spatial mean (dots) and standard deviation (envelopes) of the temporal mean in the domain, thus providing an assessment of rainfall-driven variability across the grid. The relative difference in symptomatic probability shows however the local time median, enveloped by the 10*th* and 90*th* time percentiles, thus quantifying temporal variability in that particular location. **b)**. Immunity histograms for distinct *fine-scale* age classes (colors) and the *pop-average* case (gray) accompanied by the age-dependent sigmoidal scheme of symptomatic probability, *α*(*i*_*m*_; *a*). Colors match same-age classes.

#### R1. Ages 0-1: clearance times and maternal immunity as a router

At age 0, when most agents’ maternal immunity is active, both ⟨Δ*Ī*⟩_*x*_ and ⟨Δ*Ā*⟩_*x*_ are positive and of similar magnitude (∼ +5%, Fig 8a). As at this age symptomatic probabilities between *fine-scale* and *pop-average* populations are the same (Fig 8b) the total prevalence bias, Δ*P* ≡ ⟨Δ*Ī*⟩_*x*_ + ⟨Δ*Ā*⟩_*x*_, reflects here differences in clearance times: agents in the *fine-scale* simulation carry low immunity (Fig 8b), which results in longer duration of infections, leading to a higher prevalence. Maternal immunity acts in this regime as a router rather than a source of excess prevalence - while active it unconditionally forces agents into an asymptomatic state upon exposure, partially suppressing ⟨Δ*Ī*⟩_*x*_ (E2, Fig S12 in S4. Supplementary plots.). Lower total prevalence values at age 0, with respect to age 1, correspond to the buffering effect of susceptible (*S*) newly born infants, which take at least *IIP* = 10 days to be able to contribute into the prevalence pool.

#### R2. Ages ∼ 1-4: maternal immunity waning and equal symptomatic probabilities

As maternal immunity wanes (half-life ∼ 6 months) most infections become symptomatic, yet the excess prevalence, Δ*P*, decreases, driven by the decrease in average clearance times (Fig 8a, top), relative to the *pop-average* population, as calculated from

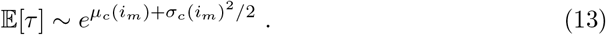

#### R3a. Ages ∼ 4-19: crossing the pyrogenic threshold

As we focus on older agents, these will experience different responses against the disease, as determined by the age-dependent factors of immunity dynamics. Both *pop-average* and *fine-scale* agents will cross the pyrogenic threshold sequentially, ordered by rainfall intensity. The shift in pyrogenic threshold with age (Eq (1) and Fig 8b) will affect agents in the *pop-average* simulation first, as these continue to have higher immunization (Fig S13 in S4. Supplementary plots.). Crossing the threshold in advance is reflected in the increase (decrease) of ⟨Δ*Ī*⟩_*x*_ (⟨Δ*Ā*⟩_*x*_), as *fine-scale* agents continue to be in the upper plateau part of the sigmoidal function controlling symptomatic (*I*) probabilities. Regions with high mean rainfall values, and therefore higher population-average immunity, will experience the transition at younger ages (ages 4–11), with wettest areas showing up to 5-fold differences in symptomatic probability (*ϵ*(*α*_95*th*_) in Fig 8a, top). Drier regions, on the other hand, lag behind, experiencing the transition during late adolescence years (ages 15-19), as these acquire immunity at a slower pace, crossing the pyrogenic threshold synchronously with the *fine-scale* agents, and thus experiencing lower relative differences in excess prevalence (Fig 8a, top and Fig S13 in S4. Supplementary plots.). The spatial variance of this crossover age, visible in the standard deviation bands of Fig 8a (bottom) and in the rainfall-driven curves of Fig 7, reflects the transmission intensity gradient across the study region: in drier, low-EIR locations immunity accumulates more slowly and the crossover shifts to older age classes, while wetter locations converge earlier. This rainfall-dependent crossing is the primary driver of the observed positive ⟨Δ*Ī*⟩_*x*_ and negative ⟨Δ*Ā*⟩_*x*_, and explains why the bias magnitude (Fig 7) scales with mean rainfall.

#### R3b. Ages ∼ 15–19: population convergence

Immunologically, agents reach adulthood around the 15–19 year age class, when total excess prevalence decays rapidly to zero for all climatic regimes (Fig 7 and Fig 8a, bottom).

#### R4. Ages >20: under-immunized adults in the *pop-average* population

Beyond the 15–19 convergence, ⟨Δ*Ī*⟩_*x*_ remains near zero, while ⟨Δ*Ā*⟩_*x*_ continues to decrease with age, becoming the greatest contribution to the total bias in older age classes (Fig 8a). In this regime, the relative magnitudes of immunity in the two populations are reversed with respect to the younger age classes: since the *pop-average* immunity is taken across all ages, it is pulled down by younger, less immune agents, thus falling below that which *fine-scale* agents have accumulated by adulthood (Fig 8b). Adults in the *pop-average* population are thus under-immunized, clearing infections more slowly, which results in higher asymptomatic prevalence than in the *fine-scale* simulation, yielding ⟨Δ*Ā*⟩_*x*_ < 0. The fact that ⟨Δ*Ā*⟩_*x*_ dominates this deficit is due to the high immunity of both populations at these ages, which saturates the symptomatic probability at the lower plateau of the sigmoidal curve, *α*_min_(*a*) (Fig 8b). The low-immunity tail of the *pop-average* immunity distribution, which remains in the finite symptomatic probability regime, generates the residual ⟨Δ*Ī*⟩_*x*_ *<* 0 values.

## Discussion

### Objectives & Results

In this study we have determined how demographic heterogeneity in immunity against malaria shapes community-level prevalence, characterizing the age strata for which the bias is highest as a function of rainfall conditions. This study compares two modeling approaches to immunity: the first accounting for the individual’s history of exposure to the disease (*fine-scale*); the second assuming that all individuals in the population share the same level of immunity (*pop-average*), thus mimicking the underlying assumptions of standard compartmental models. The differences between the two simulations, or the bias (Δ ≡ *fine-scale*−*pop-average*), then show how the two models allocate burden differently, thus indicating the limitations of the standard *pop-average* approach. During the first 1–2 years of life, when maternal immunity is still active (half-life ∼ 6 months), both symptomatic and asymptomatic prevalence biases are positive (Δ > 0) and of similar magnitude (spatial mean ≈ 5 percentage points of daily prevalence). The two components separate in populations of 3–5 years of age: the symptomatic bias remains positive and, after a small decay, grows — especially so in the wettest regions, with higher EIRs; the asymptomatic bias, in contrast, becomes zero, changing sign thereafter. Symptomatic and asymptomatic biases are maximal near the age where *pop-average* individuals cross the pyrogenic threshold (spatial means ≈ 11 and −5 points, respectively), with this crossing age being a function of local rainfall, ranging from around 4–5 years of age in high-transmission locations and extending into the 15–19 year class in the lowest transmission ones. When a certain degree of immunization has been achieved, typically in populations above 15 years of age, individual history of exposure no longer shapes symptomatic prevalence, thus becoming a secondary trait modulating clinical malaria episodes. This demographic (15+) would then be well approximated, in terms of clinical burden, by the *pop-average*’s simpler approach to malaria immunity modeling. A lower residual reservoir of asymptomatic adults is, however, predicted by the *fine-scale* modeling approach (≈ 4 points), with implications for disease transmission.

In this study, we make explicit the contrast between rainfall regimes: rainfall sets the length and intensity of the malaria transmission season, which in turn set the rate at which exposure accumulates, determining the age at which acquired immunity becomes effective. This redistribution of burden across ages is documented in field data. The study by Carneiro *et al*. performed a pooled analysis from multiple sites (*n* = 86) across sub-Saharan Africa to find that malaria burden shifts to younger age groups as transmission intensity increases, and that this effect is moderated in settings with marked seasonality [69]. The north–south gradient of the study region reproduces this behavior, with the knock-out experiment identifying acquired immunity as the mechanism responsible for the shift.

### Model evaluation & limitations

This study introduces VECTRI-ABM, a new spatially explicit, agent-based modeling framework that fully couples climate-driven vector ecology and individual-level malaria immunity dynamics. By transitioning from a compartmental to an agent-based representation of the human host, we resolve individual exposure to malaria and, consequently, obtain a granular description of acquired immunity.

This novel modeling framework was successfully calibrated against epidemiological and entomological data from Dielmo, a high-transmission perennial village in Senegal with one of the longest records on malaria incidence, stratified by age. Complementary data from Ndiop, a neighboring village with seasonal, rain-driven malaria, were used. The simulated vector dynamics reproduce the timing of the observed seasonal EIR peak, while discrepancies remain after the onset of the rainy season. We observed systematic discrepancies between observed and simulated EIRs during June and July, immediately after the onset of the rainy season. Laneri *et al*. linked early-season dynamics at Dielmo to the Nema River and permanent breeding sites used by *An. funestus* [8]. Such local hydrological processes are therefore a plausible source of discrepancy, although their contribution is not quantified here.

The regional evaluation reproduces broad spatial patterns in Senegal and The Gambia, with discrepancies following a west-to-east gradient: VECTRI-ABM simulated values exceed MAP estimates along coastal regions while falling below it inland, especially in the south-east. Several factors may contribute to such discrepancies, including human mobility, which is not represented in VECTRI-ABM. However, the comparison should be treated cautiously because MAP and VECTRI-ABM are both model-based estimates rather than direct observations. The magnitude of the fitted detection factor, 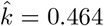, is consistent with reports on sub-patent parasite carriage: in endemic populations microscopy detects roughly half of the infections identified by PCR — 50.8% (95% CI 45.2–57.1) across 65 paired microscopy–PCR prevalence estimates [70] and 54.1% (95% ; CI 50.3–58.2) across 106 surveys [71].

### Seasonal evaluation

Because the control simulation represents the 1990–2003 pre-intervention period, whereas the *PNLP* surveillance series covers 2009–2025, this comparison should be interpreted as an evaluation of seasonal timing and shape rather than of absolute malaria burden. Despite the non-overlapping periods and substantial changes in malaria control efforts, both VECTRI-ABM and *PNLP* confirmed case data reproduce a unimodal seasonal cycle in both north (≥ 14.5^*o*^) and south (14.5^*o*^) zones, with a longer seasonal cycle in the south. The simulated incidence peak occurs, however, approximately one month earlier than the peak in confirmed cases. Simulated dynamics are consistent with previous evaluations in Senegal, which reported a rainfall maximum in August, a simulated EIR maximum in September, and a maximum of confirmed malaria cases in October [72, 73].

The one-month phase difference may partly reflect the distinction between simulated malaria fever episodes and routinely reported confirmed cases. Previous work in Senegal has attributed the lag between simulated EIR and confirmed cases to parasite development and subsequent diagnostic detection [73], of which only the latter applies to the comparison made here — VECTRI-ABM accounts for the delay between infection and symptom onset. Because care-seeking, diagnosis, and notification delays are not explicitly resolved here, their separate contributions cannot be quantified from the aggregated monthly data. Spatial aggregation of asynchronous local seasonal cycles may also affect national-scale timing.

Field evidence demonstrates substantial spatial and individual heterogeneity in exposure to *Anopheles* bites [74], whereas VECTRI-ABM assumes homogeneous mixing within each grid cell. Whether within-cell mixing assumptions contribute to the gradual *PNLP* rise and late-season tail remains untested in the present analysis.

The regional experiments set the permanent-water body contribution to zero and resolve only the rain-driven pond ecology of *An. gambiae*. Faye *et al*. [75] documented a predominant *An. funestus* population at two sites in central Senegal, sustaining residual transmission even in low-transmission conditions. This supports the biological plausibility of transmission persisting outside the main rain-driven peak, but it does not establish *An. funestus*-driven transmission as responsible for the late-season tail in the national *PNLP* series. The relevance of this mechanism depends on the vector and permanent-water configuration used in the regional simulation. Future evaluations using overlapping time periods, explicit observation-delay processes, subgrid-scale spatially heterogeneous transmission, and multiple vector ecologies would help distinguish these mechanisms.

Several simplifications may also constrain the scope of these results. The malaria module does not account for host genetic background, which may modulate host immune responses (*e*.*g*., reduced susceptibility reported in Fulani populations [76]). Furthermore, the new malaria module does not explicitly discriminate between clinical and parasite immunities, nor does it resolve clone-specific immunity acquisition. This would be tractable with a within host model of parasite development, but given the lack of solid knowledge of fundamental biological and immune mechanisms and a precise mathematical formulation to represent them [77], we opt for a more general, phenomenological parametrization of malaria dynamics in the host. Current within host models are constrained to use malaria-naive datasets [78, 79], and thus future development of a similar model in VECTRI-ABM would require further advances characterizing parasite dynamics in infected hosts from highly endemic areas.

### Model transferability

In this study, the VECTRI-ABM model has been calibrated for a high-transmission setting, typical of Senegal and The Gambia during the 1990s and early 2000s, before interventions scaled up nationally [64]. This means parameters controlling adult clinical incidence and clearance times are poorly constrained (see S5.1 - Calibration tables), since their effect is invisible in highly endemic areas. The unidentifiability of these model parameters imposes constraints on the transferability of the current VECTRI-ABM setting to modern scenarios, where transmission intensity may be one to two orders of magnitude lower. At Dielmo, one of the sites used here for calibration, the EIR fell from ∼ 482 to ∼ 7.6 infective bites per person per year between 2000 and 2012 due to intervention measures, while clinical incidence fell from 2.63 to 0.046 malaria attacks per person-year [60]. For its usage in regions where interventions are in place, and low transmission levels have been accomplished, the model should be re-calibrated using age-disaggregated clinical incidence data of low-transmission settings. This would allow the identification of poorly constrained parameters, whose effect is negligible here but relevant in this alternative scenario.

### Implications for policy-making, interventions and malaria modeling

Malaria models can be used to support decisions such as where bed nets, medicines or funding are best distributed. Most models, however, describe whole communities with a single, average level of protection against the disease, in which every individual carries the same defenses and responds equally to infection. Acquired immunity against malaria is in reality an individual trait, and depends on the number of infectious bites a given individual has received over their lifetime. Our results show that this trait influences community prevalence and individual vulnerability, and that this influence depends on the local climatic regime, characterized, in this particular case study, by rainfall.

We find that a model built on a community average is effectively describing a particular demographic: late adolescents and adults, whose lifetime exposure happens to be closest to this average. For younger populations, this approach is off, and the degree to which these age strata are misrepresented depends on the disease transmission intensity, modulated by the local rainfall conditions.

The age group where this matters most from a public health perspective - children under 5 years of age - is also the most vulnerable. For this age group, the model based on a community average underestimates clinical infections and thus the total burden, with the largest differences simulated in regions where transmission is most intense. If this approach were to be used to allocate resources for child-targeted interventions, such as seasonal malaria chemoprevention (SMC) or insecticide-treated nets (ITNs), the resulting estimates would be too conservative, especially in environments where those interventions are most needed.

The opposite effect occurs in adult populations: the simplified model overestimates the presence of asymptomatic infections, and thus incorrectly predicts the potential impact of interventions on these transmission reservoirs. Since the simple approach to immunity fails to represent both the magnitude and age distribution of the asymptomatic reservoir, intervention impacts estimates cannot be derived from it. Indeed, asymptomatic individuals are still infective to vectors, and represent a widespread reservoir, even in low-endemic areas [47], that facilitates transmission that is difficult to reach with case-management strategies. An individual representation of acquired immunity can resolve this reservoir for different age groups, and is thus better positioned to predict the impact of intervention strategies targeting the disease transmission chain.

## Conclusion

Our results suggest that individually-acquired immunity against malaria is a strong modulator of community prevalence, representing a main source of structural modeling bias whose sign and magnitude depend on background climatic conditions. This bias is mainly concentrated below 15 years of age, and the age at which it peaks is determined by local transmission intensity, set here by rainfall conditions. The new VECTRI-ABM framework accounts for these structural biases by resolving immunity at the individual level, thus providing a finer-scale heterogeneous demographic distribution of malaria burden.

Climate-sensitive agent-based frameworks will become of increasing relevance for model-informed decision making [31], especially in low transmission settings approaching elimination, where stochasticity may play a major role and levels of spatial heterogeneity are high [25]. These models could also be used to finely assess changes in malaria immunity driven by potential changes in rainfall conditions with climate change over the African continent [80].

## Data Availability

All data produced in the present study are available upon reasonable request to the authors

## Acknowledgments

We would like to thank Dr. Jean-François Trape for providing the entomological and clinical data of the Dielmo case study. We also thank the Senegalese National Malaria Control Programme (*Programme National de Lutte contre le Paludisme, PNLP*) for providing access to the routine malaria surveillance data used in the seasonal evaluation.

## Supporting information

### S0. Agent-Based Model

We here provide a model description following the ODD (Overview, Design concepts, Details) protocol [41, 42]. The choice of using an established protocol responds to the goal of promoting a complete and rigorous formulation of the design of the ABM as well as its future use and reproduction. The protocol uses 7 predefined identifiers that run as a checklist covering all features that may define any ABM. We have also included parts already discussed in the Materials and Methods section, as it is the aim of the protocol to be self-contained, even if incurring in re-statements or redundancies.

### S0.1 Purpose

The ABM developed in this study represents an expansion of the VECTRI model, by which its original compartmental SEIR malaria module is replaced. Individual-level immunity acquisition against malaria represents a potentially major trait modulating malaria incidence. The ABM was here used to resolve this trait and study its relevance for different rainfall regimes in Senegal and The Gambia. Generally, this bottom-up approach allows disease dynamics to emerge as a result of individual attributes, such as behavior shaping human-vector interactions, cumulative exposure controlling immunity levels against *P. falciparum* malaria, age, sex or socioeconomic status.

### S0.2 Entities

#### Agents

The human population is described by *agents*, each representing a group of individuals with a number of attributes. For this study agents were given eight attributes, or state variables: an identity number, *n*_*ID*_ ∈ ℕ, a fixed spatial location, (*λ, ϕ*), an age, *a* ∈ ℝ, a vector attractiveness level, *ω* ∈ ℝ, an activity status, *ψ*_*a*_ ∈ *{*0, 1}, a maternal immunity status, *ψ*_*m*_ ∈ {0, 1}, and immunity level, *i*_*m*_ ∈ [0, 1], and an infection status, *s*. The infection status follows the standard *SEIAR* nomenclature, *s* ∈ *{S, E, I, A, R}*. These attributes will be discussed below and in the S0.7 - Submodels section.

#### Spatial and temporal scales

The ABM runs on an equirectangular lon-lat grid, whose spatial resolution is adapted to the lowest resolution of the input data (see S0.6 - Input data). Even though it can be used in a point-wise manner, it is meant to run on regional scales, thus neglecting village-level idiosyncrasies that could appear as further modulators of malaria incidence. In this study, each grid cell represents an area of ∼ 0.05^*o*^ × 0.05^*o*^ (∼ 5.5 × 5.5 *km* at the equator and ∼ 30 *km*^2^ in the region under scope). The model has a fixed time step, equal to 1 day, sufficient to represent changes in disease traits.

#### Environment

At each grid cell the vector density drives the dynamics of transmission, acting as a force that modifies the disease status of the agents. The model uses daily temperature at two meter height, *T*_2*m*_ (^*o*^*C*) and rainfall, *R*_*d*_ (*mm/day*) to drive the temporal evolution of the vector population, according to the VECTRI model (see S0.7 - Submodels). No direct interaction between the agents and temperature or rainfall is considered.

### S0.3 Process overview and scheduling

We here provide a schematic overview of the ABM’s processes in a single time step, this is, how agents’ attributes change, and in what order.

Before updating the agents, the VECTRI-ABM framework integrates the environmental entities, this is, the vector, larval and egg densities, according to the local meteorological variables *T*_2*m*_(*λ, ϕ, t*), *R*_*d*_(*λ, ϕ, t*), and the updating rule given by the VECTRI model (Fig S1). This implies an update over the spatial grid. The density of susceptible vectors becoming infected is calculated from the resulting human-to-vector transmission events of the previous update, *P*_*H*→*v*_(*t* − Δ*t*).

### S0.4 Design concepts

#### Emergence

While the agent-based structure of the framework itself plays a major role in determining incidence in low transmission areas, as stochasticity becomes a dominant factor, the main seasonality, intensity and geographical distribution of malaria transmission is tightly constrained by the underlying environment, this is, by the VECTRI model. Acquired immunity is, however, a heterogeneity trait expected to modulate clinical, or symptomatic, malaria incidence at the population level and as a function of the age class and climatic context in a non-trivial way. This individual-level trait thus constitutes the source of the system’s emergent behavior.

#### Interaction

No interactions among agents are modeled.

#### Stochasticity

Transmission events and changes in the maternal immunity and activity status are considered to be stochastic processes triggered by their corresponding probabilities. The transition from *E* to either *I* or *A* is treated in an equivalent manner. Each of these probabilities are functions of diverse agent attributes and the environment, as explained in S0.7 - Submodels.

**Fig S1.**
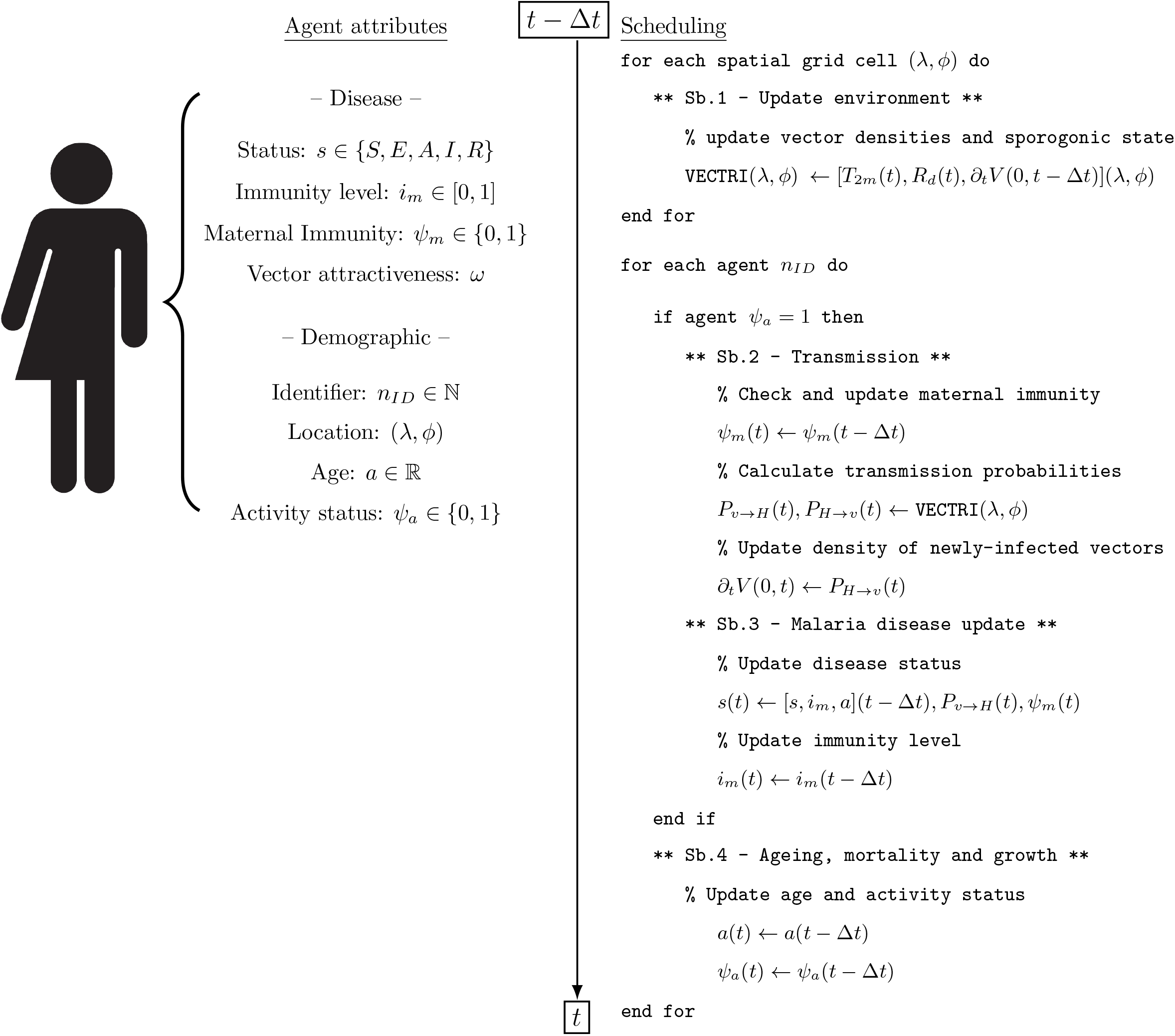
State variables, or agent attributes, and scheduling pseudo-code. Submodels (Sb.X) are described in S0.7 - Submodels. Illustration from NIAID NIH BioArt Source [81].

### S0.5 Initialization

#### Disease attributes

The immunity level is set to *i*_*m*_ = 0.5 for all agents, as this is relaxed to equilibrium during the Spin Up phase of numerical experiments (see this section’s **Model Spin Up**). The disease status, *s*, is randomly assigned. The agent’s attractiveness level, *ω*, is drawn from the log-normal distribution described in S0.7 - Submodels - Sb.2. No agents are initialized with maternal immunity, this is, *ψ*_*m*_ = 0. Maternal immunity is thus acquired during the Spin Up period when agents are re-activated (S0.7 - Submodels - Sb.4).

#### Demographic attributes

All agents are initialized as active, this is, *ψ*_*a*_ = 1. For each agent, both location (*λ, ϕ*) and age, *a*, attributes are assigned probabilistically from distributions derived from the WorldPop’s demographic estimates [67]. The number of agents in a given location, *n*_*i*_, is not directly proportional to the fraction of the total population present in that location, *h*_*i*_/∑_*j*_ *h*_*j*_. Given the limited number of agents in a simulation, *N*_*t*_, the framework first prioritizes their allocation in low density regions, in order to approach a human-to-agent ratio of 1 and maximally represent local attributes of interest, such as the population’s age structure. This is done by assigning agent locations as ∼ *ln*(*h*_*i*_ + 1)/ ∑_*j*_ *ln*(*h*_*j*_ + 1). In the limit where the number of agents matches the actual number of people in a region each agent then describes a single individual. The total number of agents is kept constant throughout the course of a simulation.

The WorldPop population density products provide gridded population counts by sex in different age bands (0, 1–4, 5–9, …, 75–79, 80+). Each grid value was here averaged over the study domain and the two sexes averaged together, giving a national fraction per band. Since these bands are unequal in width, they were transformed to single-year resolution by fitting a cubic spline to the cumulative distribution, evaluated at each band’s upper edge, and sampling it at every integer age from 0 to 79. The VECTRI-ABM model then aggregates all ages > 79 into the 79 years age band. Single-year weights were then recovered by first-differencing the interpolated cumulative curve. Interpolating the cumulative sum rather than the band densities directly allowed for a mass-conserving transformation: the spline passes exactly through the observed cumulative total at every band boundary, so the single-year weights within a band necessarily sum to that band’s observed share. The age of each agent, in years, is drawn probabilistically from the interpolated country’s age structure, assuming the age distribution to be spatially homogeneous in the region under scope - assumption that nevertheless needs to be checked (see S3. Age methods). The day of birth within this year is then drawn from a uniform distribution.

#### Model Spin Up

Each simulation starts with a relaxation time, where the first year in the climatic driving data (see S0.6 - Input data) is repeated until the difference between average immunity levels of two consecutive simulated years is below a prescribed threshold, *ϵ*. During Spin Up, the disease (*s*), activity (*ψ*_*a*_) and maternal immunity (*ψ*_*m*_) status, are let to freely evolve.

### S0.6 Input data

The VECTRI-ABM framework needs both climatic and demographic data to drive the underlying environment dynamics (VECTRI) as well as to set relevant agent attributes - their age and spatial location. For each grid cell in the system, and time step to be integrated, the framework requires daily mean air temperate at two meter height, *T*_2*m*_ (^*o*^*C*) and rainfall, *R*_*d*_ (*mm/day*), as well as the age distribution of the local human population and its density (*km*^−2^). Age and spatial density maps are considered to be constant throughout the course of a given simulation.

### S0.7 Submodels

#### Sb.1 Climate-driven vector ecology model: VECTRI

See Materials and Methods - Climate-driven vector ecology model: VECTRI.

#### Sb.2 Transmission scheme: agent-vector coupling

The coupling of both models takes place in the vector to human (*V* → *h*) and human to vector (*h* → *V* ) transmission processes, which in turn depend on the base interaction between the dipteran and vertebrate hosts, given by the human biting rate (*hbr* ). We here calculate this rate for the new VECTRI-ABM framework, from which either transmission probability is derived. Both transmission events are calculated on the agent level and, if successful, the corresponding agent, or a given density of susceptible vectors becomes infected, for vector to human and human to vector events, respectively (Fig 2).

##### Human biting rate

In VECTRI, at any given time *t*, the proportion of vectors searching for a blood meal is equal to the proportion that just laid eggs. This is regulated by the gonotrophic rate as

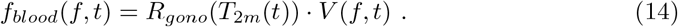

For each agent, *γ*, we then consider the vector-to-host ratio as base interaction rate, since higher human population densities relate to lower average EIRs [82]. The ratio is simply equal to *f*_*blood*_ over the local human population density, *h*. This approach, common in vector-borne disease modeling [48], yields the number of available vectors per person, assuming the former distribute equally among hosts. When the density of human hosts is big enough the interaction is rate-limited, this is, the vector search time is negligible compared to the reaction (biting) time. This reaction is in turn limited by the capacity of the vector population to interact with the host. On the other hand, when the human density is low, the dominant kinetic factor regulating the interaction is the quasi-diffusive time taken by the vector to find a rare host. This kinetic switch, between rate- and diffusion-limited processes is here modeled via a small correction of the vector-to-host ratio, yielding the base interaction rate

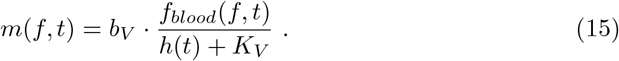

Here *K*_*V*_ is the half-saturation constant representing the diffusive properties of the vector. With this, when the number of humans in a given grid cell is low, the contact rate depends exclusively on the vector density ∼ *V*_*σ*_*/K*_*V*_ . *b*_*V*_ is the species-specific maximum interaction capacity (bites/vector/time). This biting rate is a function of the vector species [83, 84], hour of day [83–85], location (indoor/outdoor) [83–87], ecological setting (*e*.*g*., highland/lowland) [84] and the in-place intervention measures [86]. Notice the *f* dependency is not integrated away, as we put the focus on particular subsets of the vector population when addressing *h* → *V* or *V* → *h* events.

The agent-specific (*γ*) *hbr* is obtained by correcting for the number of people represented by each agent, or human-to-agent ratio, *r*_*h*_(*γ*), as

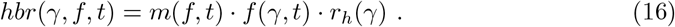

*f*(*γ, t*) is a modulation factor dependent on agent attributes that affect malaria incidence, such as individual and group behavior [88], use of interventions [89, 90] or socioeconomic characteristics [91]. These are out of the scope of this work and for this we set *f*(*γ, t*) = 1. We however emphasize the potential applications of this integrated approach to such circumstances, leaving it for future model developments. If modeled, a consideration would have to be made on the effect of the reduction factor in vector behavior, especially in interventions that might redirect risk from protected to unprotected agents, such as those involving untreated bed nets, effectively transferring risk to the local agent pool. This risk conservation hypothesis might be supported by observations where the average number of blood-fed mosquitoes does not correlate with the number of individuals in a given household [74]. Notice that without the *r*_*h*_(*γ*) correction Eq(16) would remain about the same independently on the number of agents, *N*_*t*_, in the system. This would then lead to an artificially higher number of bites for higher *N*_*t*_, as each agent would have the same interaction rate independently on the amount of people it represents. On a different note, the model suffers from a low *N*_*t*_ caveat. Since the *hbr* increases proportionally with *r*_*h*_(*γ*), when the demographic resolution becomes too low, the transmission probability (discussed below) quickly saturates to ∼ 1, overestimating infection events. Notice that if *r*_*h*_(*γ*) = 1 we return to the classical definition of the *hbr*.

##### Biting distribution

We expand the approach from [32], where the number of bites at the individual level are considered to be Poisson-distributed with a mean, *µ*, equal to the *hbr*. The new framework aims to account for population-level overdispersion in the biting distribution. Heterogeneity in exposure to anopheline mosquitoes is independent on the vector species [74]. This population-level overdispersion is here recreated from a bottom-up approach by considering biting to be heterogeneously distributed among agents. This is accomplished by assigning a heterogeneity degree, *ω*, to each agent. This degree is a constant characteristic of each agent, *ω* = *ω*(*γ*). Factoring in heterogeneity, the probability of an agent receiving *n* bites reads as

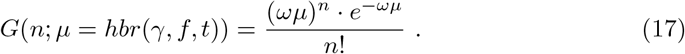

The heterogeneity parameter, *ω*, is strategically chosen to follow a Gamma distribution, fully parametrized by a vector, species-specific [74], dispersion parameter, *k*, this is

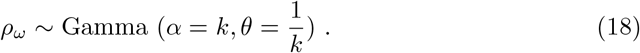

With this choice of the shape and scale parameters of the Gamma distribution we have

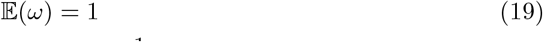

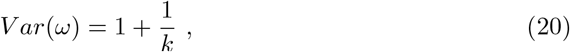

and thus *ω* can be understood as a deviation from the *hbr* (Eq 19) characterized by the overdispersion parameter, *k*, (Eq20) (*e*.*g*., Fig(S3)). The population-level bite distribution follows a Negative Binomial. This can be seen by integrating over all *ω* values (over all agents)

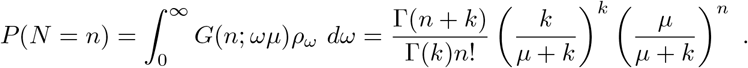

This distribution naturally captures the presence of “zeros” and clumping while preserving the total biting effort, as (see S2. Models of transmission)

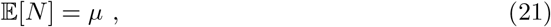

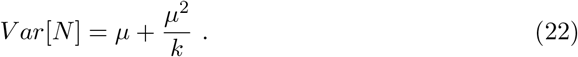

Within this framework, from an individual agent’s perspective, there are two ways to model *h* → *V* or *V* → *h* transmission (see S2. Models of transmission for details). The first uses the distribution *G*(*n*; *ωµ*) to calculate a number of events (bites) per time step and a number of infections. This method is costly, yet relevant for diseases where the severity of infection may depend on the number of successful infective bites [92, 93]. The second way only looks at the outcome “not infected” → “infected”. This simpler, coarse-grained approach that we adopt here, puts the focus on the probability to receive at least one successful infective bite. If the base probability of vector to human infection, or vice-versa, upon biting from a fully infective host is *P*_*i*_, then the probability of infection after *N* bites is

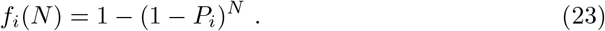

We adopt the nomenclature *i* = 0 for *h* → *V* and *i* = 1 for *V* → *h*. Then the desired transmission probability per unit time is

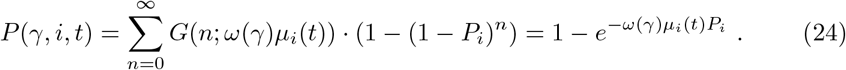

Where we select the adequate subset of the vector density for either transmission direction (*i* = 0, 1). This is, *µ*_*i*_ = *f*_*i*_*µ* with *f*_*i*_ = *V* (*i, t*)*/V*_*σ*_(*t*). Notice we dropped the *γ* dependency of *µ*, since we are from here on considering *f*(*γ, t*) = 1. In this context, *V* → *h* (*h* → *V* ) transmission solely comes from the fraction of vectors that are infective (susceptible), and managed to bite a susceptible (infective) agent. We consider the number of bites from vectors in different sporogonic states are independent. We can use the additive property of the Poisson distribution to see that the total biting “effort”, *hbr*(*γ, f, t*) = *µ* = ∑_*i*_ *µ*_*i*_, also yields a Poisson-distributed process with a mean equal to the sum of the means of all discretized subsets of sporogonic stages, and so the distribution and total “effort” of the bulk is independent on how we subdivide the vector population. A particular agent’s attractiveness is shared across any subset of the vector population, since the attractiveness of the human host, as captured by the *ω* parameter, is an intrinsic property, considered to be independent of the vector’s sporogonic state.

Of notice are the two different sources of overdispersion in our transmission model. We have a first layer accounting for deterministic, measurable covariates of transmission intensity, such as bed net usage (*f*(*γ, t*)), and a second layer, which accounts for covariates inaccessible at the individual level that might be reported in the form of biting heterogeneity across a given population (*k*) [74]. These can be, for example, volatile and non-volatile body odors [94], driving host-to-host attractiveness variability [95]. If we had *f*(*γ, t*) = 1 and *k* → ∞ then biting would be a process described by a Poisson distribution shared by all agents.

##### Vector to human transmission

If the base probability of vector to human infection upon biting from a fully infective vector is *P*_*v*0_, then the probability of an agent to be infected per unit time is

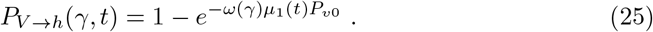

In the language of our model, the entomological inoculation rate averaged among all agents present in a given grid cell, *n*_*i*_, is

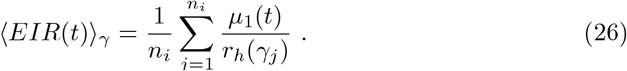

We can obtain an *EIR* stratified by different agent attributes by simply splitting the average accordingly.

##### Human to vector transmission

The probability of human to vector transmission for a given infected agent, either in the *I* or *A* states, reads as

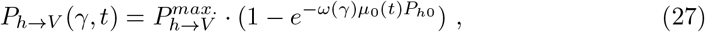

where 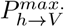 is the maximum transmission probability, partially estimated from [96, 97] (see Model calibration), and *P*_*h*0_ the human to vector probability of infection upon biting. The maximum transmission probability is a function of gametocyte density in the human host [96, 97] . In the absence of a with-in host model of parasite dynamics we here set it to be constant. Acquired immunity The influx of newly infected vectors thus reads as

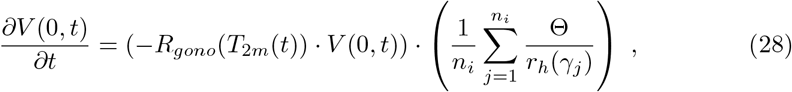

where Θ is a stochastic variable equal to 1 with probability *P*_*h*→*V*_ (*γ, t*), and zero otherwise. The sum is again corrected by the human-to-agent ration, *r*_*h*_(*γ*_*j*_), in order to ensure independency of results from *n*_*i*_ and *N*_*tot*_. The first parenthesis on the right hand side is the density of susceptible vectors searching for a blood meal, which is then multiplied by the success fraction, or the probability of host to vector transmission integrated across all infected agents in a given grid cell.

A summary of model parameters is provided in S1.b Model parameters.

#### Sb.3 - Malaria dynamics in the human host

This is a continuation of Materials and Methods - Malaria module.

##### Immunity dynamics

As our agent-based approach resolves the age distribution of the population we can explicitly consider maternal protection at the agent level. During maternal protection agents are immune to clinical malaria, while still susceptible to become asymptomatic. These asymptomatic agents still carry a parasite load and are thus infective to vectors. Maternal immunity is lost exponentially [45] at a half-life rate 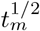.

The immunity level of a given agent rises a quantity, 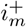, each time the agent is infected. Substantial immunity against severe malaria can develop with the first and second infections [45] while immunity against milder types is acquired more gradually, to the point it may never be complete [7]. Accordingly, we develop the two-scale scheme for immunity acquisition

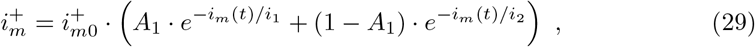

where *i*_1_ is the e-folding factor for boosted, fast acquisition, *i*_2_ that regulating the gradual posterior acquisition, 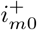 the maximum increased level per infective bite and *A*_1_ the weight of each acquisition mode.

Only agents in the recovered state, *s* = *R*, lose their immunity. This is lost exponentially at a half-life time 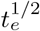, yielding the equation for the immunity level

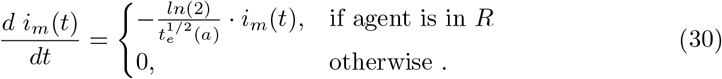

There is no universally agreed upon numerical half-life value for *P. falciparum*’s clinical immunity, as this depends on several factors such as host age and parasite antigenic type [98] as well as the differential persistence among different cellular components contributing to immunity [99]. Phenomenologically, in endemic areas, even though parasite prevalence roughly remains constant throughout age groups, the parasite burden decreases dramatically with age [48, 100] as clinical protection acts to regulate the density of parasitemia [7]. We here adopt the age-dependent loss term

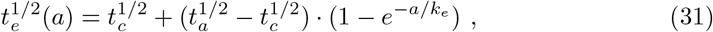

with 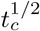 and 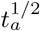 referring to the child’s and adult’s half-life, respectively, and *k* controlling the e-folding age of the transition. Broadly, the consensus lays in that clinical immunity is maintained by continuous exposure and wanes when this ceases, with age as modulator of the ability of individuals to clean infections [7]. A recent modeling study suggests, however, that immunity may play a role in regulating the production of blood-level parasite densities rather than increasing their decay rate [101]. In the absence of further evidence we opt for the above parametrization.

##### Probability of symptoms

The symptomatic probability, *α*, is modeled as a sigmoidal function of the immunity level, *i*_*m*_. This functional response has been used before when modeling the transition between symptomatics and asymptomatics with immunity acquisition [44]. The equation reads as

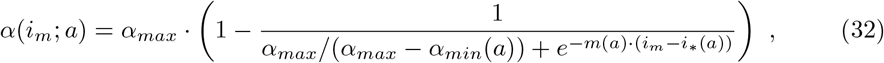

with *α*_*max*_ being the maximum fraction of a population presenting clinical malaria upon infection, *α*_*min*_ the minimum and *i*_∗_ and *m* free parameters controlling the immunity level and slope of the switch point in the sigmoidal curve, respectively. When *i*_*m*_ = *i*_∗_ the agent has 50% of being symptomatic upon infection. Studies in countries that present high levels of endemicity find diverse asymptomatic prevalences depending on age, community and testing method [102–104]. Further, evidence suggests that the pyrogenic threshold - the required parasite density to trigger a frebile reaction in the host - decreases with age in endemic areas, thus an adult’s immune system may respond more efficiently than a child’s, given similar degrees of parasite densities [105, 106]. Since the VECTRI-ABM framework does not incorporate within host parasite dynamics we here model the pyrogenic threshold and its shift with host’s age indirectly. We use *i*_*m*_ as a proxy for parasite immunity, assuming that an agent with high *i*_*m*_ will either tend to produce parasite densities below its given pyrogenic threshold or that it will simply tolerate higher densities. Since *i*_∗_ is the parameter representing the level of acquired immunity required for the agent to suppress a clinical febrile response, with a 1/2 probability, we consider it to be a function of the host’s age, this is

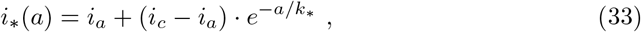

with *i*_*a*_ and *i*_*c*_ representing the immunity levels for 50% symptomatic probabilities in adults and children, respectively, and *k*_∗_ the e-folding age of the transition. With this we not only aim to capture the pyrogenic threshold, but its shift with age, representing long-lasting immune memory that the immunity level (*i*_*m*_) alone cannot fully capture. The threshold will then be lower for adults, this is *i*_*c*_ *> i*_*a*_.

Throughout a host’s life, exposure to diverse parasite variants, or antigens, results in the development of strain-specific immunity [7]. The clinical outcome of a new infection depends on the current infecting strain and the host’s long-term immune memory, characteristics that the *i*_*m*_ attribute may fail to capture. The *m* parameter in Eq(32) acts as a proxy for population-level heterogeneity in exposure history. A steep slope in the sigmoidal curve reflects an immunologically synchronized population that transitions from symptomatic to asymptomatic at similar *i*_*m*_ values. As this may characterize young, naive cohorts it fails to capture exposure history of older individuals. By decreasing *m*, the transition becomes more probabilistic, reflecting a higher degree of heterogeneity in clinical outcomes for similar *i*_*m*_ values across older agents. We here parametrize *m* as a decreasing function of the agent’s age, this is

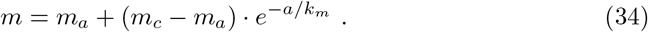

With *m*_*a*_ and *m*_*c*_ being the adult’s and children’s values and *k*_*m*_ the e-folding age for the transition. Lastly, host maturation of the innate immune system is modeled by parametrizing *α*_*min*_ as a function of age, as

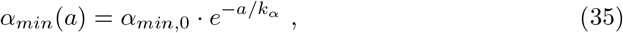

with *α*_*min*,0_ representing the symptomatic floor of newborns and *k*_*α*_ the e-folding age of transition into adulthood. This expression forces the clinical probability to trend towards a lower baseline as the agent gets older.

##### Drug treatment and recrudescence

Symptomatic agents might seek treatment and thus recover faster than their asymptomatic equivalents. Even though our symptomatic state, *I*, corresponds in practice to detected cases of malaria we make no assumption as to whether drug treatment is being used and clearance times are shared among symptomatic and asymptomatic cases.

Upon drug treatment, parasite clearance rates are drug specific [107], and some are known to trigger a drug-resistant state in *P. falciparum*. This drug-induced dormant blood-stage state of the parasite might later result in recrudescence. It is important to notice the difference between relapse, driven by dormant liver hypnozoites, associated to *P. vivax* and *P. ovale*, and recrudescence, or the return of the disease from incomplete clearance of these blood-stage persistent parasites, a phenomenon present in *P. falciparum* [96]. The proportion of a population suffering recrudescence varies wildly based on immunity and type of therapy. For example, in a study by [52], from 6 malaria-naive subjects infected with *P. falciparum* half manifested recrudescence from 11 to 27 days after infection and piperaquine treatment, a poorly efficient drug against these ring-stage dormant parasites [108]. In the absence of specific drug treatment parametrization we here consider a zero probability of recrudescence.

### Sb.4 - Ageing and mortality

Agents age at a daily time step. Age is thus treated as a floating number and is rounded down when aggregating agents by their age class.

A given agent may die with a probability *µ*_*d*_ [*day*^−1^], derived from an exponential fit of the population’s age structure, and be *re-activated* later. This *re-activation*, or growth rate, is calibrated so that growth and death rates at the population level are equal, thus ensuring a demographic equilibrium that also sustains the age structure. We set *ψ*_*a*_ = 0 when an agent dies and *ψ*_*a*_ = 1 when this is re-activated. Upon re-activation, the agent’s age is set to zero, *a* = 0, its disease status to *s* = *S*, the immunity level *i*_*m*_ = 0, the maternal immunity to *active, ψ*_*m*_ = 1, and a new attractiveness level, *ω*, is drawn from the aforementioned log-normal distribution.

## S1.a Clearance times

### Clearance time parametrization

In the malaria module (see Agent-based model of human health) clearance times are drawn from log-normal distributions, whose mean and standard deviation are functions of the immunity level, *i*_*m*_, of each particular agent. The functional relation is a simple interpolation between the corresponding parameters of log-normal distributions of naive (*Pf PR* ∼ 0%, Malaria Therapy dataset (MT)) [49] and holoendemic (*Pf PR* ∼ 75%, Ghana) [50] populations, which are considered to represent the limit scenarios *i*_*m*_ = 0 and *i*_*m*_ = 1, respectively. In Fig S2 we show the resulting parametrization of parasite clearance times.

**Fig S2.**
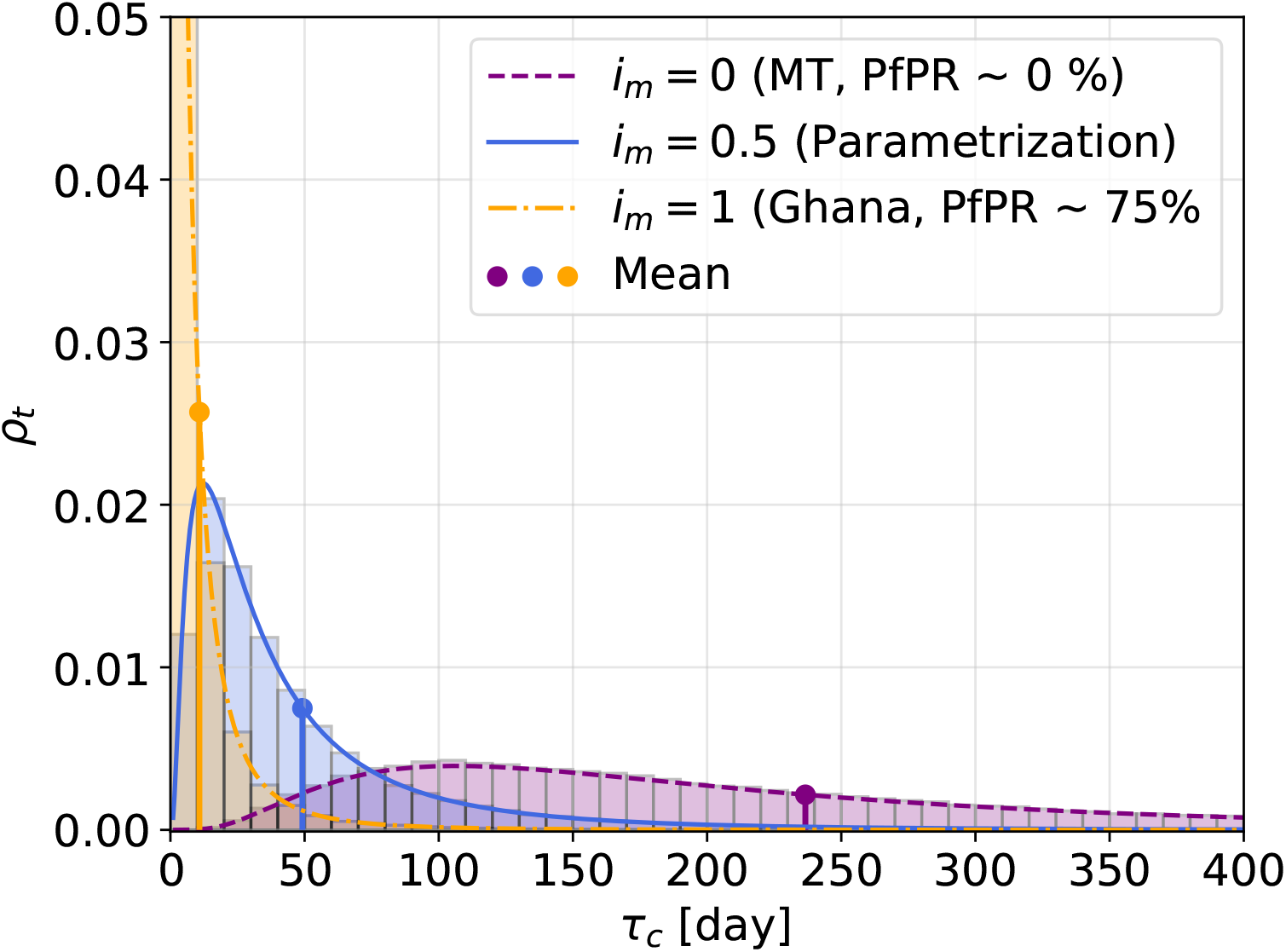
**Clearance times** of naive (*i*_*m*_ = 0), intermediate (*i*_*m*_ = 0.5) and holoendemic (*i*_*m*_ = 1) populations.

## S1.b Model parameters

**Table S1.**
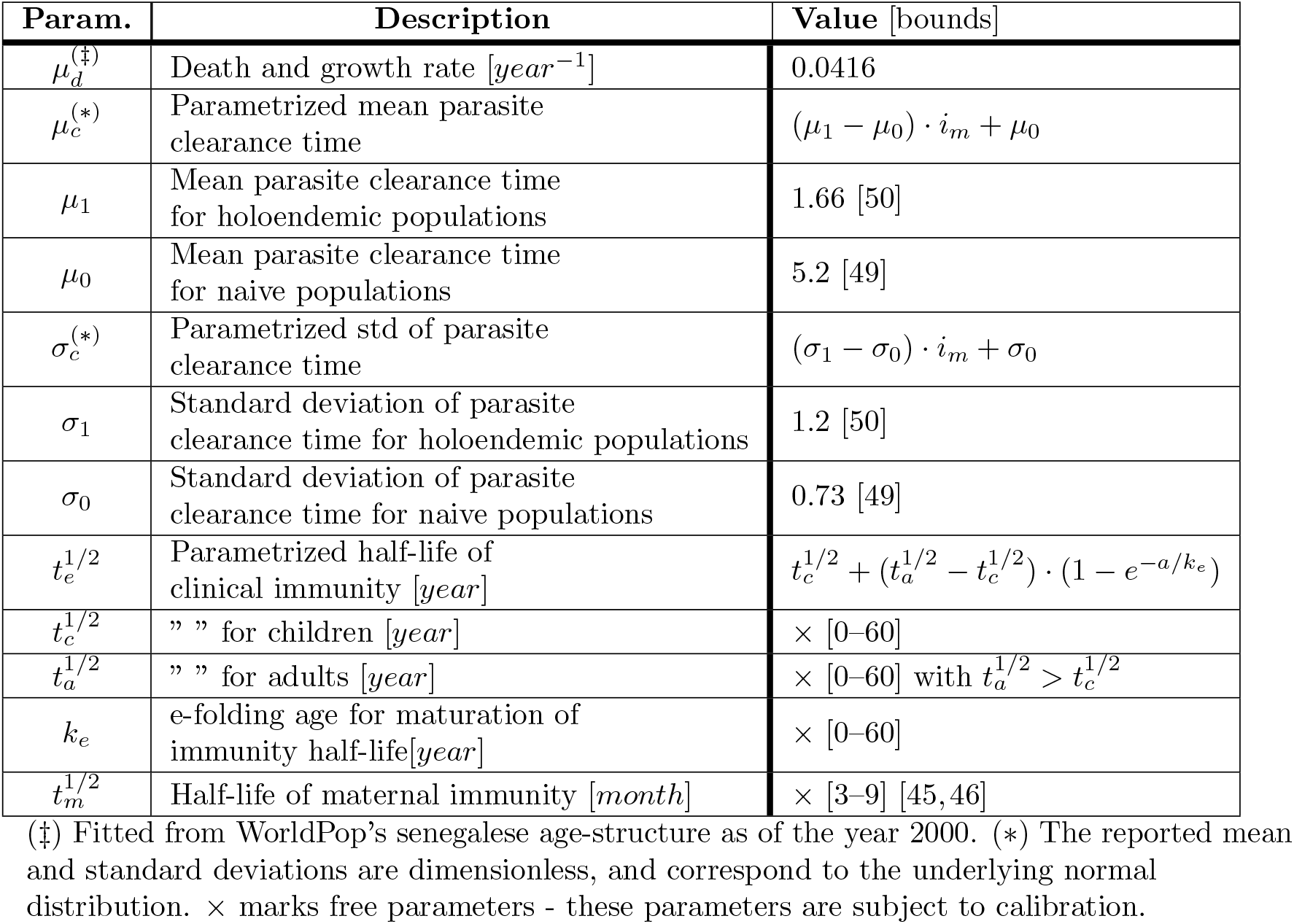
Mortality and Waiting times.

**Table S2.** Immunity dynamics.

| Param. | Description | Value [bounds] |
| --- | --- | --- |
| $i_m^+$ | Increase in immunity level [per infectious bite] | $i_{m0}^+ \cdot (A_1 \cdot e^{-i_m(t)/i_1} + (1 - A_1) \cdot e^{-i_m(t)/i_2})$ |
| $i_{m0}^+$ | Base increase in endemicity level [per infectious bite] | × [0 – 1] |
| $i_1$ | e-folding level of boosted immunity acquisition (fast) | × [0 – 1] |
| $i_2$ | e-folding level of gradual immunity acquisition (slow) | × [0 – 1] |
| $i_m^*$ | Threshold endemicity level value for the $R \rightarrow S$ transition | × [0 – 1] with $i_2 > i_1$ |
| $A_1$ | Weight of fast and slow modes of immunity acquisition | × [0 – 1] |

× marks free parameters - these parameters are subject to calibration.

**Table S3.** Symptomatics scheme.

| Param. | Description | Value [bounds] |
| --- | --- | --- |
| $\alpha(i_m)$ | Symptomatic fraction | $\alpha_{max} \cdot \left(1 - \frac{1}{\alpha_{max}/(\alpha_{max}-\alpha_{min}(a))+e^{-m(a)\cdot(i_m-i_*(a))}}\right)$ |
| $\alpha_{max}$ | Maximum symptomatic fraction | 1 |
| $m(a)$ | Slope of sigmoidal curve | $m_a + (m_c - m_a) \cdot e^{-a/k_m}$ |
| $m_a$ | Adult's slope | $\times [1 - 100]$ |
| $m_c$ | Children's slope | $\times [1 - 100]$ with $m_c > m_a$ |
| $k_m$ | e-folding age for slope switch from children to adult | $\times [0 - 60]$ |
| $i_*(a)$ | Pyrogenic threshold - immunity level of the switch point in the sigmoidal curve | $i_a + (i_c - i_a) \cdot e^{-a/k_*}$ |
| $i_a$ | Immunity level for 50% symptomatic probability in adults | $\times [0 - 1]$ |
| $i_c$ | Immunity level for 50% symptomatic probability in children | $\times [0 - 1]$ with $i_c > i_a$ |
| $k_*$ | e-folding age for pyrogenic threshold switch from children to adult | $\times [0 - 60]$ |
| $\alpha_{min}(a)$ | Minimum symptomatic fraction | $\alpha_{min}(a) = \alpha_{min,0} \cdot e^{-a/k_\alpha}$ |
| $\alpha_{min,0}$ | Minimum symptomatic fraction for $a = 0$ | $\times [0 - 1]$ |
| $k_a$ | e-folding age of minimum symptomatic fraction [year] | $\times [0 - 60]$ |

× marks free parameters - these parameters are subject to calibration.

**Table S4.** Agent-vector interactions.

| Param. | Description | Value [bounds] |
| --- | --- | --- |
| $IIP$ | Intrinsic incubation period [day] | 10 [43] |
| $k$ | Overdispersion parameter | 2.5 [74] |
| $b_V$ | Maximum vector biting rate [bites per vector per day] | $\times [0.1 - 20]$ [84] |
| $K_V$ | Half-saturation constant (vector diffusion) [$m^{-2}$] | $\times [10^{-5} - 10^0]$ |
| $P_{v0}$ | Vector to human transmission probability [per bite from infected vector] | $\times [0 - 1]$ |
| $P_{h0}$ | Human to vector transmission probability [per bite to infected human] | $\times [0 - 1]$ |
| $P_{h \rightarrow V}^{max.}$ | Maximum transmission probability | $\times [0.05 - 0.4]$ [96,97] |

× marks free parameters - these parameters are subject to calibration.

## S2. Models of transmission

### Population level

Recapitulating on the biting distribution scheme, with this interpretation biting is heterogeneously distributed among agents. We, however, don’t have access to individual-level attractiveness, but to its distribution across a given population - parametrized by the overdispersion coefficient *k*. In our setting, overdispersion is modeled at the individual level by assigning a different heterogeneity parameter, ω, to each agent. We choose this heterogeneity parameter to be gamma-distributed. By choosing the shape and scale parameters of the gamma distribution

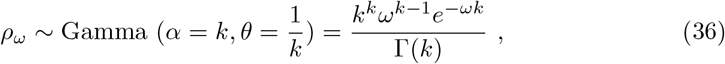

the population level bite distribution, *P*(*N* = *n*), follows a Negative Binomial. Indeed, integrating over all ω values we get

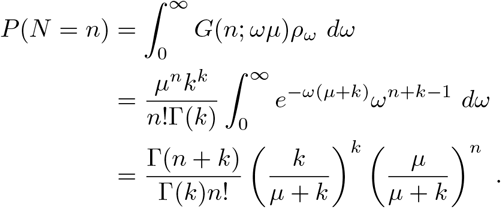

We used the identity 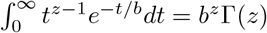 in the last step. With this model for overdispersion we conserve the global effort, *µ*, while allowing for a greater variance. Using the Moment Generating Function,

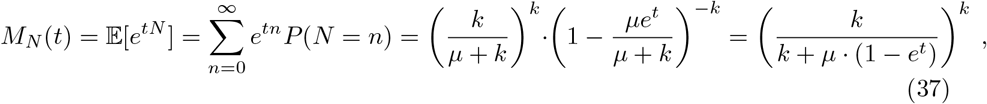

which we obtain from the Binomial series identity

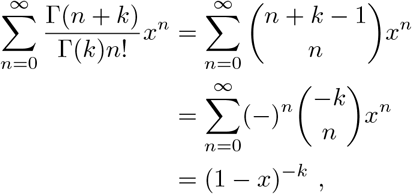

valid if |*x*| *<* 1, we see that

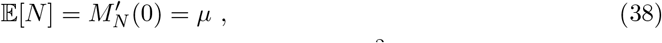

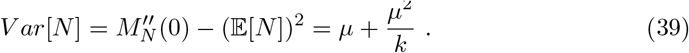

When *k* → ∞ we then recover the homogeneous Poisson model for host-vector interactions. In this sense, 1*/k* can be viewed as an “aggregation” coefficient. The overdispersion parameter, *k*, is empirically accessible (*e*.*g*., [74]), and varies across vector species and very likely across host populations.

### Implications for transmission

The overdispersion (*k*) scheme allocates an attractiveness level (*ω*) to hosts (agents), making them more or less susceptible to be bitten, with respect to some average value (*µ*). This trait modulates disease transmission, and has diverse implications at the population level, more or less relevant depending on the endemicity of the disease in the population under study. Firstly, in endemic areas, both seasonal and perennial, the cumulative effect of overdispersion has an impact on modeled prevalence estimates. Secondly, in epidemic or disease-free areas, it changes the *R*_0_ value, or the necessary conditions for the disease to “percolate” across the population.

### Transmission in endemic areas

At the population level there is an inequality between both (Poisson and NB, or homogeneous and heterogeneous Poisson models) probabilities of transmission. For the heterogeneous Poisson model we have

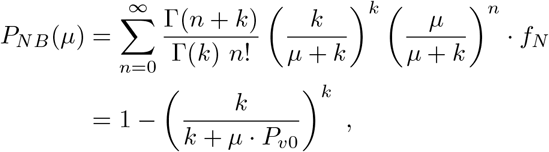

where we again used the Binomial series identity, and re-arranged terms. Mathematically we see the aforementioned inequality starting from

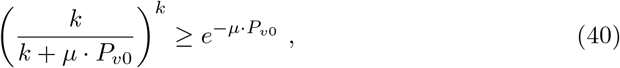

taking the reciprocal on both sides, thus reversing the equality,

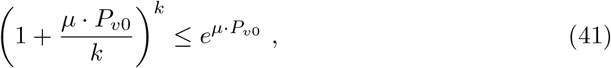

and noticing that the limit

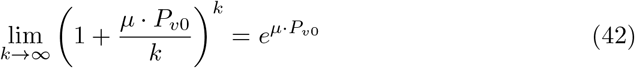

is approached from below. We then have that

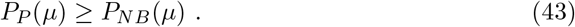

This can be understood at the agent level. Biologically, a lower population-level transmission can be caused due to saturation, whereby a high percentage of successful mosquito infections will be allocated on already infected (very attractive) hosts. Since the infection probability *f*(*N*) is concave,

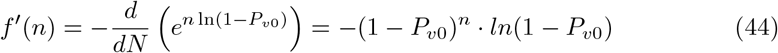

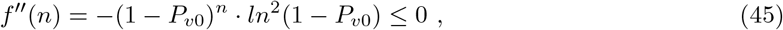

when the number of bites are pushed further to the extremes - many hosts will receive zero bites, whereas some hosts will receive an elevated number of bites, as compared to the homogeneous Poisson model - the transmission probability quickly saturates, effectively “wasting” those bites for population-level transmission purposes. The homogeneous Poisson model, on the other hand, has a more “efficient” allocation of infective bites, as the distribution number is more “moderate”.

This estimate ignores model dynamics, in its complexity, including down-stream, secondary transmission events. In this sense, it represents a lower-bound, already at a cumulative difference of 20% for some mean biting rates (Fig S4).

### Transmission in epidemic or disease-free areas

Estimations of *R*_0_ when overdispersion is resolved in simpler mechanistic models yield the correction

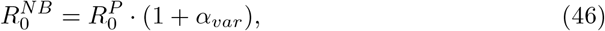

with 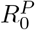 representing the standard value without oversdipersion and *α*_*var*_ being the squared coefficient of variation (*σ*^2^*/µ*^2^) (see [74] and references therein). The transmission threshold is therefore easier to meet, effectively needing higher control efforts than a homogeneous model would suggest. This is again caused by the presence of highly attractive individuals.

### Models of transmission: Tables

**Table S5.**
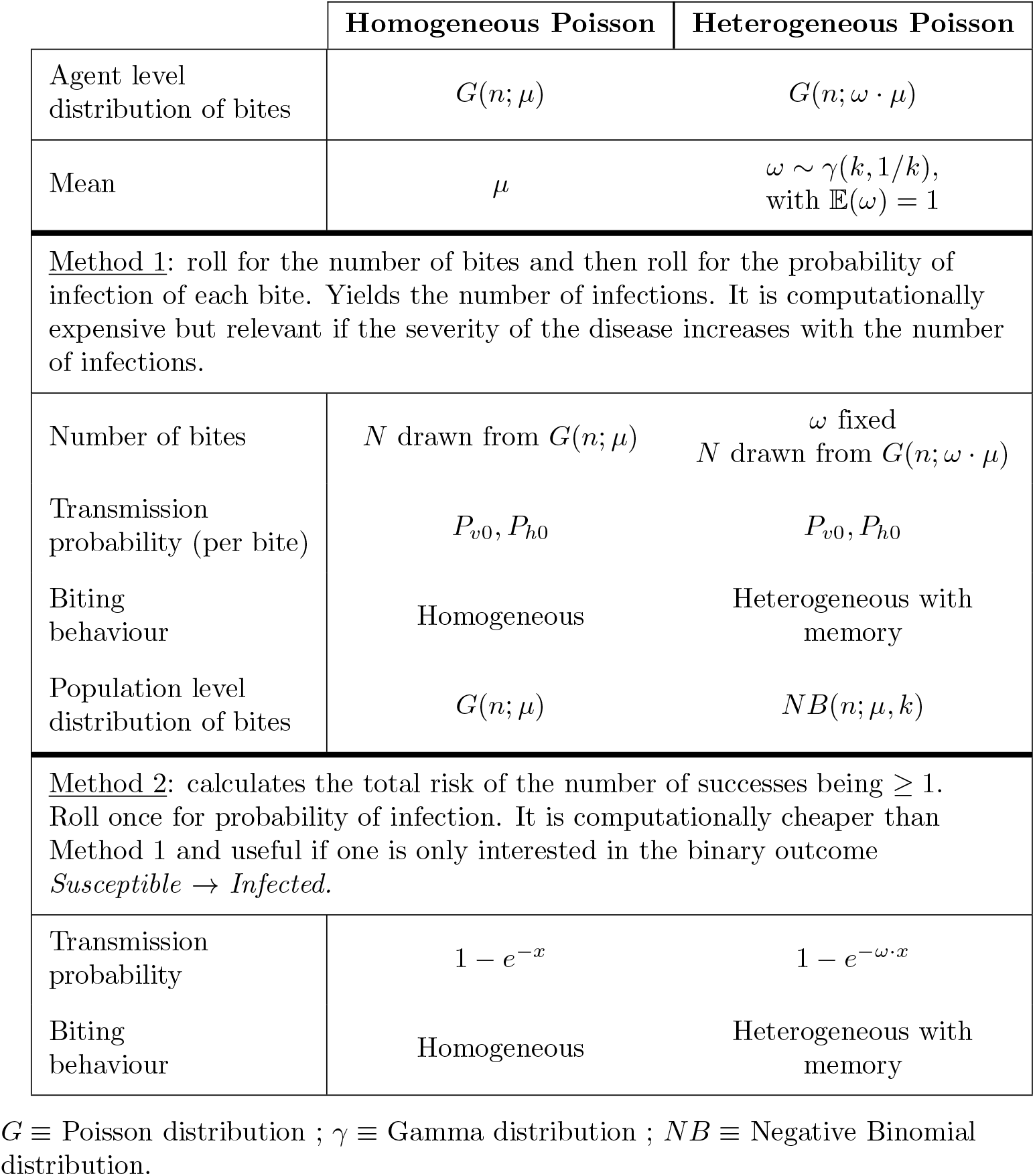
Agent-vector transmission schemes.

### Models of transmission: Figures

**Fig S3.**
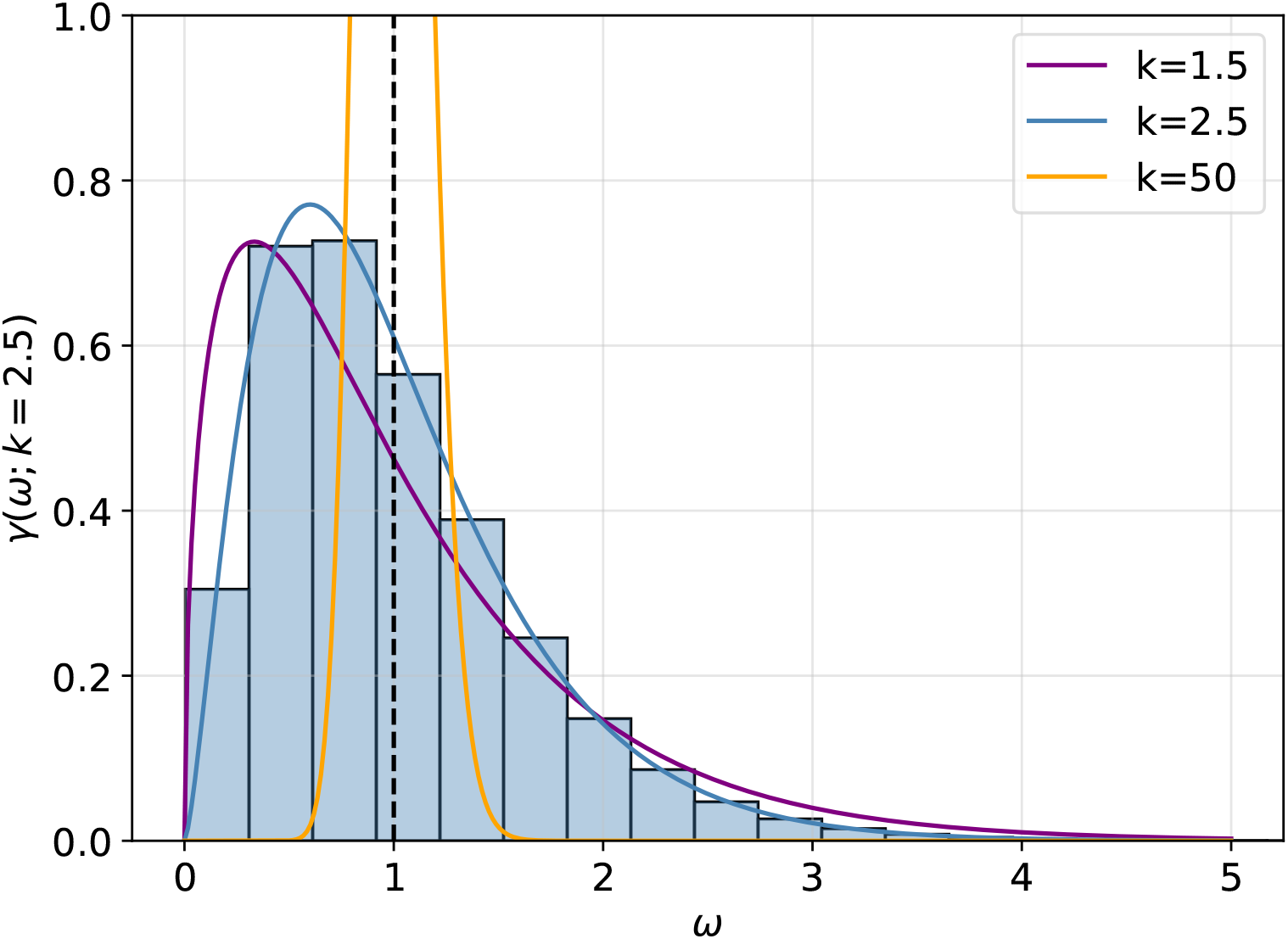
Gamma-distributed heterogeneity, *ω*. Gamma distributions for three different values of the overdispersion parameter, *k*. As *k* increases the modeled aggregation, 1*/k*, decreases, eventually recovering a homogeneous Poisson model.

**Fig S4.**
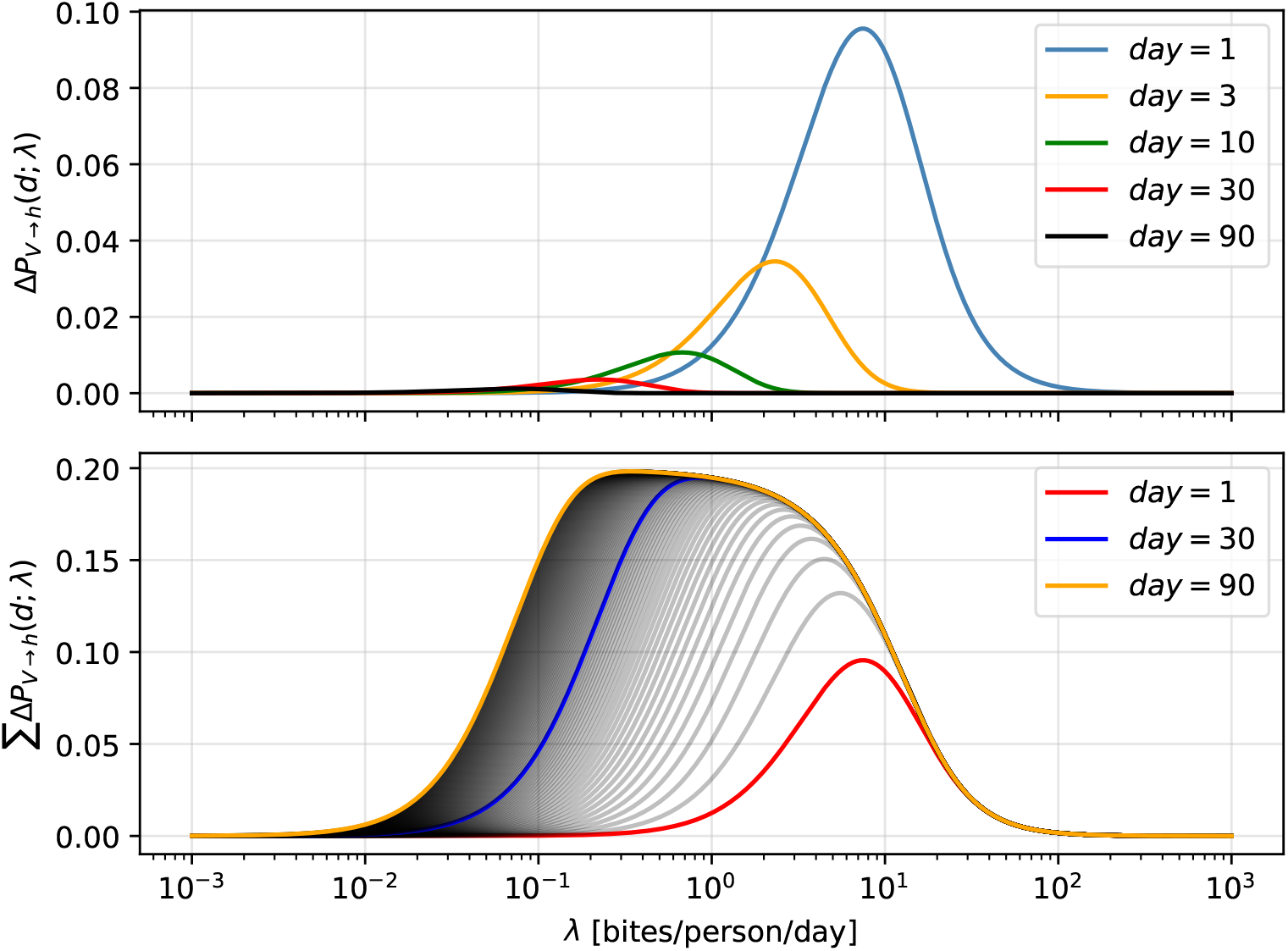
Probability differences between transmission schemes. Top. Difference in the probability of infection after *d* days as a function of the biting rate, labeled here as *λ* (*hbr* in the main text). Bottom. Cumulative difference of the previous after *d* days.

The expressions for the plots are obtained by considering a biting rate, *λ*, a success probability, *P*_*v*0_, and an overdispersion, *k*. Then the probability of infection after *d* days for the Poisson and Negative Binomial models reads as

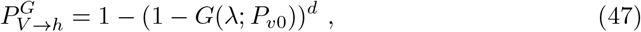

and

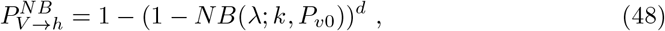

respectively. The difference shown is 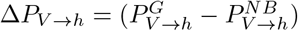

## S3. Age methods

Spatial heterogeneities in the age structure are assessed by calculating the percentage different age classes, or age ranges, represent among the local population of a given grid cell. We perform this calculation for all grid cells, and histogram the results (Fig S5). If the total population has an homogeneous spatial distribution then all counts will lie at the same percentage. From this analysis we see that spatial homogeneity in the country’s age structure is a good working hypothesis.

**Fig S5.**
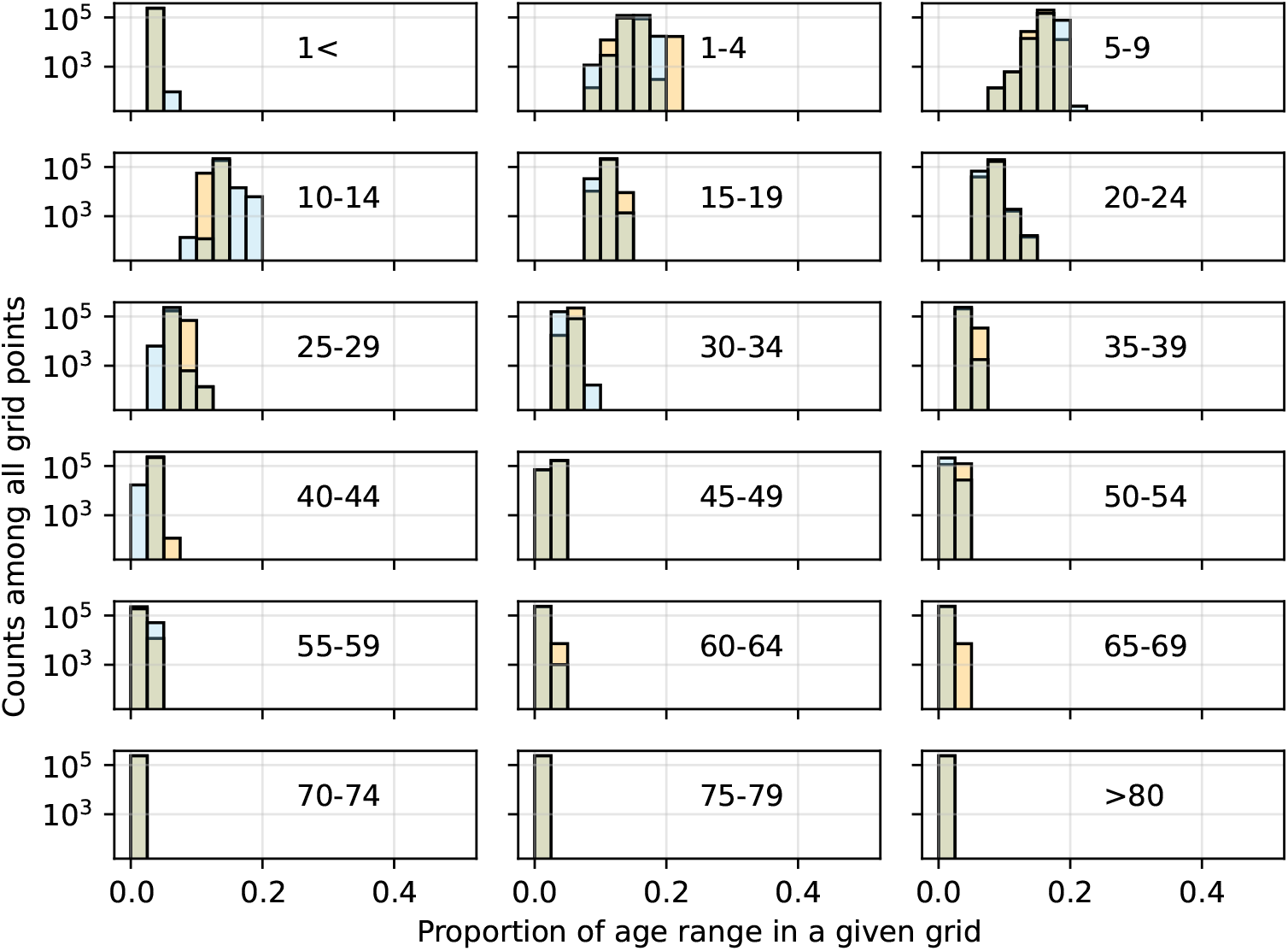
Spatial heterogeneity. Frequency of the proportion of the total population represented by each age class. Each count represents a single grid cell. The frequency is the histogram of counts from all grid cells of the simulated domain.

Senegal’s age structure as of 2000 WorldPop estimates, its interpolation and the values from the agents initialized in VECTRI-ABM.

**Fig S6.**
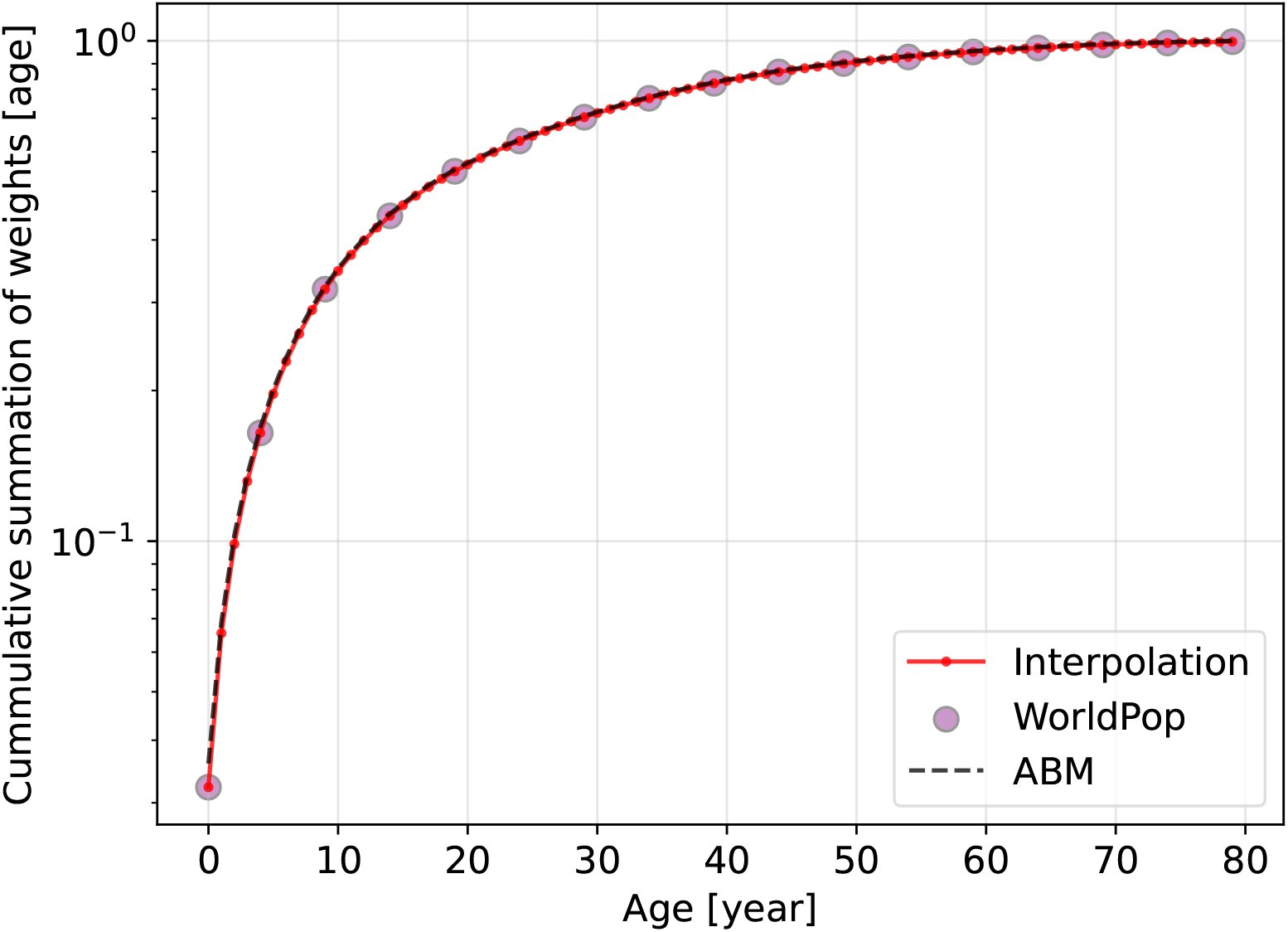
Age structure. Senegal’s age structure interpolated from WorldPop’s 2000 estimates and applied homogeneously to both Senegal and The Gambia as per the methods in S0.5 - Initialization.

## S4. Supplementary plots

**Fig S7.**
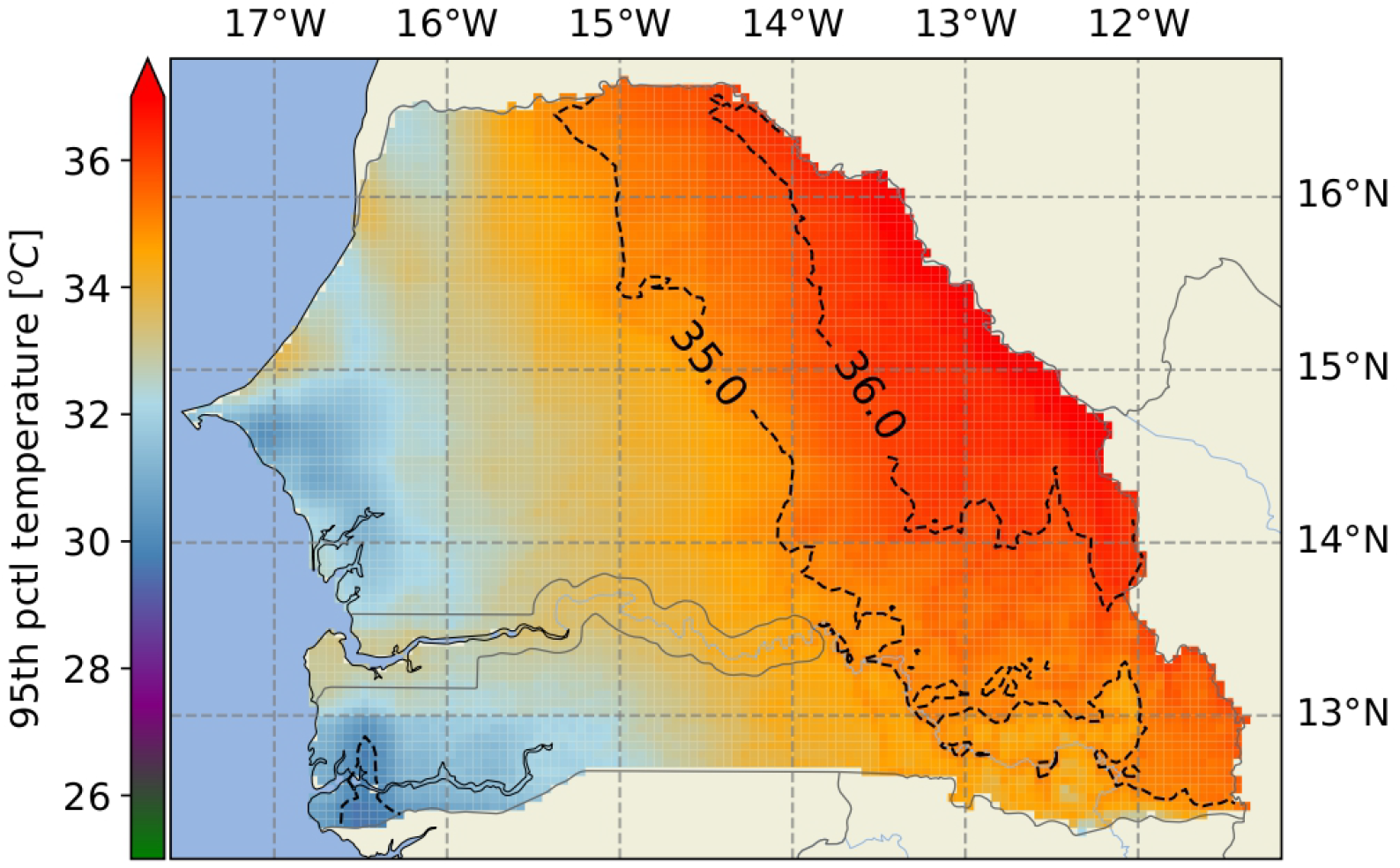
**Higher range of the two-meter air temperature**, *T*_2*m*_. 95^*th*^ percentile temperatures in the studied region during the period 1990–2003. Most values are well within suitable temperature ranges for the development and low mortality of *An. gambiae s*.*l*., with high mortalities only being reached in eastern areas, where values may exceed ∼ 37^*o*^*C*.

**Fig S8.**
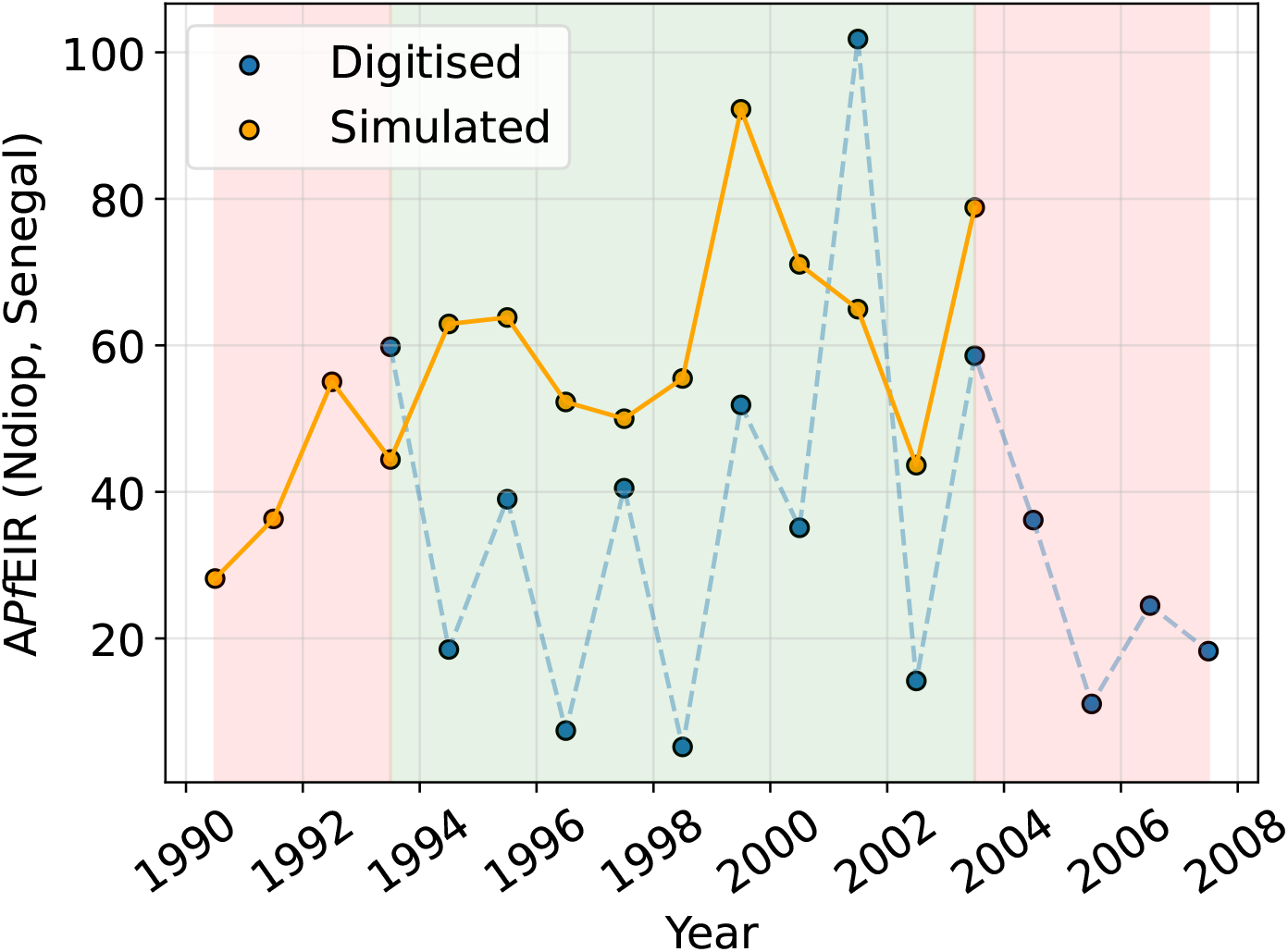
Ndiop. Year totals of *Pf* EIR (A*Pf* EIR) from VECTRI-ABM (simulated) and from [8] (digitized). The latter are the sum of digitized monthly totals. The green window marks the overlapping period between simulation and available data, with red marking the opposite. Simulated values are those resulting from the calibration exercise.

**Fig S9.**
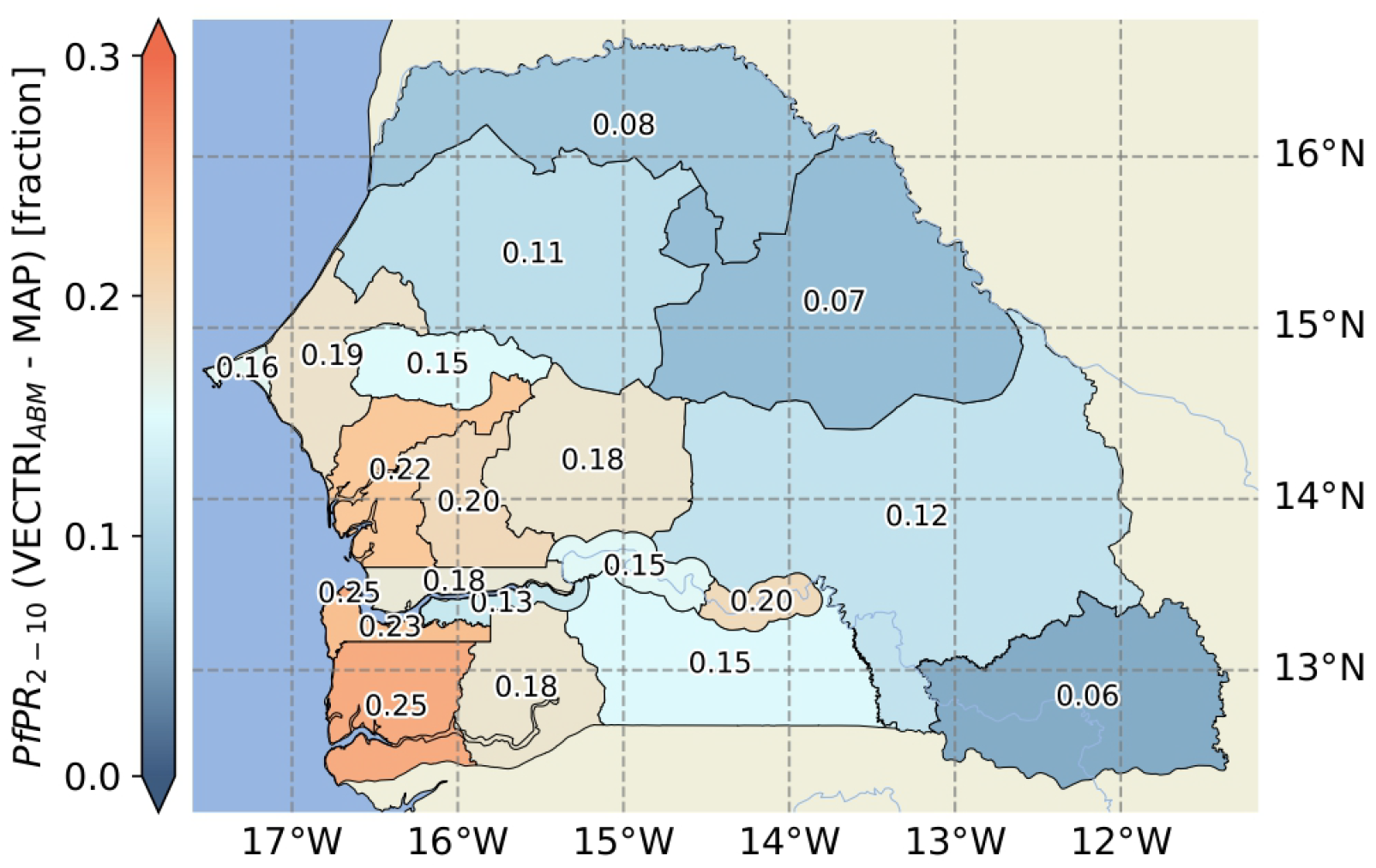
Raw VECTRI-ABM – MAP *Pf* PR_2–10_ comparison. As Fig 5, without the detection factor (*k* = 1). The color bar spans three times the range of Fig 5.

**Fig S10.**
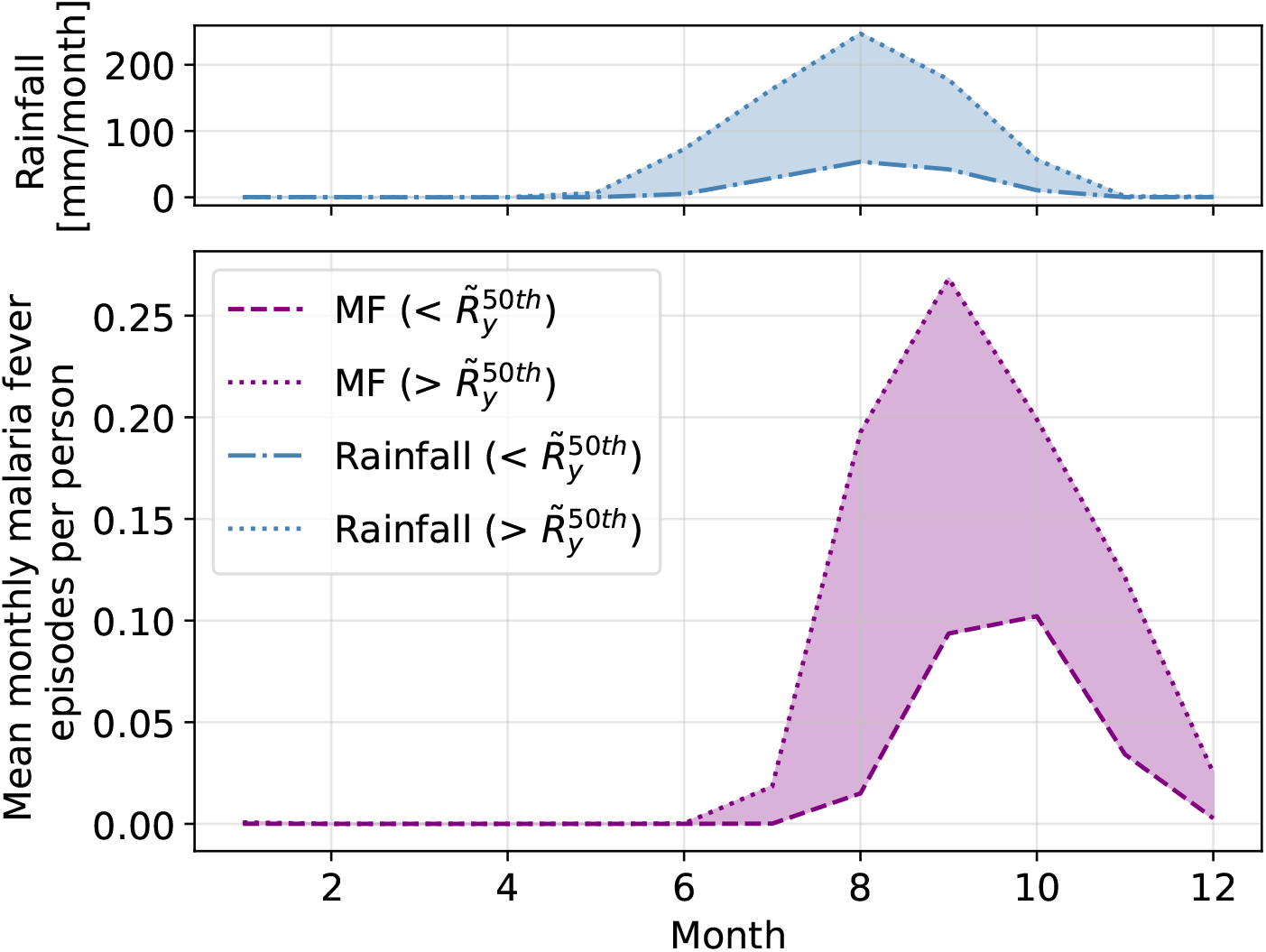
Simulated rainfall and malaria seasonality. Seasonal cycles of simulated rainfall and malaria fever episodes used to assess their relative timing. Cycles have been calculated by splitting the grid into its wetter and dryer parts, as defined by the spatial median of the time-averaged (1990–2003) yearly totals, 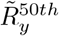.

**Fig S11.**
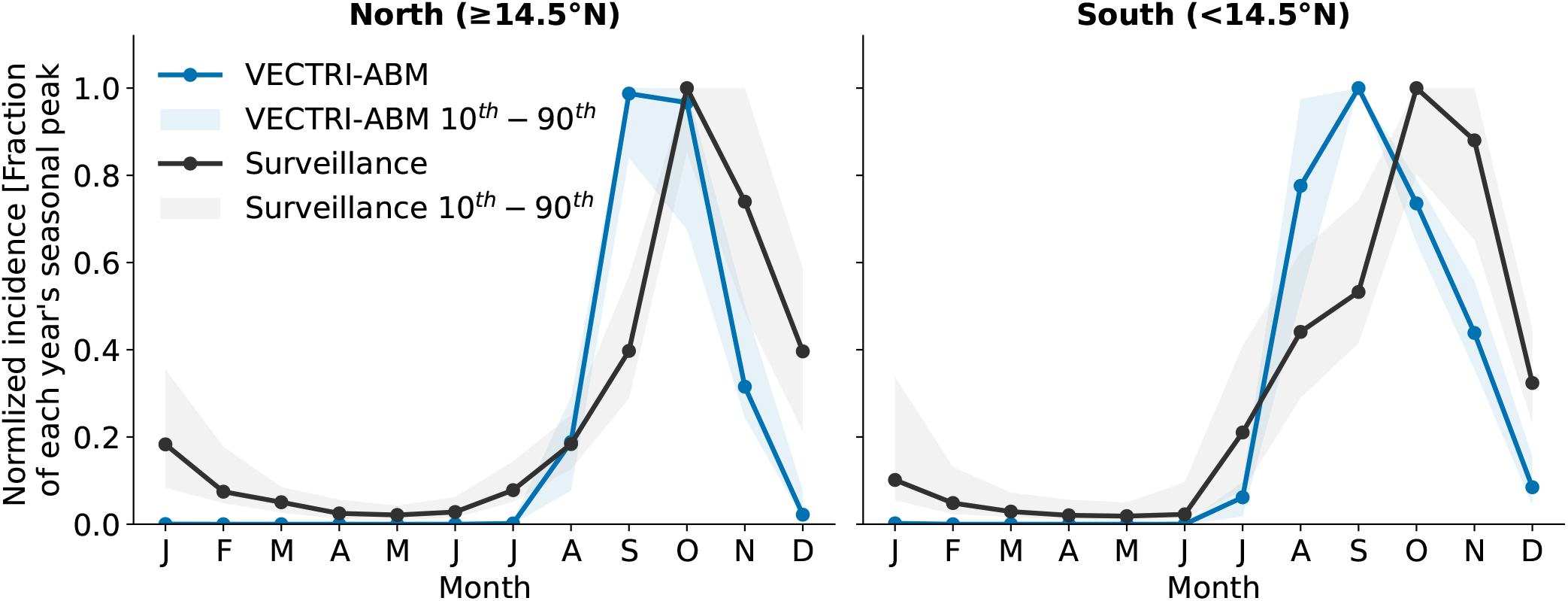
North-south comparison of normalized malaria seasonality. Normalized seasonal cycle of malaria incidence in the northern (≥ 14.5^°^N) and southern (*<* 14.5^°^N) zones of Senegal: simulated malaria fever episodes (VECTRI-ABM, 1990–2003) versus confirmed cases from the *PNLP* surveillance system (2009–2025). For each year, monthly values are normalized to that year’s maximum; solid lines show the across-year median and shaded envelopes the 10th–90th percentile range. In both zones the model peaks one month earlier than the surveillance (September versus October); summary metrics are given in Table 1.

**Fig S12.**
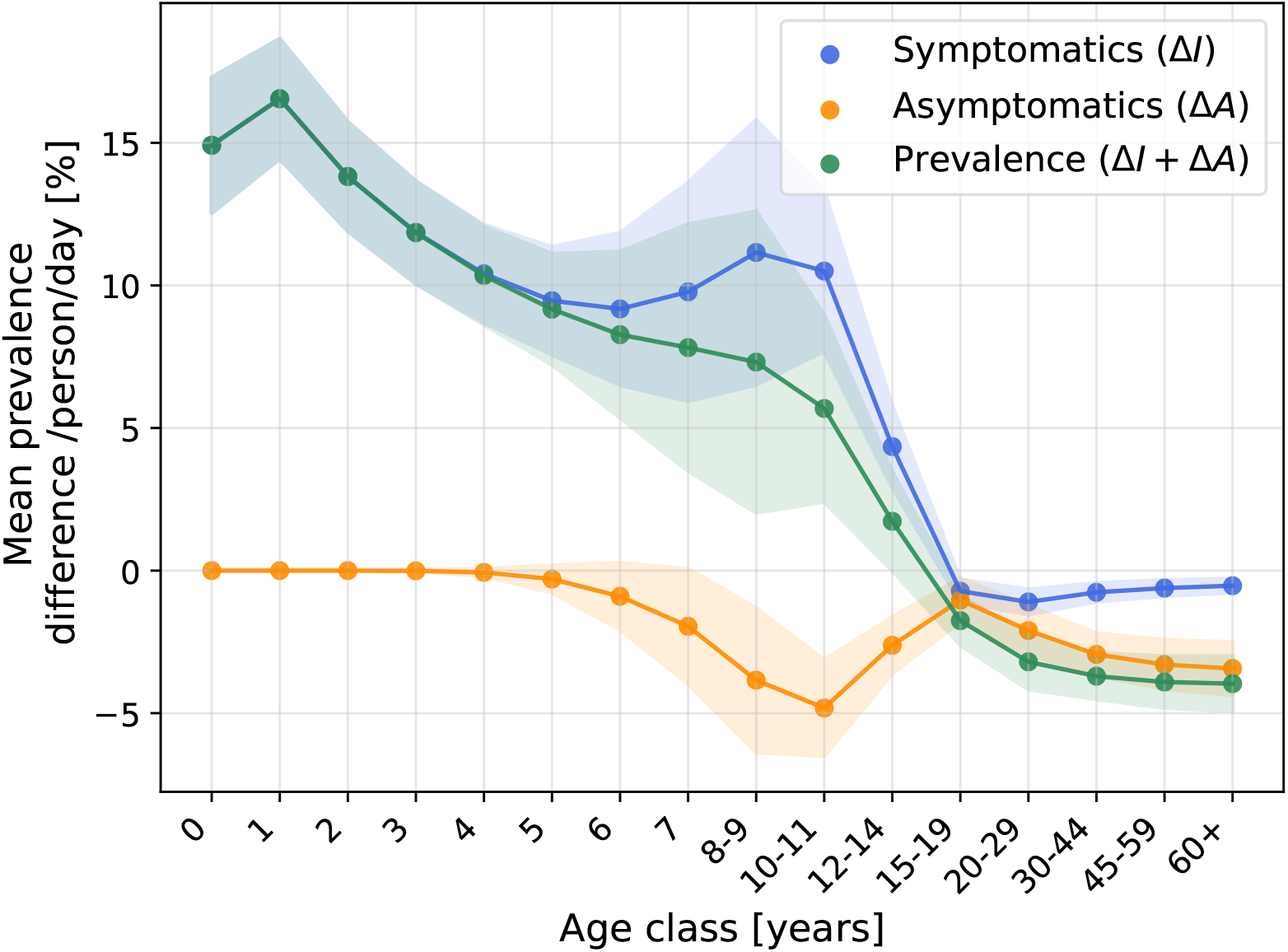
E2. Knock-out of maternal immunity. Symptomatic, asymptomatic and total mean prevalence biases in a scenario with no maternal immunity, this is *ψ*_*m*_ = 0. Summary statistics are calculated as in E1 (Fig 8).

**Fig S13.**
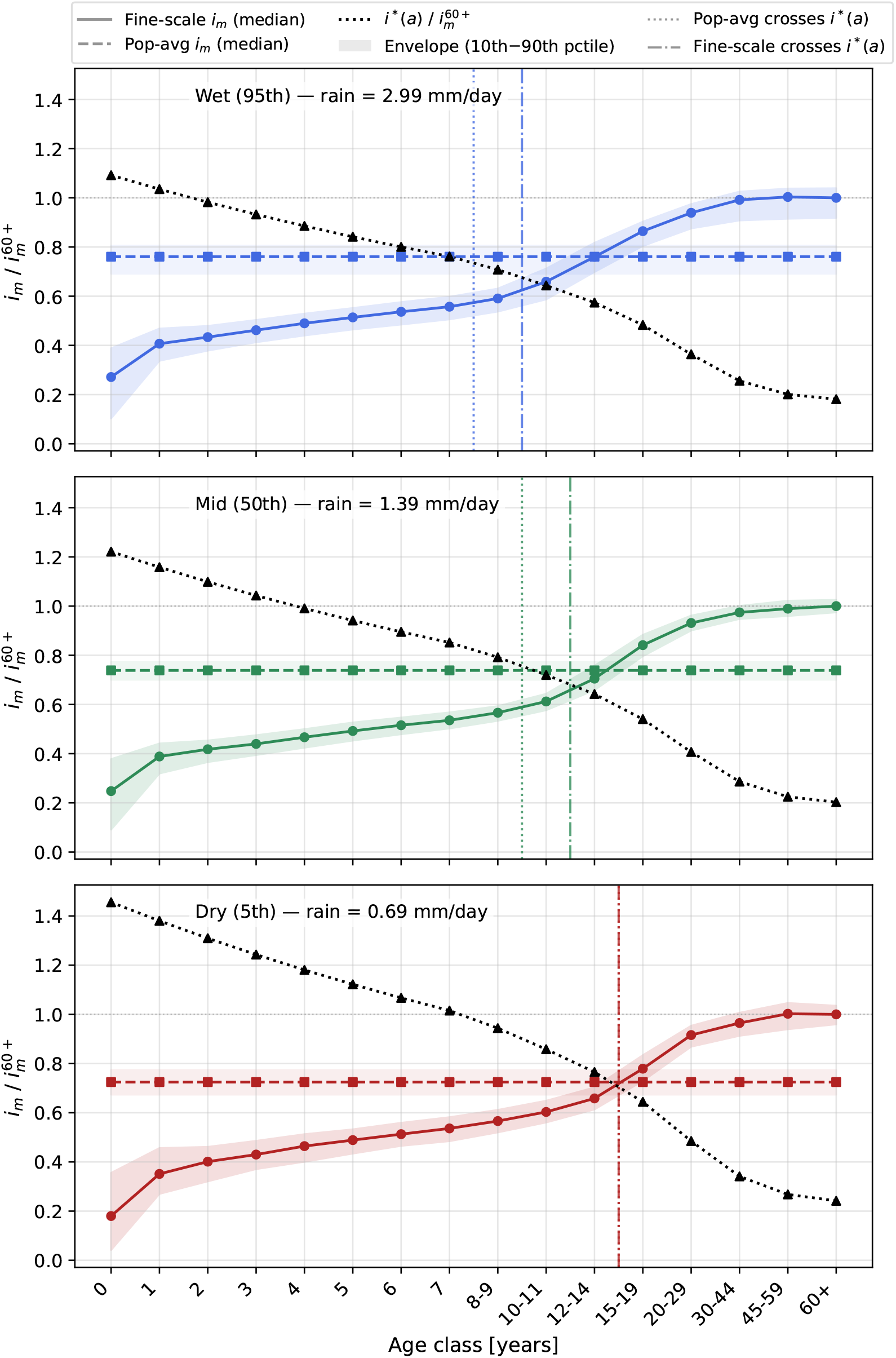
E1. Crossing the pyrogenic threshold. Pyrogenic threshold, *i*^*^(*a*), overlaid with immunity values of *pop-average* and *-fine-scale* simulations as a function of age, *a*. The crossing analysis is broken down into three single locations, selected by spatial percentiles of mean daily rainfall, in order to illustrate the differential crossing times as a function of transmission intensity. Immunity values shown are the time median (scatter points) and 10*th* and 90*th* time percentiles (envelopes) of the selected locations. For comparison purposes, immunity values are normalized by the immunity of the older age class, *i*_*m*_(60+).

## S5. Calibration

In this study we have calibrated both parameters specific to the new agent-based model as well as parameters controlling vector ecology, from the VECTRI model. In VECTRI, decay rates and advection scheme parameters (see Eqs 5-7) are specific to each vector. Different species have diverse breeding preferences (rural, urban, peri-urban) as well as climatic thresholds for the gonotrophic cycle and parasite and viral developments. We here calibrated parameters controlling developmental rates and availability of breeding sites to capture the regional context of the study.

### Development rates

Gonotrophic and sporogonic rates are modeled following the degree-day concept from Detinova [48, 109], a measure of cumulative heat above a given developmental (temperature) threshold. The concept translates into the linear function of temperate

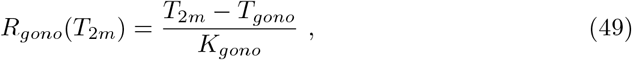

for egg development and

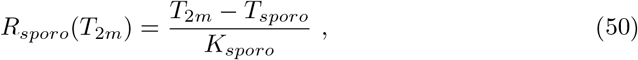

for parasite development. Here *T*_*gono*_ and *T*_*sporo*_ (^*o*^*C*) are the minimum air temperatures for development and *K*_*gono*_ and *K*_*sporo*_ (^*o*^*C* · *day*) the number of degree-days necessary for a complete development. By definition, both advection velocities can be interpreted as fractional growth rates. In this study we calibrated *K*_*gono*_ and *K*_*sporo*_.

### Carrying capacity

Temperature-dependent mortality terms are fixed functions fitted to empirical data, for a given vector species. On the other hand, mortality associated to the availability of breeding sites is highly dependent on the regional context under scope. Availability of permanent and temporary breeding sites will determine the dynamics of the system’s carrying capacity and, in turn, the larval mortality. In VECTRI, this is modeled via the “crowding” larval mortality term, which is composed by three factors: predation, crowding and rain-driven flushing as

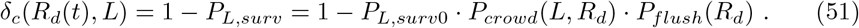

Here *R*_*d*_ stands for daily rainfall (*mm day*^−1^). *P*_*L,surv*0_ is taken as a constant. Since no parameters in the flushing term were calibrated, for an explicit development the reader is referred to [32]. The *crowd* term factors in the availability of breeding sites.

Particularly, it takes the form of a logistic equation, whose carrying capacity is determined by the maximum larval surface density biomass, *M*_*max*_ (*mg m*^−2^), and the fractional area of potential breeding sites in a given grid cell, *ω*(*R*_*d*_). If *M*_*tot*_ is the total surface larval biomass, then

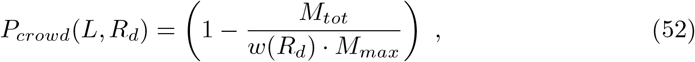

with

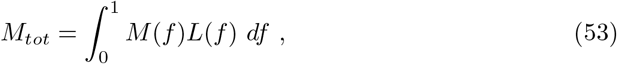

and 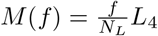. *L*_4_ is here the biomass of a single larva in its fully-grown stage. As mentioned before, the total availability of breeding sites is vector-dependent. The vector *Anopheles gambiae s*.*l*. is considered to be a rural vector that proliferates in temporary ponds [110]. On the other hand, the vector *Anopheles funestus* thrives near shady permanent or semi-permanent water bodies, such as swamps, marshes or in the vicinity of river streams [111–113]. The study location, used to calibrate the modeling framework, is located near the Nema river, feature that allows the year-round proliferation of *Anopheles funestus*, which is accompanied by *Anopheles gambiae s*.*l*. activity during the rainy season. Indeed, a previous analysis on Dielmo data found a significant correlation between rainfall and *Anopheles gambiae s*.*l*.’s EIR, whereas none was found for *An. funestus* [8]. Consequently, for the calibration step, we consider *ω*(*R*_*d*_) to be composed of the two terms

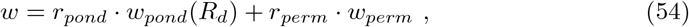

with *r*_*i*_ representing vector-specific coefficients, and *ω*_*i*_ the fractional contributions - from permanent (*ω*_*perm*_) and temporary ponds (*ω*_*pond*_) - to the total breeding site availability. For more information on the temporary pond dynamics, *dw*_*pond*_(*R*_*d*_(*t*))*/dt*, the reader is referred to [57, 58]. In this study we calibrated the parameters *r*_*pond*_, *r*_*perm*_ and two parameters from the temporary pond scheme: the curvature number, *CN*, and the maximum infiltration rate, *I*_*max*._.

Importantly, for the numerical experiments, after calibration, we set *r*_*perm*_ = 0, as this was only used temporarily to calibrate immunity parameters using data from the Dielmo project, where *Anopheles funestus* was present. The rest of the study is, however, performed on a regional scale, focusing on the rain-driven vector *Anopheles gambiae s*.*s*.

## S5.1 - Calibration tables

ToC

In the following, we present best-fit point estimates of the model parameters.

**Table S6.**
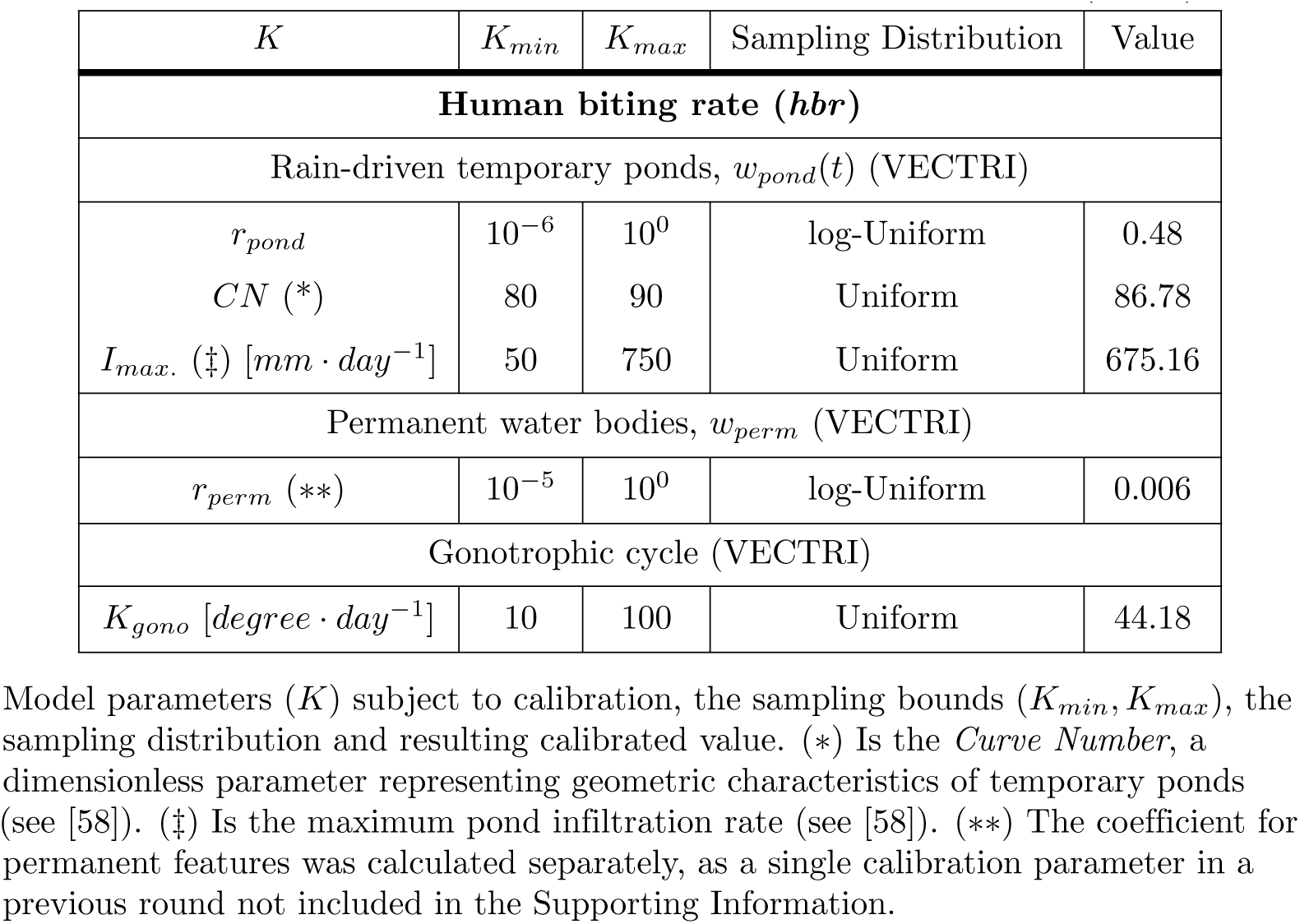
Parameters calibrated during the first fitting round (*Fit 1* ).

| $K$ | $K_{min}$ | $K_{max}$ | Sampling Distribution | Value |
| --- | --- | --- | --- | --- |
| **Human biting rate (*hbr*)** |  |  |  |  |
| Rain-driven temporary ponds, $w_{pond}(t)$ (VECTRI) | | | | |
| $r_{pond}$ | $10^{-6}$ | $10^0$ | log-Uniform | 0.48 |
| $CN$ (*) | 80 | 90 | Uniform | 86.78 |
| $I_{max}$. (‡) [$mm \cdot day^{-1}$] | 50 | 750 | Uniform | 675.16 |
| Permanent water bodies, $w_{perm}$ (VECTRI) | | | | |
| $r_{perm}$ (**) | $10^{-5}$ | $10^0$ | log-Uniform | 0.006 |
| Gonotrophic cycle (VECTRI) |  |  |  |  |
| $K_{gono}$ [$degree \cdot day^{-1}$] | 10 | 100 | Uniform | 44.18 |

Model parameters ( $K$ ) subject to calibration, the sampling bounds ( $K_{min}, K_{max}$ ), the sampling distribution and resulting calibrated value. (\*) Is the *Curve Number*, a dimensionless parameter representing geometric characteristics of temporary ponds (see [58]). (‡) Is the maximum pond infiltration rate (see [58]). (\*\*) The coefficient for permanent features was calculated separately, as a single calibration parameter in a previous round not included in the Supporting Information.

**Table S7.** Parameters calibrated during the second fitting round (*Fit 2* ).

| $K$ | $K_{min}$ | $K_{max}$ | Sampling Distribution | Value |
| --- | --- | --- | --- | --- |
| **Entomological Inoculation Rate (*EIR*)** |  |  |  |  |
| Sporogonic cycle (VECTRI) |  |  |  |  |
| $K_{sporo}$ [$^{\circ}C$ day] | 50 | 150 | Uniform | 122.35 |
| Agent-vector interactions (ABM) |  |  |  |  |
| $K_V$ [$m^{-2}$] | — | — | — | 0.0003<sup>(*)</sup> |
| $b_V$ [bites per vector per day] | — | — | — | 0.5<sup>(*)</sup> |
| $P_{h0}$ | — | — | — | 0.2 [114]<sup>(‡)</sup> |
| $P_{v0}$ | 0.01 | 0.4 | Uniform | 0.34 |
| $P_{V \rightarrow h}^{max.}$ | 0.01 | 0.4 | Uniform | 0.24 |

Model parameters ( $K$ ) subject to calibration, the sampling bounds ( $K_{min}, K_{max}$ ), sampling distribution and resulting calibrated values. <sup>(\*)</sup> were chosen before and kept fixed during calibration. <sup>(‡)</sup> was kept as estimated in referenced source, after a preliminary sensitivity analysis (not shown).

**Table S8.** Parameters calibrated during the second fitting round (*Fit 2* ).

| $K$ | $K_{min}$ | $K_{max}$ | Sampling Distribution | Value |
| --- | --- | --- | --- | --- |
| **Age-structured malaria incidence** |  |  |  |  |
| Symptomatics scheme (ABM) |  |  |  |  |
| $m_a$ | 1 | 100 | Uniform | $10^{(?)}$ |
| $m_c$ | 1 | 100 | Uniform | $60^{(?)}$ |
| $k_m$ | 1 | 60 | Uniform | $10^{(?)}$ |
| $i_a$ | 0 | 1 | Uniform | $0.15^{(?)}$ |
| $i_c$ | 0.4 | 1 | Uniform | 0.986 |
| $k_* [year]$ | 1 | 30 | Uniform | 15.87 |
| $\alpha_{min,0}$ | 0.01 | 0.5 | Uniform | 0.42 |
| $k_\alpha [year]$ | 1 | 20 | Uniform | 7.24 |
| Immunity dynamics (ABM) |  |  |  |  |
| $i_{m0}^+$ | 0.01 | 0.2 | Uniform | 0.197 |
| $i_1$ | 0.01 | 0.5 | log-Uniform | 0.239 |
| $i_2$ | 0.5 | 5 | log-Uniform | 1.727 |
| $i_m^*$ | $10^{-4}$ | $10^{-2}$ | log-Uniform | $0.001^{(\ddagger)}$ |
| $A_1$ | 0.01 | 1.0 | Uniform | 0.989 |
| Mortality and Waiting times (ABM) |  |  |  |  |
| $t_m^{1/2} [month]$ | 3 | 12 | Uniform | $6^{(?)}$ |
| $t_a^{1/2} [day]$ | 3650 | 7300 | Uniform | $3650^{(?)}$ |
| $t_c^{1/2} [day]$ | 50 | 300 | Uniform | $200^{(?)}$ |
| $k_e [year]$ | 1 | 60 | Uniform | $15^{(?)}$ |

Model parameters ( $K$ ) subject to calibration, the sampling bounds ( $K_{min}, K_{max}$ ), sampling distribution and resulting calibrated values. <sup>(?)</sup> these parameters were non-identifiable, this is, points belonging to the Pareto front were uniformly distributed - for this we just chose a value in the interval. <sup>(‡)</sup> was kept at this value before the calibration, from a preliminary sensitivity analysis within the given bounds (not shown).

## References

1. World Health Organization, Malaria, fact sheet. World Health Organization; 2023. Available from: https://www.who.int/news-room/fact-sheets/detail/malaria.

2. Sinka ME, Bangs MJ, Manguin S, Coetzee M, Mbogo CM, Hemingway J, et al. The dominant Anopheles vectors of human malaria in Africa, Europe and the Middle East: occurrence data, distribution maps and bionomic précis. Parasites & Vectors. 2010 Dec;3(1):117. Available from: https://parasitesandvectors.biomedcentral.com/articles/10.1186/1756-3305-3-117. doi:10.1186/1756-3305-3-117.

3. White BJ, Collins FH, Besansky NJ. Evolution of Anopheles gambiae in Relation to Humans and Malaria. Annual Review of Ecology, Evolution, and Systematics. 2011 Dec;42(1):111–32. Available from: https://www.annualreviews.org/doi/10.1146/annurev-ecolsys-102710-145028. doi:10.1146/annurev-ecolsys-102710-145028.

4. Paaijmans KP, Read AF, Thomas MB. Understanding the link between malaria risk and climate. Proceedings of the National Academy of Sciences. 2009 Aug;106(33):13844–9. Available from: https://pnas.org/doi/full/10.1073/pnas.0903423106. doi:10.1073/pnas.0903423106.

5. Yamba EI, Fink AH, Badu K, Asare EO, Tompkins AM, Amekudzi LK. Climate Drivers of Malaria Transmission Seasonality and Their Relative Importance in Sub-Saharan Africa. GeoHealth. 2023 Feb;7(2):e2022GH000698. Available from: https://agupubs.onlinelibrary.wiley.com/doi/10.1029/2022GH000698. doi:10.1029/2022GH000698.

6. Beloconi A, Nyawanda BO, Bigogo G, Khagayi S, Obor D, Danquah I, et al. Malaria, climate variability, and interventions: modelling transmission dynamics. Scientific Reports. 2023 May;13(1):7367. Available from: https://www.nature.com/articles/s41598-023-33868-8. doi:10.1038/s41598-023-33868-8.

7. Doolan DL, Dobaño C, Baird JK. Acquired Immunity to Malaria. Clinical Microbiology Reviews. 2009 Jan;22(1):13–36. Available from: https://journals.asm.org/doi/10.1128/CMR.00025-08. doi:10.1128/CMR.00025-08.

8. Laneri K, Paul RE, Tall A, Faye J, Diene-Sarr F, Sokhna C, et al. Dynamical malaria models reveal how immunity buffers effect of climate variability. Proceedings of the National Academy of Sciences. 2015 Jul;112(28):8786–91. Available from: https://pnas.org/doi/full/10.1073/pnas.1419047112. doi:10.1073/pnas.1419047112.

9. Yamana TK, Bomblies A, Laminou IM, Duchemin JB, Eltahir EAB. Linking environmental variability to village-scale malaria transmission using a simple immunity model. Parasites & Vectors. 2013 Dec;6(1):226. Available from: https://parasitesandvectors.biomedcentral.com/articles/10.1186/1756-3305-6-226. doi:10.1186/1756-3305-6-226.

10. Caminade C, McIntyre KM, Jones AE. Impact of recent and future climate change on vector-borne diseases. Annals of the New York Academy of Sciences. 2019 Jan;1436(1):157–73. Available from: https://nyaspubs.onlinelibrary.wiley.com/doi/10.1111/nyas.13950. doi:10.1111/nyas.13950.

11. Caminade C, Kovats S, Rocklov J, Tompkins AM, Morse AP, Colón-González FJ, et al. Impact of climate change on global malaria distribution. Proceedings of the National Academy of Sciences. 2014 Mar;111(9):3286–91. Available from: https://pnas.org/doi/full/10.1073/pnas.1302089111. doi:10.1073/pnas.1302089111.

12. Symons TL, Moran A, Balzarolo A, Vargas C, Robertson M, Lubinda J, et al. Projected impacts of climate change on malaria in Africa. Nature. 2026 Jan. Available from: https://www.nature.com/articles/s41586-025-10015-z. doi:10.1038/s41586-025-10015-z.

13. Villena OC, Arab A, Lippi CA, Ryan SJ, Johnson LR. Influence of environmental, geographic, socio-demographic, and epidemiological factors on presence of malaria at the community level in two continents. Scientific Reports. 2024 Jul;14(1):16734. Available from: https://www.nature.com/articles/s41598-024-67452-5. doi:10.1038/s41598-024-67452-5.

14. Walker PGT, Griffin JT, Ferguson NM, Ghani AC. Estimating the most efficient allocation of interventions to achieve reductions in Plasmodium falciparum malaria burden and transmission in Africa: a modelling study. The Lancet Global Health. 2016 Jul;4(7):e474–84. Available from: https://linkinghub.elsevier.com/retrieve/pii/S2214109X16300730. doi:10.1016/S2214-109X(16)30073-0.

15. Agusto FB, Gumel AB, Parham PE. QUALITATIVE ASSESSMENT OF THE ROLE OF TEMPERATURE VARIATIONS ON MALARIA TRANSMISSION DYNAMICS. Journal of Biological Systems. 2015 Dec;23(04):1550030. Available from: https://doi.org/10.1142/S0218339015500308. doi:10.1142/S0218339015500308.

16. Okuneye K, Gumel AB. Analysis of a temperature- and rainfall-dependent model for malaria transmission dynamics. Mathematical Biosciences. 2017 May;287:72–92. Available from: https://www.sciencedirect.com/science/article/pii/S0025556416300177. doi:10.1016/j.mbs.2016.03.013.

17. Smith NR, Trauer JM, Gambhir M, Richards JS, Maude RJ, Keith JM, et al. Agent-based models of malaria transmission: a systematic review. Malaria Journal. 2018 Dec;17(1):299. Available from: https://malariajournal.biomedcentral.com/articles/10.1186/s12936-018-2442-y. doi:10.1186/s12936-018-2442-y.

18. Smith T, Killeen GF, Maire N, Ross A, Molineaux L, Tediosi F, et al. MATHEMATICAL MODELING OF THE IMPACT OF MALARIA VACCINES ON THE CLINICAL EPIDEMIOLOGY AND NATURAL HISTORY OF PLASMODIUM FALCIPARUM MALARIA: OVERVIEW. The American Journal of Tropical Medicine and Hygiene. 2006 Aug;75(2 suppl):1–10. Available from: https://www.ajtmh.org/view/journals/tpmd/75/2_suppl/article-p1.xml. doi:10.4269/ajtmh.2006.75.2suppl.0750001.

19. Gurarie D, McKenzie FE. A stochastic model of immune-modulated malaria infection and disease in children. Mathematical Biosciences. 2007 Dec;210(2):576–97. Available from: https://www.sciencedirect.com/science/article/pii/S0025556407001472. doi:10.1016/j.mbs.2007.07.001.

20. Griffin JT, Hollingsworth TD, Okell LC, Churcher TS, White M, Hinsley W, et al. Reducing Plasmodium falciparum Malaria Transmission in Africa: A Model-Based Evaluation of Intervention Strategies. PLoS Medicine. 2010 Aug;7(8):e1000324. Available from: https://dx.plos.org/10.1371/journal.pmed.1000324. doi:10.1371/journal.pmed.1000324.

21. Zhu L, Marshall JM, Qualls WA, Schlein Y, McManus JW, Arheart KL, et al. Modelling optimum use of attractive toxic sugar bait stations for effective malaria vector control in Africa. Malaria Journal. 2015 Dec;14(1):492. Available from: https://malariajournal.biomedcentral.com/articles/10.1186/s12936-015-1012-9. doi:10.1186/s12936-015-1012-9.

22. Gu W, Novak RJ. Predicting the impact of insecticide-treated bed nets on malaria transmission: the devil is in the detail. Malaria Journal. 2009 Dec;8(1):256. Available from: https://malariajournal.biomedcentral.com/articles/10.1186/1475-2875-8-256. doi:10.1186/1475-2875-8-256.

23. Maire N, Shillcutt SD, Walker DG, Tediosi F, Smith TA. Cost-Effectiveness of the Introduction of a Pre-Erythrocytic Malaria Vaccine into the Expanded Program on Immunization in Sub-Saharan Africa: Analysis of Uncertainties Using a Stochastic Individual-Based Simulation Model of Plasmodium falciparum Malaria. Value in Health. 2011 Dec;14(8):1028–38. Available from: https://linkinghub.elsevier.com/retrieve/pii/S1098301511015269. doi:10.1016/j.jval.2011.06.004.

24. Nguyen TD, Olliaro P, Dondorp AM, Baird JK, Lam HM, Farrar J, et al. Optimum population-level use of artemisinin combination therapies: a modelling study. The Lancet Global Health. 2015 Dec;3(12):e758–66. Available from: https://linkinghub.elsevier.com/retrieve/pii/S2214109X1500162X. doi:10.1016/S2214-109X(15)00162-X.

25. Karl S, White MT, Milne GJ, Gurarie D, Hay SI, Barry AE, et al. Spatial Effects on the Multiplicity of Plasmodium falciparum Infections. PLOS ONE. 2016 Oct;11(10):e0164054. Available from: https://dx.plos.org/10.1371/journal.pone.0164054. doi:10.1371/journal.pone.0164054.

26. Chitnis N, Hardy D, Smith T. A Periodically-Forced Mathematical Model for the Seasonal Dynamics of Malaria in Mosquitoes. Bulletin of Mathematical Biology. 2012 May;74(5):1098–124. Available from: http://link.springer.com/10.1007/s11538-011-9710-0. doi:10.1007/s11538-011-9710-0.

27. Bomblies A, Duchemin J, Eltahir EAB. Hydrology of malaria: Model development and application to a Sahelian village. Water Resources Research. 2008 Dec;44(12):2008WR006917. Available from: https://agupubs.onlinelibrary.wiley.com/doi/10.1029/2008WR006917. doi:10.1029/2008WR006917.

28. Eckhoff PA. A malaria transmission-directed model of mosquito life cycle and ecology. Malaria Journal. 2011 Dec;10(1):303. Available from: https://malariajournal.biomedcentral.com/articles/10.1186/1475-2875-10-303. doi:10.1186/1475-2875-10-303.

29. Depinay JMO, Mbogo CM, Killeen G, Knols B, Beier J, Carlson J, et al. A simulation model of African Anopheles ecology and population dynamics for the analysis of malaria transmission. Malaria Journal. 2004 Jul;3(1):29. Available from: https://doi.org/10.1186/1475-2875-3-29. doi:10.1186/1475-2875-3-29.

30. Stryker JJ, Bomblies A. The Impacts of Land Use Change on Malaria Vector Abundance in a Water-Limited, Highland Region of Ethiopia. EcoHealth. 2012 Dec;9(4):455–70. Available from: https://doi.org/10.1007/s10393-012-0801-7. doi:10.1007/s10393-012-0801-7.

31. Martin-Makowka A, Munday JD, Tompkins AM, Caminade C, Chitnis N. Modelling the impact of climate variability on the effectiveness of seasonal indoor residual spraying.

32. Tompkins AM, Ermert V. A regional-scale, high resolution dynamical malaria model that accounts for population density, climate and surface hydrology. Malaria Journal. 2013 Dec;12(1):65. Available from: https://malariajournal.biomedcentral.com/articles/10.1186/1475-2875-12-65. doi:10.1186/1475-2875-12-65.

33. Gerardin J, Bever CA, Hamainza B, Miller JM, Eckhoff PA, Wenger EA. Optimal Population-Level Infection Detection Strategies for Malaria Control and Elimination in a Spatial Model of Malaria Transmission. PLOS Computational Biology. 2016 Jan;12(1):e1004707. Available from: https://dx.plos.org/10.1371/journal.pcbi.1004707. doi:10.1371/journal.pcbi.1004707.

34. Silal SP, Little F, Barnes KI, White LJ. Predicting the impact of border control on malaria transmission: a simulated focal screen and treat campaign. Malaria Journal. 2015 Dec;14(1):268. Available from: http://www.malariajournal.com/content/14/1/268. doi:10.1186/s12936-015-0776-2.

35. Eckhoff PA, Wenger EA, Godfray HCJ, Burt A. Impact of mosquito gene drive on malaria elimination in a computational model with explicit spatial and temporal dynamics. Proceedings of the National Academy of Sciences. 2017 Jan;114(2). Available from: https://pnas.org/doi/full/10.1073/pnas.1611064114. doi:10.1073/pnas.1611064114.

36. Pizzitutti F, Pan W, Feingold B, Zaitchik B, Álvarez CA, Mena CF. Out of the net: An agent-based model to study human movements influence on local-scale malaria transmission. PLOS ONE. 2018 Mar;13(3):e0193493. Available from: https://dx.plos.org/10.1371/journal.pone.0193493. doi:10.1371/journal.pone.0193493.

37. Gerardin J, Bever CA, Bridenbecker D, Hamainza B, Silumbe K, Miller JM, et al. Effectiveness of reactive case detection for malaria elimination in three archetypical transmission settings: a modelling study. Malaria Journal. 2017 Dec;16(1):248. Available from: https://malariajournal.biomedcentral.com/articles/10.1186/s12936-017-1903-z. doi:10.1186/s12936-017-1903-z.

38. Weiss DJ, Lucas TCD, Nguyen M, Nandi AK, Bisanzio D, Battle KE, et al. Mapping the global prevalence, incidence, and mortality of Plasmodium falciparum, 2000–17: a spatial and temporal modelling study. The Lancet. 2019 Jul;394(10195):322–31. Available from: https://linkinghub.elsevier.com/retrieve/pii/S0140673619310979. doi:10.1016/S0140-6736(19)31097-9.

39. Hay SI, Rogers DJ, Toomer JF, Snow RW. Annual Plasmodium falciparum entomological inoculation rates (EIR) across Africa: literature survey, internet access and review. Transactions of the Royal Society of Tropical Medicine and Hygiene. 2000 Mar;94(2):113–27. Available from: https://academic.oup.com/trstmh/article-lookup/doi/10.1016/S0035-9203(00)90246-3. doi:10.1016/S0035-9203(00)90246-3.

40. Plan stratégique national de lutte contre le paludisme au Sénégal, 2021-2025. Ministére de la Santé et de l’Action Sociale. Programme National de Lutte contre le Paludisme (PNLP); 2020.

41. Grimm V, Berger U, Bastiansen F, Eliassen S, Ginot V, Giske J, et al. A standard protocol for describing individual-based and agent-based models. Ecological Modelling. 2006 Sep;198(1):115–26. Available from: https://www.sciencedirect.com/science/article/pii/S0304380006002043. doi:10.1016/j.ecolmodel.2006.04.023.

42. Grimm V, Berger U, DeAngelis DL, Polhill JG, Giske J, Railsback SF. The ODD protocol: A review and first update. Ecological Modelling. 2010 Nov;221(23):2760–8. Available from: https://linkinghub.elsevier.com/retrieve/pii/S030438001000414X. doi:10.1016/j.ecolmodel.2010.08.019.

43. Cowman AF, Healer J, Marapana D, Marsh K. Malaria: Biology and Disease. Cell. 2016 Oct;167(3):610–24. Available from: https://linkinghub.elsevier.com/retrieve/pii/S009286741631008X. doi:10.1016/j.cell.2016.07.055.

44. Griffin JT, Hollingsworth TD, Reyburn H, Drakeley CJ, Riley EM, Ghani AC. Gradual acquisition of immunity to severe malaria with increasing exposure. Proceedings of the Royal Society B: Biological Sciences. 2015 Feb;282(1801):20142657. Available from: https://royalsocietypublishing.org/doi/10.1098/rspb.2014.2657. doi:10.1098/rspb.2014.2657.

45. Gupta S, Snow RW, Donnelly CA, Marsh K, Newbold C. Immunity to non-cerebral severe malaria is acquired after one or two infections. Nature Medicine. 1999 Mar;5(3):340–3. Available from: https://www.nature.com/articles/nm0399_340. doi:10.1038/6560.

46. Ghani AC, Sutherland CJ, Riley EM, Drakeley CJ, Griffin JT, Gosling RD, et al. Loss of Population Levels of Immunity to Malaria as a Result of Exposure-Reducing Interventions: Consequences for Interpretation of Disease Trends. PLoS ONE. 2009 Feb;4(2):e4383. Available from: https://dx.plos.org/10.1371/journal.pone.0004383. doi:10.1371/journal.pone.0004383.

47. Bousema T, Okell L, Felger I, Drakeley C. Asymptomatic malaria infections: detectability, transmissibility and public health relevance. Nature Reviews Microbiology. 2014 Dec;12(12):833–40. Available from: https://www.nature.com/articles/nrmicro3364. doi:10.1038/nrmicro3364.

48. Eikenberry SE, Gumel AB. Mathematical modeling of climate change and malaria transmission dynamics: a historical review. Journal of Mathematical Biology. 2018 Oct;77(4):857–933. Available from: http://link.springer.com/10.1007/s00285-018-1229-7. doi:10.1007/s00285-018-1229-7.

49. Sama W, Dietz K, Smith T. Distribution of survival times of deliberate Plasmodium falciparum infections in tertiary syphilis patients. Transactions of the Royal Society of Tropical Medicine and Hygiene. 2006 Sep;100(9):811–6. Available from: https://academic.oup.com/trstmh/article-lookup/doi/10.1016/j.trstmh.2005.11.001. doi:10.1016/j.trstmh.2005.11.001.

50. Bretscher MT, Maire N, Chitnis N, Felger I, Owusu-Agyei S, Smith T. The distribution of Plasmodium falciparum infection durations. Epidemics. 2011 Jun;3(2):109–18. Available from: https://linkinghub.elsevier.com/retrieve/pii/S175543651100020X. doi:10.1016/j.epidem.2011.03.002.

51. Schneider P, Schoone G, Schallig H, Verhage D, Telgt D, Eling W, et al. Quantification of Plasmodium falciparum gametocytes in differential stages of development by quantitative nucleic acid sequence-based amplification. Molecular and Biochemical Parasitology. 2004 Sep;137(1):35–41. Available from: https://www.sciencedirect.com/science/article/pii/S0166685104001379. doi:10.1016/j.molbiopara.2004.03.018.

52. Farid R, Dixon MW, Tilley L, McCarthy JS. Initiation of gametocytogenesis at very low parasite density in Plasmodium falciparum infection. The Journal of Infectious Diseases. 2017 Apr;215(7):1167–74. Available from: https://academic.oup.com/jid/article/215/7/1167/3813451. doi:10.1093/infdis/jix035.

53. Mosquito Eggs. NIAID Visual & Medical Arts. (10/7/2024). Mosquito Eggs. NIAID NIH BIOART Source. bioart.niaid.nih.gov/bioart/362;.

54. Mosquito Larva. NIAID Visual & Medical Arts. (10/7/2024). Mosquito Larva. NIAID NIH BIOART Source. bioart.niaid.nih.gov/bioart/366;.

55. Anopheles. NIAID Visual & Medical Arts. (10/7/2024). Anopheles. NIAID NIH BIOART Source. bioart.niaid.nih.gov/bioart/16;.

56. Garrido Zornoza M, Caminade C, Tompkins AM. The effect of climate change and temperature extremes on Aedes albopictus populations: a regional case study for Italy. Journal of The Royal Society Interface. 2024 Nov;21(220):20240319. Available from: https://royalsocietypublishing.org/doi/10.1098/rsif.2024.0319. doi:10.1098/rsif.2024.0319.

57. Asare EO, Tompkins AM, Amekudzi LK, Ermert V. A breeding site model for regional, dynamical malaria simulations evaluated using in situ temporary ponds observations. Geospatial Health. 2016 Mar;11(1s). Available from: http://www.geospatialhealth.net/index.php/gh/article/view/390. doi:10.4081/gh.2016.390.

58. Asare EO, Tompkins AM, Bomblies A. A Regional Model for Malaria Vector Developmental Habitats Evaluated Using Explicit, Pond-Resolving Surface Hydrology Simulations. PLOS ONE. 2016 Mar;11(3):e0150626. Available from: https://dx.plos.org/10.1371/journal.pone.0150626. doi:10.1371/journal.pone.0150626.

59. Akiba T, Sano S, Yanase T, Ohta T, Koyama M. Optuna: A Next-generation Hyperparameter Optimization Framework. In: Proceedings of the 25th ACM SIGKDD International Conference on Knowledge Discovery & Data Mining. Anchorage AK USA: ACM; 2019. p. 2623–31. Available from: https://dl.acm.org/doi/10.1145/3292500.3330701. doi:10.1145/3292500.3330701.

60. Trape JF, Tall A, Sokhna C, Ly AB, Diagne N, Ndiath O, et al. The rise and fall of malaria in a west African rural community, Dielmo, Senegal, from 1990 to 2012: a 22 year longitudinal study. The Lancet Infectious Diseases. 2014 Jun;14(6):476–88. Available from: https://linkinghub.elsevier.com/retrieve/pii/S1473309914707121. doi:10.1016/S1473-3099(14)70712-1.

61. Yamba EI, Tompkins AM, Fink AH, Ermert V, Amelie MD, Amekudzi LK, et al. Monthly Entomological Inoculation Rate Data for Studying the Seasonality of Malaria Transmission in Africa. Data. 2020 Mar;5(2):31. Available from: https://www.mdpi.com/2306-5729/5/2/31. doi:10.3390/data5020031.

62. Robert V, Dieng H, Lochouarn L, Traoré SF, Trape Jimondon F, et al. La transmission du paludisme dans la zone de Niakhar, Sénégal. Tropical Medicine & International Health. 1998 Aug;3(8):667–77. Available from: https://onlinelibrary.wiley.com/doi/10.1046/j.1365-3156.1998.00288.x. doi:10.1046/j.1365-3156.1998.00288.x.

63. Pfeffer DA, Lucas TCD, May D, Harris J, Rozier J, Twohig KA, et al. malariaAtlas: an R interface to global malariometric data hosted by the Malaria Atlas Project. Malaria Journal. 2018 Dec;17(1):352. Available from: https://malariajournal.biomedcentral.com/articles/10.1186/s12936-018-2500-5. doi:10.1186/s12936-018-2500-5.

64. Steketee RW, Campbell CC. Impact of national malaria control scale-up programmes in Africa: magnitude and attribution of effects. Malaria Journal. 2010 Dec;9(1):299. Available from: https://malariajournal.biomedcentral.com/articles/10.1186/1475-2875-9-299. doi:10.1186/1475-2875-9-299.

65. Verdin A, Funk C, Peterson P, Landsfeld M, Tuholske C, Grace K. Development and validation of the CHIRTS-daily quasi-global high-resolution daily temperature data set. Scientific Data. 2020 Sep;7(1):303. Available from: https://www.nature.com/articles/s41597-020-00643-7. doi:10.1038/s41597-020-00643-7.

66. Funk C, Peterson P, Landsfeld M, Pedreros D, Verdin J, Shukla S, et al. The climate hazards infrared precipitation with stations—a new environmental record for monitoring extremes. Scientific Data. 2015 Dec;2(1):150066. Available from: https://www.nature.com/articles/sdata201566. doi:10.1038/sdata.2015.66.

67. Bondarenko M, Priyatikanto R, Tejedor-Garavito N, Zhang W, McKeen T, Cunningham A, et al. The spatial distribution of population broken down by gender and age groupings in 2015-2030 at a resolution of 30 arc (approximately 1km at the equator) R2025A version v1. WorldPop - School of Geography and Environmental Science, University of Southampton; 2025. Available from: https://doi.org/10.5258/SOTON/WP00846. doi:10.5258/SOTON/WP00846. doi:10.5258/SOTON/WP00846.

68. PNLP. Bulletin Epidémiologique Annuel du paludisme au Sénégal en 2019. 2020.

69. Carneiro I, Roca-Feltrer A, Griffin JT, Smith L, Tanner M, Schellenberg JA, et al. Age-Patterns of Malaria Vary with Severity, Transmission Intensity and Seasonality in Sub-Saharan Africa: A Systematic Review and Pooled Analysis. PLoS ONE. 2010 Feb;5(2):e8988. Available from: https://dx.plos.org/10.1371/journal.pone.0008988. doi:10.1371/journal.pone.0008988.

70. Okell L, Ghani A, Lyons E, Drakeley C. Submicroscopic Infection in Plasmodium falciparum –Endemic Populations: A Systematic Review and Meta-Analysis. The Journal of Infectious Diseases. 2009 Nov;200(10):1509–17. Available from: https://academic.oup.com/jid/article-lookup/doi/10.1086/644781. doi:10.1086/644781.

71. Okell LC, Bousema T, Griffin JT, Ouédraogo AL, Ghani AC, Drakeley CJ. Factors determining the occurrence of submicroscopic malaria infections and their relevance for control. Nature Communications. 2012 Dec;3(1):1237. Available from: https://www.nature.com/articles/ncomms2241. doi:10.1038/ncomms2241.

72. Fall P, Diouf I, Deme A, Sene D. Assessment of Climate-Driven Variations in Malaria Transmission in Senegal Using the VECTRI Model. Atmosphere. 2022 Mar;13(3):418. Available from: https://www.mdpi.com/2073-4433/13/3/418. doi:10.3390/atmos13030418.

73. Diouf I, Ndione JA, Gaye AT. Malaria in Senegal: Recent and Future Changes Based on Bias-Corrected CMIP6 Simulations. Tropical Medicine and Infectious Disease. 2022 Nov;7(11):345. Available from: https://www.mdpi.com/2414-6366/7/11/345. doi:10.3390/tropicalmed7110345.

74. Guelbéogo WM, Gonçalves BP, Grignard L, Bradley J, Serme SS, Hellewell J, et al. Variation in natural exposure to anopheles mosquitoes and its effects on malaria transmission. eLife. 2018 Jan;7:e32625. Available from: https://elifesciences.org/articles/32625. doi:10.7554/eLife.32625.

75. Faye MB, Dia AK, Diouf AF, Moussa B, Coulibaly Y, Diop M, et al. Role of Anopheles funestus in sustaining Residual Malaria Transmission in Central Senegal. 2026.

76. Torcia MG, Santarlasci V, Cosmi L, Clemente A, Maggi L, Mangano VD, et al. Functional deficit of T regulatory cells in Fulani, an ethnic group with low susceptibility to Plasmodium falciparum malaria. Proceedings of the National Academy of Sciences. 2008 Jan;105(2):646–51. Available from: https://pnas.org/doi/full/10.1073/pnas.0709969105. doi:10.1073/pnas.0709969105.

77. Camponovo F, Lee TE, Russell JR, Burgert L, Gerardin J, Penny MA. Mechanistic within-host models of the asexual Plasmodium falciparum infection: a review and analytical assessment. Malaria Journal. 2021 Dec;20(1):309. Available from: https://malariajournal.biomedcentral.com/articles/10.1186/s12936-021-03813-z. doi:10.1186/s12936-021-03813-z.

78. Collins KA, Wang CYT, Adams M, Mitchell H, Rampton M, Elliott S, et al. A controlled human malaria infection model enabling evaluation of transmission-blocking interventions. Journal of Clinical Investigation. 2018 Apr;128(4):1551–62. Available from: https://www.jci.org/articles/view/98012. doi:10.1172/JCI98012.

79. Cao P, Collins KA, Zaloumis S, Wattanakul T, Tarning J, Simpson JA, et al. Modeling the dynamics of Plasmodium falciparum gametocytes in humans during malaria infection. eLife. 2019 Oct;8:e49058. Available from: https://elifesciences.org/articles/49058. doi:10.7554/eLife.49058.

80. Parham PE, Michael E. Modeling the Effects of Weather and Climate Change on Malaria Transmission. Environmental Health Perspectives. 2010 May;118(5):620–6. Available from: https://pubs.acs.org/evhpaz/article-lookup/doi/10.1289/ehp.0901256. doi:10.1289/ehp.0901256.

81. Unisex Icon. NIAID Visual & Medical Arts. (10/7/2024). Unisex Icon. NIAID NIH BIOART Source. bioart.niaid.nih.gov/bioart/13;.

82. Kelly-Hope LA, McKenzie FE. The multiplicity of malaria transmission: a review of entomological inoculation rate measurements and methods across sub-Saharan Africa. Malaria Journal. 2009 Dec;8(1):19. Available from: https://malariajournal.biomedcentral.com/articles/10.1186/1475-2875-8-19. doi:10.1186/1475-2875-8-19.

83. Ototo EN, Mbugi JP, Wanjala CL, Zhou G, Githeko AK, Yan G. Surveillance of malaria vector population density and biting behaviour in western Kenya. Malaria Journal. 2015 Dec;14(1):244. Available from: https://malariajournal.biomedcentral.com/articles/10.1186/s12936-015-0763-7. doi:10.1186/s12936-015-0763-7.

84. Nzioki I, Machani MG, Onyango SA, Kabui KK, Githeko AK, Ochomo E, et al. Differences in malaria vector biting behavior and changing vulnerability to malaria transmission in contrasting ecosystems of western Kenya. Parasites & Vectors. 2023 Oct;16(1):376. Available from: https://parasitesandvectors.biomedcentral.com/articles/10.1186/s13071-023-05944-5. doi:10.1186/s13071-023-05944-5.

85. Sangbakembi-Ngounou C, Costantini C, Longo-Pendy NM, Ngoagouni C, Akone-Ella O, Rahola N, et al. Diurnal biting of malaria mosquitoes in the Central African Republic indicates residual transmission may be “out of control”. Proceedings of the National Academy of Sciences. 2022 May;119(21):e2104282119. Available from: https://pnas.org/doi/full/10.1073/pnas.2104282119. doi:10.1073/pnas.2104282119.

86. Cooke MK, Kahindi SC, Oriango RM, Owaga C, Ayoma E, Mabuka D, et al. ‘A bite before bed’: exposure to malaria vectors outside the times of net use in the highlands of western Kenya. Malaria Journal. 2015 Dec;14(1):259. Available from: http://www.malariajournal.com/content/14/1/259. doi:10.1186/s12936-015-0766-4.

87. Sherrard-Smith E, Skarp JE, Beale AD, Fornadel C, Norris LC, Moore SJ, et al. Mosquito feeding behavior and how it influences residual malaria transmission across Africa. Proceedings of the National Academy of Sciences. 2019 Jul;116(30):15086–95. Available from: https://pnas.org/doi/full/10.1073/pnas.1820646116. doi:10.1073/pnas.1820646116.

88. Monroe A, Moore S, Koenker H, Lynch M, Ricotta E. Measuring and characterizing night time human behaviour as it relates to residual malaria transmission in sub-Saharan Africa: a review of the published literature. Malaria Journal. 2019 Dec;18(1):6. Available from: https://malariajournal.biomedcentral.com/articles/10.1186/s12936-019-2638-9. doi:10.1186/s12936-019-2638-9.

89. Tchole AIM, Ye RZ, Xu Q, Li ZW, Liu JY, Wang SS, et al. Epidemiological behaviour and interventions of malaria in Niger, 2010–2019: a time-series analysis of national surveillance data. Malaria Journal. 2024 Jan;23(1):30. Available from: https://malariajournal.biomedcentral.com/articles/10.1186/s12936-024-04835-z. doi:10.1186/s12936-024-04835-z.

90. Umugwaneza A, Mutsaers M, Ngabonziza JCS, Kattenberg JH, Uwimana A, Ahmed A, et al. Half-decade of scaling up malaria control: malaria trends and impact of interventions from 2018 to 2023 in Rwanda. Malaria Journal. 2025 Feb;24(1):40. Available from: https://malariajournal.biomedcentral.com/articles/10.1186/s12936-025-05278-w. doi:10.1186/s12936-025-05278-w.

91. Atusingwize E, Deane K, Musoke D. Social determinants of malaria in low- and middle-income countries: a mixed-methods systematic review. Malaria Journal. 2025 May;24(1):165. Available from: https://malariajournal.biomedcentral.com/articles/10.1186/s12936-025-05407-5. doi:10.1186/s12936-025-05407-5.

92. Sondo P, Derra K, Lefevre T, Diallo-Nakanabo S, Tarnagda Z, Zampa O, et al. Genetically diverse Plasmodium falciparum infections, within-host competition and symptomatic malaria in humans. Scientific Reports. 2019 Jan;9(1):127. Available from: https://www.nature.com/articles/s41598-018-36493-y. doi:10.1038/s41598-018-36493-y.

93. Eldh M, Hammar U, Arnot D, Beck HP, Garcia A, Liljander A, et al. Multiplicity of Asymptomatic Plasmodium falciparum Infections and Risk of Clinical Malaria: A Systematic Review and Pooled Analysis of Individual Participant Data. The Journal of Infectious Diseases. 2020 Feb;221(5):775–85. Available from: https://academic.oup.com/jid/article/221/5/775/5581561. doi:10.1093/infdis/jiz510.

94. Montell C. The sensory arsenal mosquitoes use to find us. Trends in Parasitology. 2025 Jul;41(7):591–602. Available from: https://linkinghub.elsevier.com/retrieve/pii/S1471492225001357. doi:10.1016/j.pt.2025.05.004.

95. Ellwanger JH, Cardoso JDC, Chies JAB. Variability in human attractiveness to mosquitoes. Current Research in Parasitology & Vector-Borne Diseases. 2021;1:100058. Available from: https://linkinghub.elsevier.com/retrieve/pii/S2667114X21000522. doi:10.1016/j.crpvbd.2021.100058.

96. Bousema T, Drakeley C. Epidemiology and Infectivity of Plasmodium falciparum and Plasmodium vivax Gametocytes in Relation to Malaria Control and Elimination. Clinical Microbiology Reviews. 2011 Apr;24(2):377–410. Available from: https://journals.asm.org/doi/10.1128/CMR.00051-10. doi:10.1128/CMR.00051-10.

97. Churcher TS, Bousema T, Walker M, Drakeley C, Schneider P, Ouédraogo AL, et al. Predicting mosquito infection from Plasmodium falciparum gametocyte density and estimating the reservoir of infection. eLife. 2013 May;2:e00626. Available from: https://elifesciences.org/articles/00626. doi:10.7554/eLife.00626.

98. Akpogheneta OJ, Duah NO, Tetteh KKA, Dunyo S, Lanar DE, Pinder M, et al. Duration of Naturally Acquired Antibody Responses to Blood-Stage Plasmodium falciparum Is Age Dependent and Antigen Specific. Infection and Immunity. 2008 Apr;76(4):1748–55. Available from: https://journals.asm.org/doi/10.1128/IAI.01333-07. doi:10.1128/IAI.01333-07.

99. Moncunill G, Mayor A, Bardají A, Puyol L, Nhabomba A, Barrios D, et al. Cytokine Profiling in Immigrants with Clinical Malaria after Extended Periods of Interrupted Exposure to Plasmodium falciparum. PLoS ONE. 2013 Aug;8(8):e73360. Available from: https://dx.plos.org/10.1371/journal.pone.0073360. doi:10.1371/journal.pone.0073360.

100. Trape JF, Rogier C, Konate L, Diagne N, Bouganali H, Canque B, et al. The Dielmo Project: a Longitudinal Study of Natural Malaria Infection and the Mechanisms of Protective Immunity in a Community Living in a Holoendemic Area of Senegal. The American Journal of Tropical Medicine and Hygiene. 1994 Aug;51(2):123–37. Available from: https://www.ajtmh.org/view/journals/tpmd/51/2/article-p123.xml. doi:10.4269/ajtmh.1994.51.123.

101. Felger I, Maire M, Bretscher MT, Falk N, Tiaden A, Sama W, et al. The Dynamics of Natural Plasmodium falciparum Infections. PLoS ONE. 2012 Sep;7(9):e45542. Available from: https://dx.plos.org/10.1371/journal.pone.0045542. doi:10.1371/journal.pone.0045542.

102. Dal-Bianco MP, Köster KB, Kombila UD, Kun JFJ, Grobusch MP, Ngoma GM, et al. High Prevalence of Asymptomatic Plasmodium falciparum Infection in Gabonese Adults. The American Journal of Tropical Medicine and Hygiene. 2007 Nov;77(5):939–42. Available from: https://www.ajtmh.org/view/journals/tpmd/77/5/article-p939.xml. doi:10.4269/ajtmh.2007.77.939.

103. Salgado C, Ayodo G, Macklin MD, Gould MP, Nallandhighal S, Odhiambo EO, et al. The prevalence and density of asymptomatic Plasmodium falciparum infections among children and adults in three communities of western Kenya. Malaria Journal. 2021 Dec;20(1):371. Available from: https://malariajournal.biomedcentral.com/articles/10.1186/s12936-021-03905-w. doi:10.1186/s12936-021-03905-w.

104. Agaba BB, Rugera SP, Mpirirwe R, Atekat M, Okubal S, Masereka K, et al. Asymptomatic malaria infection, associated factors and accuracy of diagnostic tests in a historically high transmission setting in Northern Uganda. Malaria Journal. 2022 Dec;21(1):392. Available from: https://malariajournal.biomedcentral.com/articles/10.1186/s12936-022-04421-1. doi:10.1186/s12936-022-04421-1.

105. Gatton ML, Cheng Q. Evaluation of the pyrogenic threshold for Plasmodium falciparum malaria in naive individuals. The American journal of tropical medicine and hygiene. 2002 May;66(5):467–73. Available from: https://www.ajtmh.org/view/journals/tpmd/66/5/article-p467.xml. doi:10.4269/ajtmh.2002.66.467.

106. Rogier C, Commenges D, Trape JF. Evidence for an Age-Dependent Pyrogenic Threshold of Plasmodium falciparum Parasitemia in Highly Endemic Populations. The American Journal of Tropical Medicine and Hygiene. 1996 Jun;54(6):613–9. Available from: https://www.ajtmh.org/view/journals/tpmd/54/6/article-p613.xml. doi:10.4269/ajtmh.1996.54.613.

107. White NJ. Malaria parasite clearance. Malaria Journal. 2017 Dec;16(1):88. Available from: http://malariajournal.biomedcentral.com/articles/10.1186/s12936-017-1731-1. doi:10.1186/s12936-017-1731-1.

108. Wilson DW, Langer C, Goodman CD, McFadden GI, Beeson JG. Defining the Timing of Action of Antimalarial Drugs against Plasmodium falciparum. Antimicrobial Agents and Chemotherapy. 2013 Mar;57(3):1455–67. Available from: https://journals.asm.org/doi/10.1128/AAC.01881-12. doi:10.1128/AAC.01881-12.

109. Detinova TS, Bertram DS, World Health Organization. Age-grouping methods in Diptera of medical importance, with special reference to some vectors of malaria. World Health Organization; 1962.

110. Kristan M, Fleischmann H, Della Torre A, Stich A, Curtis CF. Pyrethroid resistance/susceptibility and differential urban/rural distribution of Anopheles arabiensis and An. gambiae s.s. malaria vectors in Nigeria and Ghana. Medical and Veterinary Entomology. 2003 Sep;17(3):326–32. Available from: https://resjournals.onlinelibrary.wiley.com/doi/10.1046/j.1365-2915.2003.00449.x. doi:10.1046/j.1365-2915.2003.00449.x.

111. Hinne IA, Attah SK, Mensah BA, Forson AO, Afrane YA. Larval habitat diversity and Anopheles mosquito species distribution in different ecological zones in Ghana. Parasites & Vectors. 2021 Apr;14(1):193. Available from: https://parasitesandvectors.biomedcentral.com/articles/10.1186/s13071-021-04701-w. doi:10.1186/s13071-021-04701-w.

112. Kahamba NF, Finda M, Ngowo HS, Msugupakulya BJ, Baldini F, Koekemoer LL, et al. Using ecological observations to improve malaria control in areas where Anopheles funestus is the dominant vector. Malaria Journal. 2022 Dec;21(1):158. Available from: https://malariajournal.biomedcentral.com/articles/10.1186/s12936-022-04198-3. doi:10.1186/s12936-022-04198-3.

113. Nambunga IH, Ngowo HS, Mapua SA, Hape EE, Msugupakulya BJ, Msaky DS, et al. Aquatic habitats of the malaria vector Anopheles funestus in rural south-eastern Tanzania. Malaria Journal. 2020 Dec;19(1):219. Available from: https://malariajournal.biomedcentral.com/articles/10.1186/s12936-020-03295-5. doi:10.1186/s12936-020-03295-5.

114. Ermert V, Fink AH, Jones AE, Morse AP. Development of a new version of the Liverpool Malaria Model. I. Refining the parameter settings and mathematical formulation of basic processes based on a literature review. Malaria Journal. 2011 Dec;10(1):35. Available from: https://malariajournal.biomedcentral.com/articles/10.1186/1475-2875-10-35. doi:10.1186/1475-2875-10-35.

